# Falling short: Inadequate target attainment in guideline-based therapy of nontuberculous mycobacterial disease

**DOI:** 10.64898/2026.09.24.26363927

**Authors:** Trevor Jaan Shoaf, Tyler Dierckman, Noah Strawhacker, Marina de Lorena Muniz Surani, Meghna Nair, Erika Bawab, Patricio Escalante, Thomas Dick, Véronique Dartois, Elsje Pienaar

## Abstract

Nontuberculous mycobacterial (NTM) infections are a scourge that affect growing patient populations due to the rise of risk factors associated with these opportunistic pathogens. Cure rates are unacceptably low despite intensive multidrug and multi-year therapy. The issue of poor drug exposure relative to inferior potency of repurposed antibiotics against NTM pathogens is broadly recognized as a major factor underlying dismal success rates. Yet, a quantitative and systematic evaluation of probabilities of pharmacokinetic-pharmacodynamic (PK-PD) target attainment (PTA) is lacking. Here, we leveraged published population PK models and minimum inhibitory concentration distributions across large patient populations to generate PTAs for standard of care antibiotics used in the treatment of NTM infections. PTAs of tuberculosis drugs were included in our analysis as a baseline since tuberculosis cure rates are excellent compared to NTM diseases. We found that PK-PD target attainment for NTM was low overall and that PTAs were generally consistent with reported clinical outcomes. Specifically, drugs for which genetic resistance is associated with worse outcome showed favorable PTAs, consistent with their contribution to cure. These include macrolides for *M. abscessus* and the *M. avium* complex (MAC), and rifampicin for *M. kansasii*. Bedaquiline emerged as a promising option against *M. abscessus* and MAC, and clofazimine against the latter, consistent with recent clinical reports. Omadacycline PTAs predict improved efficacy compared to tigecycline for patients with *M. abscessus* and suggest clinical utility for MAC. As is common practice for antibacterials, PTA considerations could inform NTM treatment guidelines and help select drug combinations in future trials.

## INTRODUCTION

Antibiotic treatment of nontuberculous mycobacterial (NTM) pulmonary disease (PD) is long, intensive, and poorly tolerated. Cure rates remain unacceptably low for a group of infections that have been ascribed to culturable pathogens for more than seven decades (1, 2). Two major pathogens account for 85-90% of the lung disease burden caused by NTMs: the *Mycobacterium avium* complex (MAC) and *M. abscessus* (Mab) (3). Standard of care (SOC) largely relies on unoptimized antibiotics repurposed from other disease indications, including tuberculosis (TB). The multidrug and often multiyear treatment of Mab-PD and MAC-PD is overall longer with worse side effects than a typical multidrug resistant TB regimen (4). Despite this intense treatment schedule, a large meta-analysis revealed 46% successful Mab-PD treatment (5, 6), defined as sustained sputum culture conversion for ≥ 12 months while on therapy, or sustained culture conversion without microbiological relapse until the end of therapy. Treatment success is worse for Mab-PD caused by subsp *abscessus,* which is generally macrolide resistant, than for macrolide susceptible subsp *massiliense*: 27-34% versus 54-57% treatment success were reported, respectively, across multiple studies (6, 7)). Likewise, pre-existing macrolide resistance is associated with worse treatment outcome for MAC-PD. Success with the macrolide-rifampicin-ethambutol SOC varies across studies (8). A systematic review of 17 datasets in > 9000 MAC-PD patients revealed high outcome heterogeneity and an overall 5-year mortality rate of 27% (8). Evaluating the respective contribution of the three SOC agents has proven elusive given the diverse array of multidrug regimens (9). Natural history and retrospective studies have revealed up to 52% spontaneous sputum conversion in patients with stable MAC-PD (10, 11), as well as similar disease progression and mortality in MAC-PD patients whether treated or not (12), challenging the contribution of chemotherapy to clinical outcome.

Several known pharmacological factors join forces to limit antibiotic performance. Current NTM treatments include agents that were repurposed from TB, Gram-positive and Gram-negative indications with limited consideration of their markedly lower potency against NTM pathogens. For instance, rifampicin (RIF) is a pillar of TB, *M. kansasii* (Mka)-PD and MAC-PD treatment. However, its MIC (minimum inhibitory concentration) distributions center around 0.06 mg/L against *M. tuberculosis* (Mtb) or Mka (13), but 2 to 8 mg/L for MAC (14), well above the clinical breakpoint of 0.5 mg/L firmly established for TB (15). Applying standard pharmacokinetic-pharmacodynamic (PK-PD) concepts thus precludes a role for RIF in MAC-PD treatment (16). Consistent with and expanding on these findings, the rifampicin-azithromycin-ethambutol SOC did not reduce MAC bacterial burden and only partially prevented growth in the hollow fiber system (HFS) (17). In contrast, rifamycin-based regimens deliver acceptable cure rates for Mka*-* PD (3), in line with Mka RIF MICs being close to Mtb MICs.

Unlike most bacterial infections, NTM treatment is poorly supported by evidence-based quantitative PK-PD metrics, in part due to a lack of antibiotics that meet basic PK-PD targets. Epidemiological cutoffs (ECOFF) have been thoroughly inferred from MIC distributions (18). They are generally higher than the NTM breakpoints suggested by CLSI, which are not strongly supported by clinical efficacy data. ECOFFs are also much higher than the PK-PD based breakpoints defined by EUCAST (18). As a first step towards the rational and quantitative prioritization of promising drug candidates and drug regimens, we have systematically assessed probabilities of target attainment (PTA), which have not been calculated in a comprehensive manner for NTM antibiotics. While it is generally recognized that current NTM treatments are suboptimal because many drugs have been repurposed from TB without adequate dose optimization, here we provide the first systematic quantitative assessment of how much they fall short of available PK-PD targets. We posit that these PTAs generated side-by-side using a standardized approach and combined with geographically diverse MIC distributions will help prioritize novel agents and regimens with best potential for clinical utility, refine doses and dosing frequency, and highlight key gaps in NTM-specific PK-PD targets.

## RESULTS

To generate PTAs, we compiled and integrated published population PK models and PD metrics for 12 antibiotics primarily used to treat MAC- and Mab-PD. Omadacycline (OMC) was added as a newly approved oral tetracycline that completed the first successful randomized placebo-controlled clinical trial in Mab-PD patients, demonstrating high clinical success and culture improvement rates, including macrolide-resistant cases (19, 20). TB and pulmonary infection caused by Mka were included in our analyses as reference disease indications with acceptable cure rates compared to MAC- and Mab-PD. For each pathogen, MIC distributions from different patient populations and publications were retrieved and compiled to minimize biases (**Supplementary Dataset 1**). The resulting aggregated distributions are strongly aligned with a comprehensive study reporting MICs for 1686 MAC isolates and 1014 MAB isolates from 12 European laboratories (18). For the selection of appropriate PK-PD targets, we adopted the following rationale (detailed explanations for each agent are provided in the Methods section). When available, plasma PK-PD targets established for TB were used since (i) these drugs are included in WHO treatment recommendations, enabling extensive clinical analyses of PK-PD versus outcomes across large patient populations and evidence-based PTAs, (ii) the site of disease and the immunopathology of TB and NTM-PD are similar, leading to comparable PK at the site of infection and (iii) the presence of drug tolerant bacterial populations in lung lesions is a hallmark of mycobacterial infections in general. When available, NTM PK-PD targets – most often generated in the HFS (21, 22) – were also considered. For broad spectrum antibiotics used against Gram-positive and/or Gram-negative bacteria, widely accepted PK-PD targets, as well as targets established in the HFS, were used in the absence of mycobacterium-specific targets from clinical studies (**Table 1**). For clarithromycin (CLR), a broadly accepted epithelium lining fluid (ELF) PK-PD target was used as established and validated for lung infections.

**Table 1:**
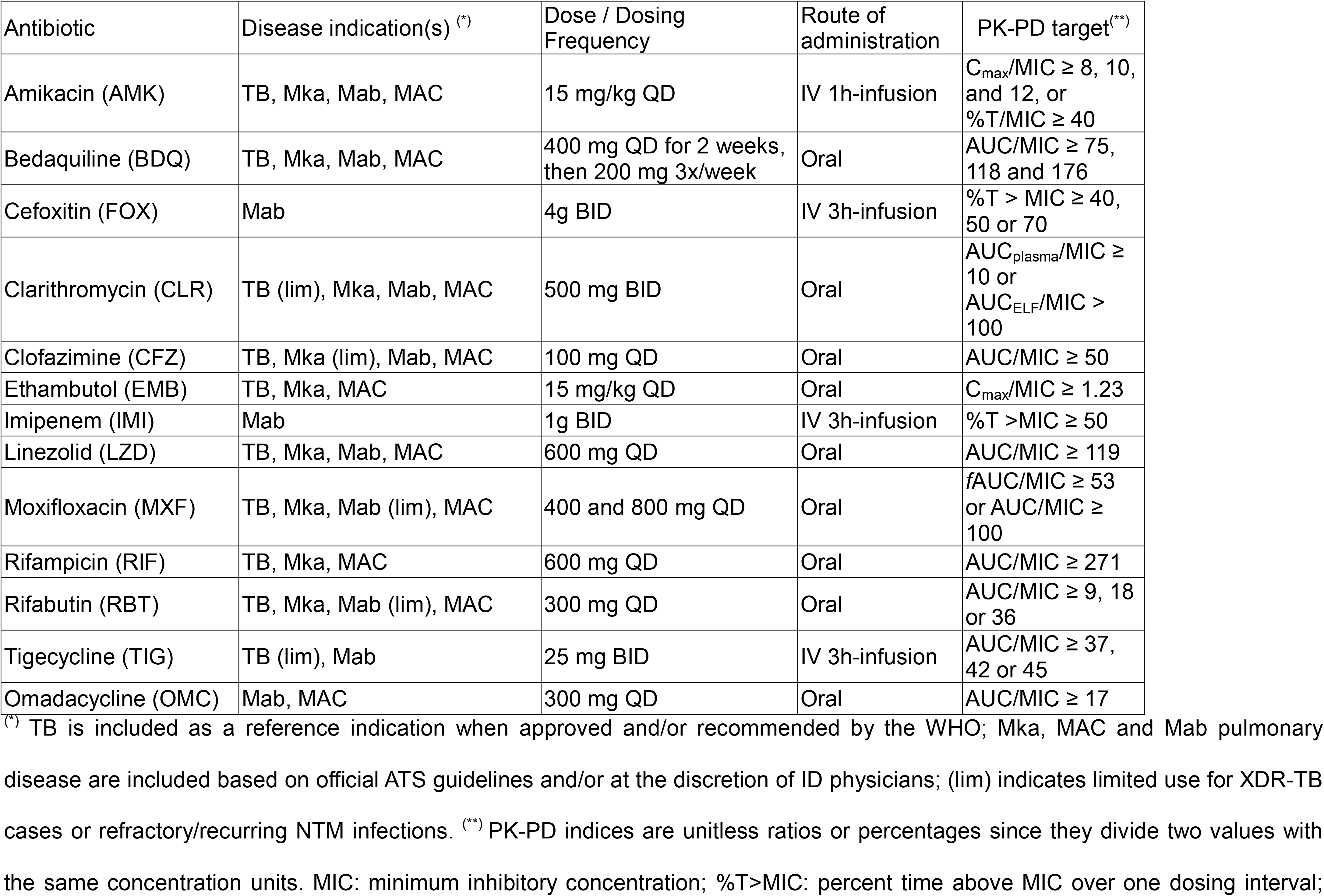
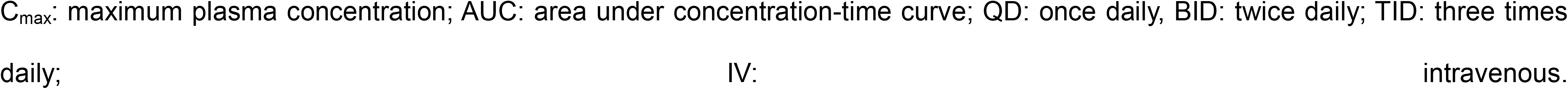
Antibiotic dosing schedules and PK-PD targets.

| Antibiotic | Disease indication(s) <sup>(*)</sup> | Dose / Dosing Frequency | Route of administration | PK-PD target<sup>(**)</sup> |
| --- | --- | --- | --- | --- |
| Amikacin (AMK) | TB, Mka, Mab, MAC | 15 mg/kg QD | IV 1h-infusion | $C_{max}/MIC \geq 8, 10,$<br>and 12, or<br>$\%T/MIC \geq 40$ |
| Bedaquiline (BDQ) | TB, Mka, Mab, MAC | 400 mg QD for 2 weeks,<br>then 200 mg 3x/week | Oral | $AUC/MIC \geq 75,$<br>118 and 176 |
| Cefoxitin (FOX) | Mab | 4g BID | IV 3h-infusion | $\%T > MIC \geq 40,$<br>50 or 70 |
| Clarithromycin (CLR) | TB (lim), Mka, Mab, MAC | 500 mg BID | Oral | $AUC_{plasma}/MIC \geq 10$ or<br>$AUC_{ELF}/MIC > 100$ |
| Clofazimine (CFZ) | TB, Mka (lim), Mab, MAC | 100 mg QD | Oral | $AUC/MIC \geq 50$ |
| Ethambutol (EMB) | TB, Mka, MAC | 15 mg/kg QD | Oral | $C_{max}/MIC \geq 1.23$ |
| Imipenem (IMI) | Mab | 1g BID | IV 3h-infusion | $\%T > MIC \geq 50$ |
| Linezolid (LZD) | TB, Mka, Mab, MAC | 600 mg QD | Oral | $AUC/MIC \geq 119$ |
| Moxifloxacin (MXF) | TB, Mka, Mab (lim), MAC | 400 and 800 mg QD | Oral | $fAUC/MIC \geq 53$<br>or $AUC/MIC \geq 100$ |
| Rifampicin (RIF) | TB, Mka, MAC | 600 mg QD | Oral | $AUC/MIC \geq 271$ |
| Rifabutin (RBT) | TB, Mka, Mab (lim), MAC | 300 mg QD | Oral | $AUC/MIC \geq 9, 18$<br>or 36 |
| Tigecycline (TIG) | TB (lim), Mab | 25 mg BID | IV 3h-infusion | $AUC/MIC \geq 37,$<br>42 or 45 |
| Omadacycline (OMC) | Mab, MAC | 300 mg QD | Oral | $AUC/MIC \geq 17$ |

<sup>(\*)</sup> TB is included as a reference indication when approved and/or recommended by the WHO; Mka, MAC and Mab pulmonary disease are included based on official ATS guidelines and/or at the discretion of ID physicians; (lim) indicates limited use for XDR-TB cases or refractory/recurring NTM infections. <sup>(\*\*)</sup> PK-PD indices are unitless ratios or percentages since they divide two values with the same concentration units. MIC: minimum inhibitory concentration; $\%T > MIC$ : percent time above MIC over one dosing interval;
$C_{\max}$ : maximum plasma concentration; AUC: area under concentration-time curve; QD: once daily, BID: twice daily, TID: three times daily; IV: intravenous.

We first focused on RIF, the pillar of TB and Mka-PD treatment, also included in the 3-drug standard of care of MAC infections. To achieve at least 40% PTA, the RIF MIC should be ≤ 0.125 mg/L and ∼60-65% of Mtb and Mka clinical isolates meet that criterion. At the mode Mtb MIC of 0.062 mg/L, the PTA was > 80%. In contrast, 0.4% of MAC isolates have an MIC ≤ 0.125 mg/L and the PTA was 0% at the mode MICs of RIF (4 to 8 mg/L) (**Figure 1A**). In contrast, we next profiled imipenem (IMI), an injectable β-lactam only reserved for use during the intensive treatment phase of Mab-PD. To achieve 80% PTA, the IMI MIC should be ≤ 4 mg/L and ∼10% of Mab clinical isolates meet that criterion (**Figure 1B**).

**Figure 1.**
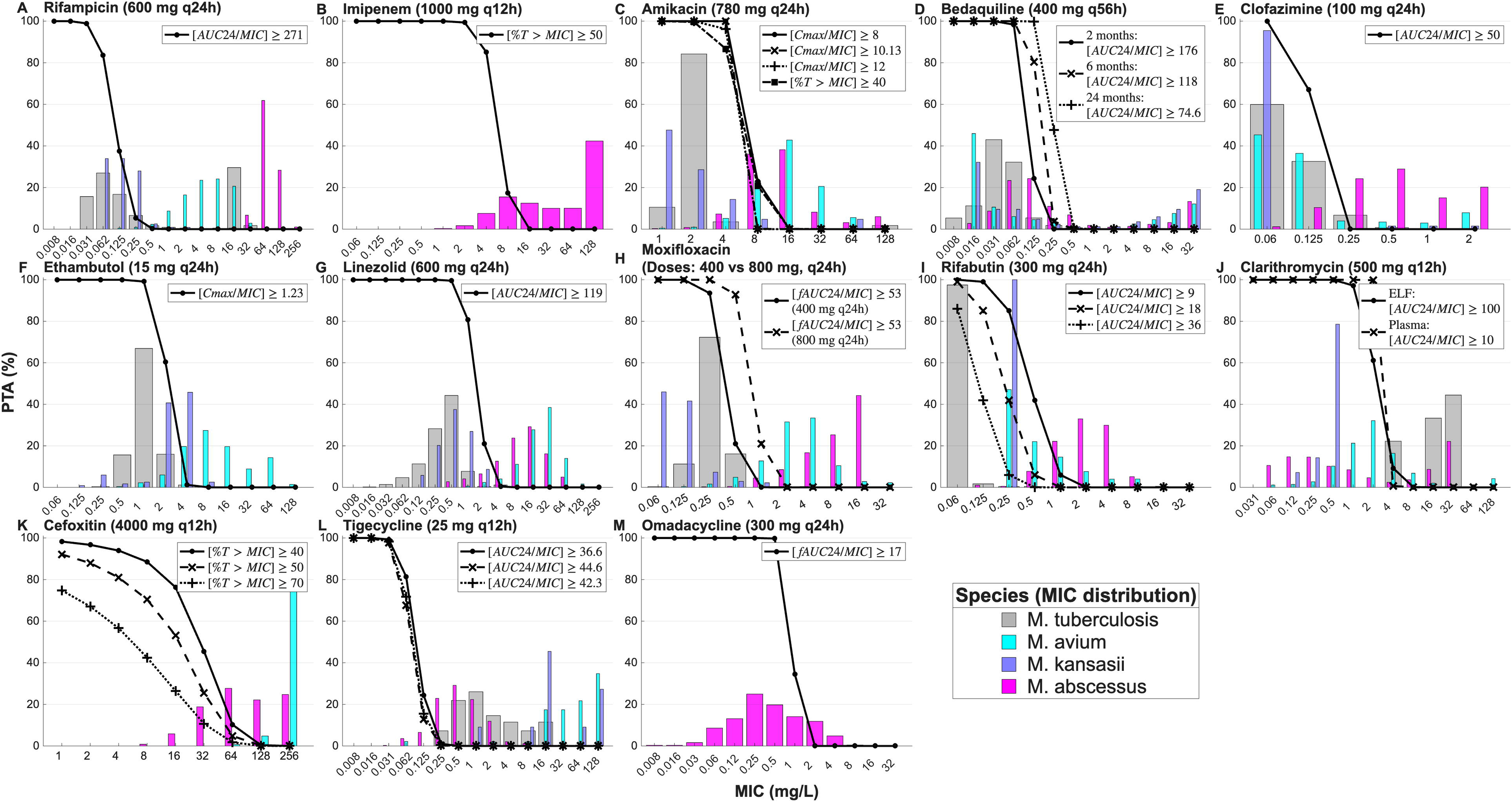
Probabilities of PK-PD target attainment (PTA) for 13 antibiotics used to treat lung infections caused by four mycobacterial pathogens: Mtb, Mka, MAC and Mab. Monte Carlo simulations (n = 10,000 patients) were generated using published population PK models and steady state parameters. PK-PD targets are indicated within each PTA plot with justifications provided in the Methods. Doses, routes and frequency are those recommended for the treatment of mycobacterial infections as follows. Amikacin: 1-hour intravenous infusion of 15 mg/kg once daily (QD); bedaquiline: 400 mg daily for 2 weeks followed by 200 mg three times weekly; clofazimine: 100 mg QD; clarithromycin: 500 mg twice daily (BID); ethambutol: 15 mg/kg QD; cefoxitin: 3-hour intravenous infusion of 4 grams BID; imipenem: 3-hour intravenous infusion of 1 gram BID; linezolid: 600 mg QD; moxifloxacin: 400 and 800 mg QD; rifabutin: 300 mg QD; rifampicin: 600 mg QD; tigecycline: 1-hour intravenous infusion of 25 mg BID; omadacycline: 300 mg QD.

In general, PTAs for Mtb were high (PTA > 80% at least up to the mode MIC) at the approved doses and dosing regimens of antibiotics routinely used against TB, consistent with the high cure rates observed when WHO-recommended regimens are used (**Figure 1C-I**). This mostly held true for drugs used against Mka infections, known to have the highest cure rates among NTM infections (3). However, among the three SOC agents (RIF/RBT, CLR, and EMB) used in Mka-PD treatment, EMB achieved PTA of 60% or greater only for the lower end of the Mka MIC distribution (≤2 mg/L; i.e. ∼52% of Mka isolates) while RIF showed PTA of 60% or greater up to MIC ∼0.1 mg/L (i.e. 34% of Mka isolates). CLR using epithelium lining fluid (ELF) exposure achieved > 90% PTA across the entire MIC range for Mka (**Figure 1J**).

For MAC, CLR (using ELF PK), CFZ and BDQ appeared to achieve acceptable PTA in the susceptible range of their MIC distributions: > 90% PTA up to the mode MIC for CLR (**Figure 1J**), > 60% PTA against ∼80% of the isolates for CFZ (**Figure 1E**) and > 80% PTA over the entire susceptible range for BDQ (**Figure 1D**). BDQ displayed a bimodal MIC distribution with high-level resistance observed in an unexpectedly high fraction of NTM isolates compared to TB. The PTAs of EMB and RIF for MAC-PD were indicative of poor performance at the clinically used doses. For RBT, we found up to 80% PTA at MAC MICs ≤ 0.25 mg/L (∼47% of MAC isolates). This suggests that drug susceptibility testing (DST) may warrant the use of RBT in a subset of MAC-PD patients only (**Figure 1I**), although there remains some uncertainty in the appropriate PK-PD targets for RBT. The PTA of AMK, recommended as an add-on in cases of cavitary, severe or refractory MAC-PD, dropped below 20% at MICs > 4 mg/L, which includes 91% of MAC isolates (**Figure 1C**). These results are expected if placed in the context of clinical breakpoints established for TB and Gram-positive or -negative lung infections (**Table 2**).

**Table 2.** Comparison of clinical breakpoints established for TB and Gram-positive or -negative infections with susceptibility breakpoints adopted for NTM by the CLSI (18).

|  | NTM (CLSI) | Other indications |
| --- | --- | --- |
| Amikacin (AMK) | 16 to 32 mg/L | 2 mg/L for TB |
| Bedaquiline (BDQ) | n.r. | 0.25 mg/L for TB |
| Cefoxitin (FOX) | n.r. | 8 mg/L for Gram-negative |
| Clarithromycin (CLR) | 16 mg/L for MAC-PD<br>4 mg/L for Mab-PD | 0.25 mg/L for *Streptococcus pneumoniae*<br>8 mg/L for *Haemophilus influenza* |
| Clofazimine (CFZ) | n.r. | 0.5 to 1 mg/L for TB |
| Ethambutol (EMB) | n.r. | 5 mg/L for TB |
| Imipenem (IMI) | 8 to 16 mg/L | 1 to 2 mg/L for Gram-negative |
| Linezolid (LZD) | 16 mg/L | 1 to 2 mg/L for TB |
| Moxifloxacin (MXF) | 2 mg/L | 0.25 to 0.5 mg/L for TB |
| Rifampicin (RIF) | 8 mg/L | 0.5 mg/L for TB |
| Rifabutin (RBT) | n.r. | 0.125 mg/L for TB |
| Tigecycline (TIG) | n.r. | 0.25 to 0.5 mg/L for Gram-negative or -positive |
| Omadacycline (OMC) | n.r. | 0.5 mg/L for *S. pneumoniae* <sup>(*)</sup> |

n.r.: no official susceptibility breakpoint established or reported by CLSI or EUCAST.
(\*) FDA endorsed https://www.fda.gov/drugs/development-resources/omadacycline-injection-and-oral-products

For Mab, BDQ but not CFZ showed acceptable PTA, with BDQ achieving >80% PTA for ∼60% of Mab isolates and CFZ reaching 60% PTA for only 14% of Mab isolates. CLR displayed a largely bimodal MIC distribution for Mab due to the widespread resistance mediated by *erm*41 in Mab subsp. *abscessus* (23), resulting in 100% PTA in the susceptible portion of the distribution representing mostly subsp. *massiliense* (**Figure 1J**).

During the intensive phase of Mab-PD therapy, one to three injectables, depending on macrolide susceptibility, are selected among AMK, FOX, IMI and TIG, for a duration of 4 to 12 weeks, at the clinician’s discretion. Due to the long duration of this intensive phase, these agents are administered at lower doses and less often than recommended when treating Gram-negative or Gram-positive infections for 1-2 weeks. For example, the recommended dosing schedule for IMI and FOX is a 3h-infusion TID but this is reduced to BID in Mab-PD patients, and doses are selected within the lower end of the approved range particularly in elderly patients. Likewise, the most common TIG intravenous dose for Mab-PD is 25 mg BID due to gastrointestinal and liver toxicity, whereas 50 mg BID is used for pneumonia after a 100 mg loading dose. Four-gram BID infusions of FOX and 1-gram BID infusions of IMI achieved PTAs indicative of limited clinical utility (> 20%) only at the lower end of the MIC distribution (**Figure 1B, C, K, L**). The PTAs improved when we simulated TID dosing for IMI and FOX, and 50 mg BID for TIG, but only to a limited extent (**Supplementary Figure 1**). This suggests that TID β-lactam infusions could be tested in clinical research settings for their potential to accelerate sputum conversion in patients with MICs indicative of acceptable PTA (≤ 32 mg/L for FOX and ≤8 mg/L for IMI). Compared to TIG, OMC emerged as a promising oral and well tolerated tetracycline with 100% PTA for > 70% of the Mab isolates (**Figure 1M**).

LZD and MXF, both oral agents included in the Mab-PD treatment guidelines (24), had poor PTA (at or close to 0%, **Figure 1J-H**) over almost the entire MIC ranges of Mab and MAC, not surprisingly when considering the clinical breakpoints established for MDR-TB (**Table 2**). RBT was considered as an emerging repurposed antibiotic against Mab-PD and an alternative to RIF against MAC infections. The Mab PTAs do not suggest clinical utility for RBT (**Figure 1I**).

Overall, the PTAs of the antibiotics surveyed in this study were remarkably poor for MAC and Mab compared to Mtb and Mka **(Figure 2**), in line with the long treatment duration, poor cure rates and frequent recurrence. Nonetheless, our results identify opportunities where focused DST or dosing adjustments may impact treatment outcomes.

**Figure 2.**
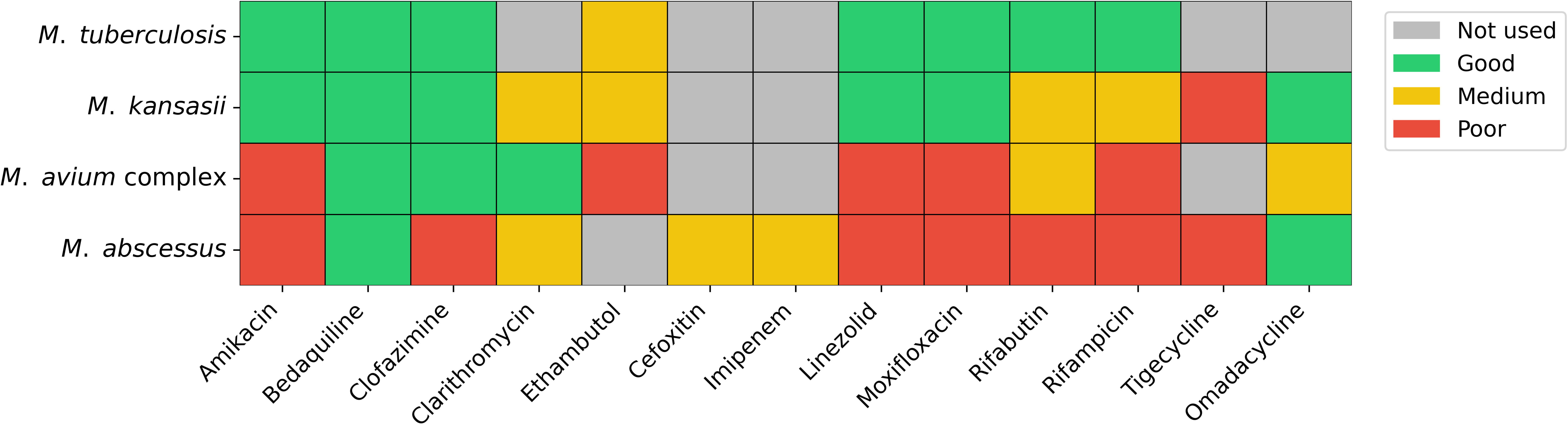
Qualitative expected clinical utility by drug and pathogen, based on PTA.

## DISCUSSION

To place the generally poor cure rates of NTM-PD in the context of PK-PD target attainment, we generated PTAs for 12 antibiotics included in their treatment guidelines, 8 of which are included in the treatment of drug susceptible, MDR and/or XDR TB, along with OMC. Consistent with the superior TB cure rates compared to NTM-PD, we found PTAs ≥ 90% at least up to the modal Mtb MIC for the 8 antibiotics used to treat TB. PTAs were also generally positive for Mka, for which treatment with a rifamycin-based regimen is usually successful (3). In contrast, most PTAs were poor over a substantial portion of the MIC range for MAC and Mab. The macrolide CLR was the notable exception for Mka, Mab and MAC, with acceptable PTA over the susceptible MIC range, consistent with worse treatment outcomes for widespread macrolide-resistant versus susceptible NTM-PD (6, 25, 26). Azithromycin (AZI) is more commonly used than CLR to treat NTM-PD but DST almost exclusively relies on CLR as a surrogate of AZI (3) due to historical data availability, stability and reliability. We did not calculate PTAs for AZI given the paucity of MIC distributions, but very low PTA was determined in a Mab-HFS study (27). Our results also indicate that CFZ and BDQ for MAC-PD, as well as BDQ for Mab-PD, deserve further clinical investigations with careful considerations of partner drugs since they synergize with – but may also antagonize – other antibiotic classes (28, 29).

Against MAC, two of the three SOC agents (rifamycin-EMB) displayed remarkably poor PTAs. consistent with a recent meta-analysis showing that omitting the rifamycin was non-inferior to the 3-drug regimen in MAC-PD patients (30). The poor PTAs partially explain the significant rates of acquired macrolide resistance while on treatment with those three drugs, in patients and in the HFS (26, 31), in addition to RIF decreasing CLR exposure via induction of CYP3A4 (32). The clinical utility of RIF against MAC-PD has been increasingly questioned (16), in line with the lack of correlation between RIF MIC and treatment outcome (9) and the disconnect between TB breakpoints and MAC MIC distributions outside of clinically achievable concentration ranges. In a landmark HFS study, the RIF-AZI-EMB SOC did not achieve stasis when plasma PK was simulated against intracellular and extracellular *M. avium.* Simulating ELF PK achieved stasis for 3 days followed by growth, a slightly better performance than when plasma PK was reproduced (17). Replacing RIF with CFZ in the SOC of MAC-PD delivered improved efficacy in the HFS (33). Macrolide use is recommended against macrolide resistant cases owing to their beneficial immunomodulatory effect (3), further complicating assessment of their contribution to microbiological outcome and disease progression.

The overall poor PTA of the SOC used to treat MAC-PD is consistent with the underreported and often ignored natural history of MAC-PD. A recent – albeit small – retrospective study showed no difference in mortality and disease progression between NTM patients who were treated and not treated, the majority of which had sputum positive MAC-PD (12). Similarly, studies of untreated MAC-PD patients revealed up to 40 to 50% spontaneous sputum conversion depending on disease severity (10, 11), not very different from the average 54– 65% of patients achieving sustained culture conversion in a systematic review of pulmonary MAC treatment (34). These observations call into question the contribution of SOC chemotherapy to MAC-PD clinical outcome, in line with our PTA profiles.

The poor performance of MAC and Mab treatments is compounded by the fact that doses most often prescribed by clinicians are at the lower end of the approved range due to moderate to severe side effects upon long treatment duration. In addition, many patients are affected by comorbidities and/or elderly, further limiting the tolerability of chemotherapeutics. Our dosing simulations of the injectables (FOX, IMI and TIG) at the higher end of the approved range/frequency, known to be required for effective treatment of Gram-negative and Gram-positive infections despite lower MIC distributions, showed improved PTA. This suggests that higher doses could be beneficial if well-tolerated by patients whose drug susceptibility profiles justify it. Likewise, CFZ is generally dosed at 100 mg daily to MAC-PD and TB patients while it is approved up to 200 mg daily. Literature review indicates that the 100 mg dose is driven by efficacy against leprosy, as well as economical and perceived side effect considerations (35). As suggested, a loading phase and higher dose may both improve efficacy and shorten treatment duration.

Most of the 8 drugs used against TB and NTM-PD display dramatically left-shifted MIC distributions against Mtb compared to MAC or Mab. Modal MAC/Mab MICs are 4 to 64-fold higher than their Mtb counterparts, calling into question the use of these agents to treat NTM-PD. While evidence-based critical concentrations and clinical breakpoints have been established for TB drugs, limited consensus exists in the treatment of NTM-PD (18). Notably, the WHO TB breakpoints for AMK, MXF and LZD are 2, 0.5 and 1 mg/L (15), respectively. In contrast, the CLSI NTM breakpoints of these three agents are 32, 2 and 16 mg/L, and EUCAST’s PK-PD based breakpoints are 1, 0.25 and 2 mg/L (18). In addition, MIC distributions are markedly wider for NTMs than Mtb due to the higher genetic diversity of NTMs.

AMK is a poorly tolerated injectable with serious oto- and nephrotoxicity. PTAs were poor at the recommended dose of 15 mg/kg, consistent with published PK-PD studies (36) and with recent Mab HFS data showing that T>MIC may be the PK-PD driver and that the proposed target is not achievable in patients at 15 mg/kg (37). AMK liposome inhalation suspension (ALIS) is better tolerated and has shown promising results in small cohorts of patients with refractory disease. The PTA of ALIS at the site of disease would be very informative to optimize its dose and use, but pulmonary PK data have not been published to our knowledge.

Likewise, TIG is a poorly tolerated injectable with serious gastrointestinal intolerance. Yet, it has been suggested that 100 mg BID instead of the approved 100 mg daily is needed for effective treatment of Gram-negative and Gram-positive community-acquired pneumonia. In contrast, the Mab-PD recommended dose is 25 mg BID and Mab MICs are markedly higher. OMC, a new orally bioavailable tetracycline, recently completed the first promising clinical trial in Mab-PD patients (19, 20) and could be considered as a substitute for TIG, based on much improved PTA, HFS results (21) and tolerability.

This work comes with the following limitations. First, PK-PD targets specific to NTM pathogens based on clinical evidence are not yet available. Thus, we have used different sources of PK-PD targets with priority given to those based on clinical treatment outcomes of pulmonary infections. While we provide a rationale for each drug, we acknowledge that the approach is suboptimal, highlighting the unmet need for NTM-specific PK-PD targets and MIC breakpoints. Our PTA set relies on PK-PD targets that use plasma PK (except for CLR) and classic MICs obtained in broth against replicating cultures. We propose that the dataset constitutes a harmonized starting point that can be expanded with site-of-disease PK (38) and potency values against specialized bacterial populations found in immune cells and cavity caseum (39). MICs of single drugs were used whereas combinations may cause synergy or antagonism that could modulate MIC distributions and therefore PTAs of a drug regimen. Second, our analysis excludes non-pulmonary diseases such as skin and surgical site infections, meningitis, or prosthetic infections, and excludes mycobacterial pathogens other than Mtb, Mka, MAC and Mab. Finally, peer reviewed and data rich population PK models were selected, re-implemented and evaluated against the source publications. PK models obtained with specific patient populations would constitute a useful expansion of the present study.

Collectively, our results do not come as a surprise given the large shift in MIC distributions between the pathogens against which doses were optimized and MAC/Mab, while the doses used in long-term NTM-PD treatment are either identical or lower. NTM-PD guidelines recommend multidrug regimens, but clinical success is primarily driven by RIF for Mka- and macrolides for Mab- and MAC-PD. Resistance to these agents is associated with treatment failure, because companion drugs lack independent bactericidal activity to achieve sterilization and fail to protect the few efficacious drugs against emergence of resistance. The management of NTM-PD remains a challenge characterized by a high reliance on a very limited number of effective antibiotics. Yet, comprehensive DST of patient isolates prior to treatment initiation could be an invaluable resource to design patient-tailored regimens with optimal PTAs even if far from ideal.

Our study reveals ample room for improvement and indicates that therapeutic and drug discovery efforts with a strategic focus on PK-PD and evidence-based breakpoints could improve cure rates and even shorten treatment duration.

## METHODS

Antibiotics were selected based on ATS guidelines for the treatment of NTM-PD (40) and key metrics for each are listed in **Table 1**.

### Pharmacokinetic models

For each antibiotic, a population PK model was selected based on consistency with other literature, relevance of target population used for data collection, and model reproducibility. PK model equations and diagrams are presented for each antibiotic in **Supplementary Dataset 2**. PK models were implemented in Matlab, and code is available at https://github.itap.purdue.edu/ElsjePienaarGroup/NTM_PTAs_Public.

Each PK model was simulated for 10,000 virtual patients. Visual comparisons between pharmacokinetic time courses of our simulations and the source papers were used to assess successful implementation (e.g. peak concentrations, half-life, etc.). Model-predicted AUC values were also compared to typical ranges (**Table S1** in **Supplementary Dataset 2**).

### MIC distributions

MIC distributions of each antibiotic against the relevant pathogens were compiled from the literature, ensuring that several studies and patient populations were included for each pathogen (**Supplementary Dataset 1**). For MAC, the two major species, *M. avium* and *M. intracellulare*, were included and aggregated, based on overall similar profiles when averaging large and diverse patient populations (41). For Mab, MIC distributions against the three subspecies Mab *abscessus, massiliense* and *bolletii* were included and aggregated.

### Justification of PK-PD target selection

PK-PD targets supported by clinical outcomes are largely missing for NTM infections. When available, we used PK-PD targets established for TB based on clinical outcome rather than PK-PD targets determined in the HFS since the site of infection, host factors and physiological state of the pathogen all have a marked impact on PK-PD and are not recapitulated in the HFS. We acknowledge the limitations of using different sources of PK-PD targets for non-TB drugs, which further underscores the dire need for NTM-specific PK-PD targets grounded in clinical data.

#### Amikacin (AMK)

No clinical PK-PD targets specific to mycobacterial infections were available in the literature. We found two HFS studies that used Mab (37, 42) and reported different PK-PD targets as follows: (1) C_max_/MIC > 3.2 to achieve 80% Emax in the Mab HFS where serum PK profiles of increasing AMK doses were simulated. Only stasis was achieved even at very high C_max_/MIC. AMK was thus poorly active in the Mab HFS. The authors also hint at %T>MIC potentially being a better driver of efficacy than C_max_/MIC but did not pursue conclusive analyses to confirm this hypothesis. (2) Consistent with the hypothesis raised by the previous authors, 40% T> MIC was found to achieve EC_80_ for growth inhibition, acknowledging that 40% T> MIC is almost unachievable in patients as currently administered. The authors also showed that using a PK-PD target of C_max_/MIC > 3.2 vastly overestimates PTA of 80% E_max_ at human doses (36). We also found an *M. tuberculosis* (Mtb) HFS study where C_max_/MIC > 10 achieved 90% E_max_ (43) where E_max_ is ∼ 5 log CFU kill.

Given the discrepant PTAs and variable outcomes that emerged from these HFS studies, we included C_max_/MIC > 10 and 40% T>MIC as the PK-PD targets for AMK and added C_max_/MIC of ≥ 8 or ≥ 12 as these are broadly adopted targets for Gram-positive and Gram-negative infections (44, 45).

#### Bedaquiline (BDQ)

We relied on a compelling clinical TB study where distinct PK-PD targets were established to achieve three decreased orders of stringency: 2-month sputum culture conversion, 6-month sputum culture conversion and overall positive outcome (46), as indicated.

#### Cefoxitin (FOX)

%T>MIC is the driver of efficacy for β-lactams and 40 to 70% T>MIC is standard for cephalosporins across disease indications. C_ss_>MIC has also been suggested for the treatment of Mab-PD (47) which would require continuous infusion with significant tolerability and practical issues. Thus, we calculated PTAs at 40, 50 and 70% T>MIC, for a dosing regimen of 3-hour 4-gram infusions twice daily, standard for *Mab* patients. To treat other infections that require short term therapy, FOX is commonly administered by infusion 3 to 4 times daily. This is not practical given the length of the intensive phase in the treatment of Mab infections. Nevertheless, we show the PTAs for three times 3-hour 4-gram daily infusions in Supplementary Figure 1 to quantify the potential improvement.

#### Clarithromycin (CLR)

CLR is a broad-spectrum antibiotic with a long history of usage against Gram-positive and Gram-negative infections. AUC_ELF_/MIC > 100 has been established as a PK-PD target to achieve 90% PTA for lung infections (48). Since ELF/plasma ratios of approximately 10 were measured across several pulmonary disposition studies (49), we used both AUC_plasma_/MIC > 10 and AUC_ELF_/MIC > 100.

#### Clofazimine (CFZ)

In a study with 105 patients, AUC/MIC > 50.5 for CFZ in combination treatment was associated with faster sputum clearance in MDR-TB (50). We used this metric in the absence of additional evidence-based PK-PD targets for CFZ. It synergizes with several antibiotics in vitro, and therefore its PK-PD target may vary depending on the background regimen.

#### Ethambutol (EMB)

Although several PK-PD targets have been reported for EMB (51–53), we found that only C_max_/MIC > 1.23 is supported by published data. This value was established in a MAC hollow fiber system study, to achieve 90% of effective concentration in serum (53).

#### Imipenem (IMI)

We used the broadly established 50% T>MIC across infectious diseases and simulated a dosing regimen of 3-hour 1-gram infusions twice daily, standard for *Mab* patients. Similar to FOX, IMI is commonly administered by infusion 3 times daily to treat other infections that require short term therapy. Thus, we show the PTAs for three times 3-hour 1-gram daily infusions in Supplementary Figure 1.

#### Linezolid (LZD)

A broadly accepted PK-PD target is AUC/MIC of 80 to 120 (reviewed in (54) with 7 references to clinical studies) across pathogens and infectious diseases. In the TB HFS, a PK-PD target of AUC/MIC > 119 achieved 80% of maximum kill and was adopted in our PTA calculations (55)

#### Moxifloxacin (MXF)

A broadly accepted PK-PD target against Gram-negative, Gram-positive and mycobacterial infections is AUC/MIC > 100 (45, 56, 57). A free AUC/MIC > 53 has been established in the TB hollow fiber system (58), which aligns with AUC/MIC > 100 given the ∼50% unbound plasma concentrations of MXF. We used the standard daily dose of 400 mg as well as a high dose of 800 mg, which has been suggested based on translational PK-PD models (59, 60), tested in a recent clinical trial (NCT01329250) and prescribed by clinicians to patients with low-level MXF resistance (61).

#### Rifampicin (RIF)

An AUC/MIC ≥ 271 was established in the mouse model of chronic TB as the PK-PD target required to achieve a 1-log CFU reduction in the lungs (62). This target was tested in a population modeling and Monte Carlo simulation study (63, 64). At the standard daily dose of 600 mg, the rates of target attainment (AUC/MIC ≥ 271) ranged from 78 to 100% for MICs between 0.01 and 0.1 mg/L. Their work and that of others generally support the need to evaluate higher doses of RIF for the treatment of TB patients. We used this conservative target to avoid underestimating the PTA of RIF against NTM infections, which is already very poor compared to TB.

#### Rifabutin (RBT)

RBT is generally used instead of RIF to treat TB-HIV patients due to its lower induction of CYP3A4 (which metabolizes many antiretroviral agents). There is no published PK-PD target for RBT against TB. We found an AUC range of 3.8 to 5.2 mg*h/L generally associated with favorable outcome (65, 66). The clinical breakpoint (CB) of RBT is set at 0.5 mg/L by the WHO (67) and some studies suggest 0.25 or 0.125 mg/L (68). Selecting an average AUC of 4.5 mg*h/L and the three CBs of 0.125, 0.25 and 0.5 mg/L returned PK-PD targets of AUC/MIC ≥ 9, 18 and 36, consistent with (69) where AUC_[0-24]_/MIC = 0.05 mg/L *24h/0.064 mg/L = ∼ 20

#### Tigecycline (TIG)

In the Mtb HFS, an AUC/MIC of 42 was associated with maximum kill (70). In the Mab HFS, AUC/MIC of 37 and 45 were found to achieve 80% E_max_ or 1-log kill, respectively. The E_max_ was modest even at very high simulated doses, with rapid rebound. These three values were used as targets in the absence of PK-PD targets based on clinical outcome. In published Monte Carlo simulations, the standard dose of 100 mg/day did not achieve stasis (71), in line with recent studies indicating that 100 mg every 12 h is needed for community-acquired pneumonia (72). During the intensive phase of Mab-PD treatment, however, 25 mg BID is recommended due to dose-limiting gastrointestinal toxicity (73), and was therefore used to generate the TIG PTA in the main text. We also included PTAs at 50 mg BID as a supplementary figure.

#### Omadacycline (OMC)

Given the serious side effects and poor PTA of TIG, we included OMC as a recently approved oral and better tolerated tetracycline that achieved promising results in a recent Phase 2b trial against pulmonary Mab infections (19). In vitro and animal studies have shown that the PK/PD driver is AUC/MIC in models of *S. aureus, S. pneumonia, E. coli* and *A. baumannii* infection. Given the lack of published NTM clinical or preclinical PK-PD studies, we used the following PK-PD targets: (a) *f*AUC/MIC drawn from a publication that focuses on Acute Bacterial Skin and Skin Structure Infection (ABSSSI) and Community-Acquired Bacterial Pneumonia (CABP) (74) and (b) AUC/MIC identified in the TB hollow fiber system (75). The two sets of PK-PD targets were similar: *S. pneumoniae*: median target for 1-log10 reduction for AUC/MIC =17.4; *S. aureus*: median target for stasis for AUC/MIC = 21.9). In the TB HFS, 50% of E_max_ (4.64 log10 reduction in CFU/mL) was achieved at AUC/MIC = 22.9 and in the Mab HFS, an average AUC/MIC of 23 achieved 80% of E_max_ (75). Taking these collective clinical and HFS metrics into account, and prioritizing in vivo over HFS studies, we selected AUC/MIC ≥ 17 at an oral dose of 300 mg QD used in the phase 2 Mab trial.

### Probability of Target Attainment (PTA)

PTAs were calculated using published PK-PD targets (AUC/MIC, % time over MIC (%T>MIC), C_max_/MIC) that were selected according to the rationale described above and are listed in **Table 1**. PTA is computed as percentage of 10,000 simulated patients that achieve the PK-PD target.

## Data Availability

All data produced in the present work are contained in the manuscript and associated supplemental information. Any additional information will be made available upon reasonable request to the authors.

https://github.itap.purdue.edu/ElsjePienaarGroup/NTM_PTAs_Public

## ACKNOWLEDGEMENTS

We thank Drs Chuck Daley and Ken Olivier for discussing antibiotic dosing and frequency in the treatment of NTM-PD patients. We are grateful to Dr Min Xie for critical reading of the manuscript. This work was supported by NIH-NIAID R01-AI184502 and R01-AI132374 to TD and VD, and NIH-NIAID R01-AI172838 to PE and EP.

