## Supplementary figure 1 for "Falling short: Inadequate target attainment in guideline-based therapy of nontuberculous mycobacterial disease"

**Figures**

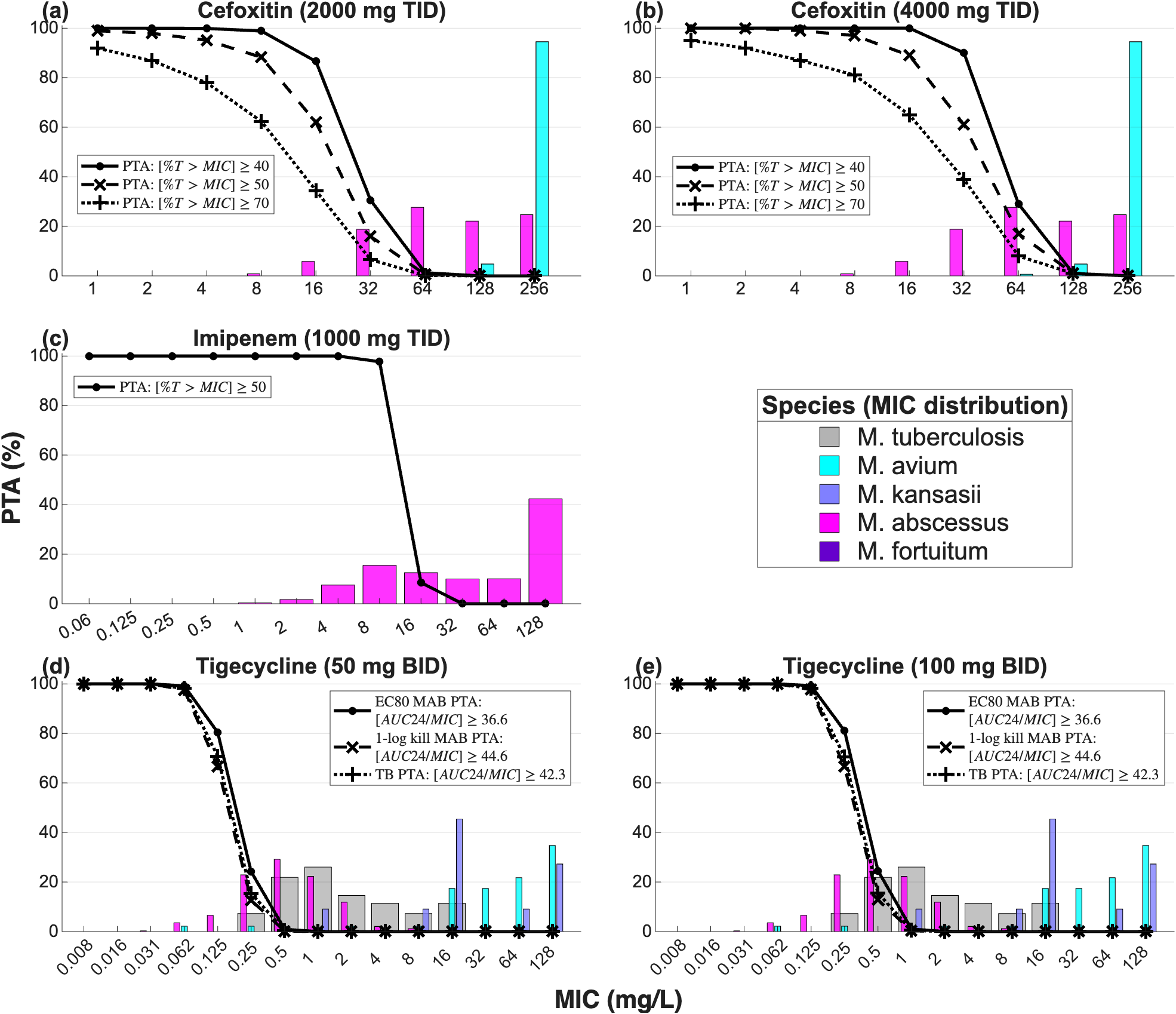

**Supplementary Figure 1:** Probabilities of PK-PD target attainment (PTA) at increased doses or dosing frequency for cefoxitin (FOX), imipenem (IMI) and tigecycline (TIG). Monte Carlo simulations (n = 10,000 patients) were generated using published population PK models and parameters at steady state. PK-PD targets are indicated within each PTA plot. Doses, routes and frequency are those recommended for the treatment of mycobacterial infections as follows. For FOX, 3-hour TID intravenous infusions of 2 grams and 4 grams were simulated. The latter shows moderate improvement of PTA compared to the same dose infused BID. This is consistent with a study suggesting continuous FOX infusion to improve PTA by achieving C_ss_>MIC for the treatment of Mab-PD, with significant tolerability and practical issues [1]. For IMI, 3-hour TID intravenous infusions of 1 gram were simulated, again showing moderate improvement compared to BID. Only patients at the lower end of the MIC distribution would benefit from higher three times daily infusions of FOX or IMI. For TIG, 1-hour BID intravenous infusions of 50 mg and 100 mg were simulated. The PTAs suggest that higher doses, often poorly tolerated, would not be of significant benefit to most patients.
