## Supplementary dataset 2 for "Falling short: Inadequate target attainment in guideline-based therapy of nontuberculous mycobacterial disease"

S5 Simulated steady-state central compartment concentration-time profiles for amikacin showing the mean and 90% prediction interval across 10,000 virtual patients receiving 15 mg/kg q24h. 12

S6 Schematic of the ten-compartment population pharmacokinetic model for bedaquiline, showing the absorption compartment ($q_0$), five transit compartments ($q_1$–$q_5$), final transit compartment ($q$), central compartment ($C$), and two peripheral compartments ($C_2$) and ($C_3$), with systemic clearance (CL) and intercompartmental transfer (Q_2, Q_3). . . . 14

S7 Steady-state $\text{AUC}_{0\text{–}24}$ distributions across 10,000 simulated patient profiles for bedaquiline at 400 mg q24h for two weeks followed by 200 mg three times weekly. Distributions reflect interindividual variability drawn from the parameter distributions in Table S3. . . . 15

S8 Simulated steady-state central compartment concentration-time profiles for bedaquiline across 10,000 virtual patients during the maintenance phase of 200 mg three times weekly (following 400 mg q24h for two weeks). . . . 16

S9 Simulated steady-state central compartment concentration-time profiles for bedaquiline showing the mean and 90% prediction interval across 10,000 virtual patients during the maintenance phase of 200 mg three times weekly (following 400 mg q24h for two weeks). . . . 17

S10 Schematic of the three-compartment population pharmacokinetic model for clofazimine, showing the absorption compartment ($q_0$), central compartment ($C$), and two peripheral compartments ($C_{p1}$) and ($C_{p2}$), with systemic clearance (CL/F) and intercompartmental transfer (Q1/F, Q2/F). . . . 19

S11 Steady-state $\text{AUC}_{0\text{–}24}$ distributions across 10,000 simulated patient profiles for clofazimine at 100 mg q24h. Distributions reflect interindividual variability drawn from the parameter distributions in Table S4. . . . 19

S12 Simulated steady-state central compartment concentration-time profiles for clofazimine across 10,000 virtual patients receiving 100 mg q24h. . . . 20

S13 Simulated steady-state central compartment concentration-time profiles for clofazimine showing the mean and 90% prediction interval across 10,000 virtual patients receiving 100 mg q24h. 21

S14 Schematic of the three-compartment population pharmacokinetic model for clarithromycin, showing the absorption compartment ($q_0$), central compartment ($C$), peripheral compartment ($C_2$), and epithelial compartment ($C_3$), with systemic clearance (CL/F) and intercompartmental transfer rate constants (K_12, K_21, K_13, K_31). . . . 23

S15 Steady-state $\text{AUC}_{0\text{–}24}$ distributions across 10,000 simulated patient profiles for clarithromycin at 500 mg q12h. Distributions reflect interindividual variability drawn from the parameter distributions in Table S5. . . . 23

S16 Simulated steady-state central compartment concentration-time profiles for clarithromycin across 10,000 virtual patients receiving 500 mg q12h. . . . 24

S17 Simulated steady-state epithelial lining fluid concentration-time profiles for clarithromycin across 10,000 virtual patients receiving 500 mg q12h. . . . 25

S18 Simulated steady-state central compartment concentration-time profiles for clarithromycin showing the mean and 90% prediction interval across 10,000 virtual patients receiving 500 mg q12h. 26

S19 Simulated steady-state epithelial lining fluid concentration-time profiles for clarithromycin showing the mean and 90% prediction interval across 10,000 virtual patients receiving 500 mg q12h. 27

S20 Schematic of the two-compartment population pharmacokinetic model for ethambutol with transit-compartment absorption, showing the absorption input ($q$), central compartment ($C$), and peripheral compartment ($C_p$), with systemic clearance (CL_F), intercompartmental clearance (Q_F), and HIV covariate effect on bioavailability (F). . . . . . . . . . . . . . . . . . . . . . 29
S21 Steady-state $AUC_{0-24}$ distributions across 10,000 simulated patient profiles for ethambutol at 15 mg/kg q24h. Distributions reflect interindividual variability drawn from the parameter distributions in Table S6. . . . . . . . . . . . . . . . . . . . . . . . . . . . . . . . . . . . . . 30
S22 Simulated steady-state central compartment concentration-time profiles for ethambutol across 10,000 virtual patients at 15 mg/kg q24h. . . . . . . . . . . . . . . . . . . . . . . . . . . . . . . 31
S23 Simulated steady-state central compartment concentration-time profiles for ethambutol showing the mean and 90% prediction interval across 10,000 virtual patients at 15 mg/kg q24h. . . 32
S24 Schematic of the one-compartment population pharmacokinetic model for cefoxitin, showing the central compartment ($C$) with systemic clearance (CL) and a short IV infusion input. Drug is eliminated from the central compartment by a first-order process. . . . . . . . . . . . 34
S25 Steady-state $AUC_{0-24}$ distribution across the simulated virtual-patient population for cefoxitin at 4 g q12h administered as a 3-hour IV infusion. The distribution reflects interindividual variability in V_c and CL drawn from the parameters in Table S7. . . . . . . . . . . . . . . . . 35
S26 Simulated steady-state central-compartment concentration-time profiles for cefoxitin across the virtual-patient population receiving 4 g q12h as a 3-hour IV infusion. . . . . . . . . . . . 36
S27 Simulated steady-state central-compartment concentration-time profiles for cefoxitin showing the median (50th percentile) and the 5th–95th percentile (90%) prediction interval across the virtual-patient population receiving 4 g q12h as a 3-hour IV infusion. . . . . . . . . . . . . . . 37
S28 Schematic of the two-compartment population pharmacokinetic model for imipenem, showing the central compartment ($C$) and peripheral compartment ($C_p$), with systemic clearance (CL_F), intercompartmental clearance (Q_F), and 3-hour IV infusion input. . . . . . . . . . . 38
S29 Steady-state $AUC_{0-24}$ distributions across 10,000 simulated patient profiles for imipenem at 1000 mg q12h administered as a 3-hour IV infusion. Distributions reflect interindividual variability drawn from the parameter distributions in Table S8. . . . . . . . . . . . . . . . . . . 39
S30 Simulated steady-state central compartment concentration-time profiles for imipenem across 10,000 virtual patients receiving 1000 mg q12h as a 3-hour IV infusion. . . . . . . . . . . . . . 40
S31 Simulated steady-state central compartment concentration-time profiles for imipenem showing the mean and 90% prediction interval across 10,000 virtual patients receiving 1000 mg q12h as a 3-hour IV infusion. . . . . . . . . . . . . . . . . . . . . . . . . . . . . . . . . . . . . . . . . 41
S32 Schematic of the eight-compartment population pharmacokinetic model for linezolid, showing the absorption compartment ($q_0$), five transit compartments ($q_1$–$q_5$), final pre-absorption compartment ($q$), and central compartment ($C$), with absorption rate constant (K_a), transit rate constant (k_tr), bioavailability (F), and systemic clearance (CL). . . . . . . . . . . . . . 43
S33 Steady-state $AUC_{0-24}$ distributions across 10,000 simulated patient profiles for linezolid at 600 mg q24h. Distributions reflect interindividual variability drawn from the parameter distributions in Table S9. . . . . . . . . . . . . . . . . . . . . . . . . . . . . . . . . . . . . . . . 44
S34 Simulated steady-state central compartment concentration-time profiles for linezolid across 10,000 virtual patients receiving 600 mg q24h. . . . . . . . . . . . . . . . . . . . . . . . . . . . 45
S35 Simulated steady-state central compartment concentration-time profiles for linezolid showing the mean and 90% prediction interval across 10,000 virtual patients receiving 600 mg q24h. . 46
S36 Schematic of the one-compartment population pharmacokinetic model for moxifloxacin, showing the absorption compartment ($C_q$) and central compartment ($C$), with first-order absorption rate constant (ka) and systemic clearance (CL_F). . . . . . . . . . . . . . . . . . . . . . . 47
S37 Steady-state $AUC_{0-24}$ distributions across 10,000 simulated patient profiles for moxifloxacin at 400 mg q24h and 800 mg q24h. Distributions reflect interindividual variability drawn from the parameter distributions in Table S10. . . . . . . . . . . . . . . . . . . . . . . . . . . . . 48
S38 Simulated steady-state central compartment concentration-time profiles for moxifloxacin across 10,000 virtual patients receiving 400 mg q24h and 800 mg q24h. . . . . . . . . . . . . . . . . 49

S39 Simulated steady-state central compartment concentration-time profiles for moxifloxacin showing the mean and 90% prediction interval across 10,000 virtual patients receiving 400 mg q24h and 800 mg q24h. . . . . 50
S40 Schematic of the population pharmacokinetic model for rifabutin, showing the transit-compartment absorption input ($m_{abs}$), rifabutin central compartment ($C_{C,RFB}$), rifabutin peripheral compartment ($C_{P,RFB}$), des-rifabutin central compartment ($C_{C,dRFB}$), and des-rifabutin peripheral compartment ($C_{P,dRFB}$), with first-pass metabolism fraction (F_m), absorption rate constant (k_a), systemic clearances (CL_1, CL_2), intercompartmental clearances (Q_1, Q_2), and central compartment conversion clearance (Q_3). . . . . 52
S41 Steady-state $AUC_{0-24}$ distributions across 10,000 simulated patient profiles for rifabutin at 300 mg q24h. Distributions reflect interindividual variability drawn from the parameter distributions in Table S11. . . . . 53
S42 Simulated steady-state central compartment concentration-time profiles for rifabutin across 10,000 virtual patients receiving 300 mg q24h. . . . . 54
S43 Simulated steady-state central compartment concentration-time profiles for rifabutin showing the mean and 90% prediction interval across 10,000 virtual patients receiving 300 mg q24h. . . 55
S44 Schematic of the one-compartment population pharmacokinetic model for rifampin with transit-compartment absorption, showing the absorption compartment ($q_0$) and central compartment ($C$), with absorption rate constant (k_a), transit rate constant (k_tr), bioavailability (F), and systemic clearance (CL/F). . . . . 57
S45 Steady-state $AUC_{0-24}$ distributions across 10,000 simulated patient profiles for rifampin at 600 mg q24h. Distributions reflect interindividual variability drawn from the parameter distributions in Table S12. . . . . 58
S46 Simulated steady-state central compartment concentration-time profiles for rifampin across 10,000 virtual patients receiving 600 mg q24h. . . . . 59
S47 Simulated steady-state central compartment concentration-time profiles for rifampin showing the mean and 90% prediction interval across 10,000 virtual patients receiving 600 mg q24h. . . 60
S48 Schematic of the two-compartment population pharmacokinetic model for tigecycline, showing the central compartment ($C$) and peripheral compartment ($C_p$), with systemic clearance (CL_F), intercompartmental clearance (Q_F), and 3-hour IV infusion input with loading dose. . 61
S49 Steady-state $AUC_{0-24}$ distributions across 10,000 simulated patient profiles for tigecycline at 25 mg q12h administered as a 3-hour IV infusion. Distributions reflect interindividual variability drawn from the parameter distributions in Table S13. . . . . 62
S50 Simulated steady-state central compartment concentration-time profiles for tigecycline across 10,000 virtual patients receiving a loading dose followed by 25 mg q12h as a 3-hour IV infusion. 63
S51 Simulated steady-state central compartment concentration-time profiles for tigecycline showing the mean and 90% prediction interval across 10,000 virtual patients receiving a loading dose followed by 25 mg q12h as a 3-hour IV infusion. . . . . 64
S52 Schematic of the four-compartment population pharmacokinetic model for omadacycline, showing two parallel absorption compartments ($q_1, q_2$), central compartment ($C$), and peripheral compartment ($P$), with absorption rate constants (k_a,1, k_a,2), lag time (T_lag), systemic clearance (CL_F,1), and intercompartmental clearance (CL_F,2). . . . . 66
S53 Steady-state $AUC_{0-24}$ distributions across 10,000 simulated patient profiles for omadacycline at 300 mg q24h. Distributions reflect interindividual variability drawn from the parameter distributions in Table S14. . . . . 67
S54 Simulated steady-state central compartment concentration-time profiles for omadacycline across 10,000 virtual patients receiving 300 mg q24h. . . . . 68
S55 Simulated steady-state central compartment concentration-time profiles for omadacycline showing the mean and 90% prediction interval across 10,000 virtual patients receiving 300 mg q24h. 69

### List of Tables

S1 Pharmacokinetic model summary across antibiotics: model structure, dosing used in the present work, typical literature AUC range, mean AUC and $C_{max}$ from the reimplemented model, and supporting references. . . . . . . . . . . . . . . . . . . . . . . . . . . . . . . . . . . . 7

S2 Population pharmacokinetic parameters for the two-compartment amikacin model. Values represent typical population means; IIV is expressed as percent coefficient of variation ($\%CV$). 9

S3 Population pharmacokinetic parameters for the ten-compartment bedaquiline model. Values represent typical population means; IIV is expressed as percent coefficient of variation ($\%CV$). 13

S4 Population pharmacokinetic parameters for the three-compartment clofazimine model. Values represent typical population means; IIV and IOV are expressed as percent coefficient of variation ($\%CV$). . . . . . . . . . . . . . . . . . . . . . . . . . . . . . . . . . . . . . . . 18

S5 Population pharmacokinetic parameters for the three-compartment clarithromycin model. Values represent typical population means; IIV is expressed as percent coefficient of variation ($\%CV$). . . . . . . . . . . . . . . . . . . . . . . . . . . . . . . . . . . . . . . . . 22

S6 Population pharmacokinetic parameters for the transit-compartment ethambutol model. Values represent typical population means; IIV and IOV are expressed as percent coefficient of variation ($\%CV$). . . . . . . . . . . . . . . . . . . . . . . . . . . . . . . . . . . . . . . . 28

S7 Population pharmacokinetic parameters for the one-compartment cefoxitin model. Values are the typical population means reported in the original source [32]; interindividual variability (IIV) is expressed as percent coefficient of variation ($\%CV$). . . . . . . . . . . . . . . . . . . . . 33

S8 Population pharmacokinetic parameters for the two-compartment imipenem model. Values represent typical population means; IIV is expressed as percent coefficient of variation ($\%CV$). 38

S9 Population pharmacokinetic parameters for the transit-compartment linezolid model. Values represent typical population means; IIV and IOV are expressed as percent coefficient of variation ($\%CV$). . . . . . . . . . . . . . . . . . . . . . . . . . . . . . . . . . . . . . . . 42

S10 Population pharmacokinetic parameters for the one-compartment moxifloxacin model. Values represent typical population means; IIV is expressed as percent coefficient of variation ($\%CV$). 47

S11 Population pharmacokinetic parameters for the transit-compartment rifabutin model with parallel des-rifabutin metabolite tracking. Values represent typical population means; IIV is expressed as percent coefficient of variation ($\%CV$). . . . . . . . . . . . . . . . . . . . . . . . . . 51

S12 Population pharmacokinetic parameters for the transit-compartment rifampin model. Values represent typical population means; $\theta$ denotes the dose-type effect coefficient, $\eta$ denotes the interindividual variability variance, and $\kappa$ denotes the interoccasional variability variance. . . 56

S13 Population pharmacokinetic parameters for the two-compartment tigecycline model. Values represent typical population means; IIV is expressed as percent coefficient of variation ($\%CV$). 61

S14 Population pharmacokinetic parameters for the two-compartment omadacycline model with parallel absorption. Values represent typical population means; IIV is expressed as percent coefficient of variation ($\%CV$). . . . . . . . . . . . . . . . . . . . . . . . . . . . . . . . . . . . 65

### A Comparison of literature versus reimplemented AUC values

Table S1: Pharmacokinetic model summary across antibiotics: model structure, dosing used in the present work, typical literature AUC range, mean AUC and $C_{max}$ from the reimplemented model, and supporting references.

| Antibiotic | # Comp. | Variability | Dose | Typical AUC range (mg·h/L) | Reimplemented Means ± s.d. | | References & comments |
|---|---|---|---|---|---|---|---|
| | | | | | AUC (mg·h/L) | $C_{max}$ (mg/L) | |
| Amikacin (AMK) [44] | 2 | IIV | 15 mg/kg 1h infusion QD | AUC: ∼150; $C_{max}$: 35–50 mg/L at end of 30–60 min infusion | 219.49 ± 46.39 | 59.00 ± 6.78 | AUC: 154.53 ± 29.91 mg·h/L in 6 HV [15]; 152.82 ± 50.83 mg·h/L [21]. $C_{max}$: 35–50 mg/L [39]; 35–45 mg/L median [23]; 55.30–16.88 mg/L [21]. |
| Bedaquiline (BDQ) [27] | 10 | IIV | 400/200 mg | ∼20–24 at steady state | 18.99 ± 4.97 | 1.39 ± 0.38 | 21.648 mg·h/L at wk 8 [13]; ∼25 mg·h/L [29]. |
| Cefoxitin (FOX) [32] | 3 | IIV | 4 g BID | ≈448 for a 4 g 5-min infusion BID; ≈500 for a 4 g 8-h infusion BID | 830.10 ± 460.47 | 92.73 ± 34.68 | Concentration–time profiles replotted and AUCs estimated from [11] and [25], assuming dose proportionality. |
| Clarithromycin (CLR) [20] | 3 | IIV | 500 mg BID | 36 (range 17–67) | 29.27 ± 3.92 | 1.70 ± 0.22 | 15–20 mg·h/L for ONE 500 mg dose [36]; 36 mg·h/L total AUC for TWO 500 mg doses [4]. |
| Clofazimine (CFZ) [1] | 4 | IIV, IOV | 100 mg QD | 4–16 | 6.93 ± 1.38 | 0.34 ± 0.07 | Weekly AUC reported as 60 mg·h/L → ∼8.5 daily; $C_{ave}$ of 0.36 $\mu$g/mL [2] → AUC ≈ 8.64 mg·h/L; 15.97 mg·h/L (IQR 11.7–21.7) [18], but the same paper shows daily AUC = 7.3 mg·h/L at 200 mg, implying ∼4 mg·h/L at 100 mg. |
| Ethambutol (EMB) [24] | 3 | IIV, IOV | 15 mg/kg QD | 13–23 at 15 mg/kg | 22.15 ± 6.15 | 2.76 ± 0.82 | ∼25–29 mg·h/L at 25 mg/kg [34]; 23.6 mg·h/L (IQR 20.5–28.9) at 15–25 mg/kg [12]; 13 mg·h/L at 15 mg/kg [19]. |
| Imipenem (IPM) [9] | 2 | IIV | 1000 mg 3h infusion BID | 120–288 | 229.05 ± 40.94 | 25.97 ± 3.05 | 63.9 ± 9.1 mg·h/L after ONE 1-g infusion → ∼128 mg·h/L BID [40]; 109.8 mg·h/L (103.2–116.8) after ONE 1-g infusion → ∼220 mg·h/L BID [45]; 144.22 ± 73.63 mg·h/L after ONE 2-h 1-g infusion → ∼288 mg·h/L BID [22]. |
| Linezolid (LZD) [3] | 2 | IIV, IOV | 600 mg QD | 80–130 | 183.92 ± 77.57 | 14.42 ± 4.06 | 84.6 mg·h/L (64.7–125.0) [28]; 138.1 ± 27.6 on D1 and 234 ± 55.2 on D7 at 600 mg BID [8]; 80–128 mg·h/L [42]; $AUC_{0-12}$ = 96.73 ± 56.45 mg·h/L [14]. |
| Moxifloxacin (MXF) [38] | 2 | IIV | 400 mg QD | 20–55 at 400 mg QD | 43.37 ± 13.41 | 3.96 ± 1.09 | 27–48 mg·h/L [31]; 55 (36–79) mg·h/L [35]; ∼40 mg·h/L [49]; 32.78 mg·h/L (IQR 22.75–47.31) [33]; 19–30 mg·h/L [10]. |
| Rifampicin (RIF) [46] | 2 | IIV | 600 mg QD | 15–60 | 33.19 ± 19.37 | 6.34 ± 2.62 | Meta-analysis summary estimate of 38.73 mg·h/L at steady state across 35 studies [43]. |
| Rifabutin (RBT) [16] | 5 | IIV | 300 mg QD | 3–5 | 4.50 ± 2.17 | 0.45 ± 0.20 | 2.71 (1.39–3.98) mg·h/L at wk 2–4; 2.97 (0.60–4.67) mg·h/L at wk 4–5; 4.36 (1.73–6.09) mg·h/L at wk 6–7 [17]; 4.08 mg·h/L [7]; 5.4 mg·h/L [41]. |
| Tigecycline (TIG) [6] | 2 | IIV | 25 mg 1h infusion BID | 1.6–3.5 | 3.68 ± 1.68 | 0.37 ± 0.10 | 2.3 mg·h/L [30]; 1.6 mg·h/L (NDA document); $AUC_{0-12}$ = 1.784 ± 0.173 mg·h/L at 25 mg → $AUC_{0-24}$ ≈ 3.5 mg·h/L [47]. |
| Omadacycline (OMC) [48] | 4 | IIV | 300 mg QD | 9–20 | 20.13 ± 4.24 | 1.63 ± 0.30 | 19.4–20.4 mg·h/L [48]; 11.156 mg·h/L [26]; 10.19 (8.35–11.01) [37]; 8.5–13 mg·h/L ([5], Table S5). |

### B Introduction

All of the following antibiotic models are available for download at github.itap.purdue.edu/ElsjePienaarGroup. Several equations recur across the drug-specific models and are collected here; subsequent sections reference them as needed rather than restating them.

*Infusion rate*

The infusion rate used by all intravenously administered drugs is given by:

$$\texttt{infusion rate} = R(t) = \begin{cases} \frac{\texttt{dose}}{\tau_{\text{inf}}} & 0 \leq t < \tau_{\text{inf}} \\ 0 & t \geq \tau_{\text{inf}} \end{cases} \tag{1}$$

where $R(t)$ is the infusion rate as a function of time, $\texttt{dose}$ is the administered dose amount (mg) for the dosing period, $\tau_{\text{inf}}$ is the infusion duration (h), and $t$ is the elapsed time within the current dosing period.

*Oral absorption*

The gamma constant used in the oral absorption compartment is given by:

$$\Gamma_{\texttt{n}} = \sqrt{2\pi} \cdot \texttt{n}^{\texttt{n}+0.5} \cdot \exp(-\texttt{n}) \tag{2}$$

where $\Gamma_n$ is the constant used in the oral absorption compartment and $\texttt{n}$ is the number of transport compartments.

*Interindividual and interobservational variability*

Interindividual and interobservational variability are introduced by:

$$\theta_i = \theta \cdot \exp \eta_i \tag{3}$$

where $\theta_i$ is the parameter value for the $i$-th subject, $\theta$ is the mean value, and $\eta_i$ is a random interindividual or interobservational variable, normally distributed with a mean of zero and variance $\omega^2$.

The coefficient of variation is related to the log-domain variance by:

$$\text{CV} = \sqrt{\exp(\omega^2) - 1} \tag{4}$$

Equation 4 is used to convert between the coefficient of variation of the parameter values and the variance in the log-domain ($\omega^2$), depending on which value is reported in the source paper.

The infusion rate (Equation 1) is used by all intravenous (IV) drugs (AMK, FOX, IMI, TIG). The gamma ($\Gamma$) term (Equation 2) is used by EMB and RIF. The CV relationship (Equation 4) is used by every model carrying an interindividual variability (IIV) or interobservational variability (IOV) to determine the patient-specific coefficient values for the Monte Carlo simulations.

### C Amikacin

#### C.1 Amikacin model parameters

Table S2: Population pharmacokinetic parameters for the two-compartment amikacin model. Values represent typical population means; IIV is expressed as percent coefficient of variation ($\%CV$).

| Parameter | Mean | IIV % CV | Units |
|---|---|---|---|
| Vc | 8.92 | 15 | L |
| Vp | 11.4 | 25 | L |
| CL | 3.6 | 21 | L/h |
| Q | 4.43 | 30 | L/h |

`Vc` represents volume of distribution for the central compartment; `Vp` represents the volume of distribution for the peripheral compartment; `CL` represents the system clearance; `Q` represents the transfer rate between the central compartment and the peripheral compartment.

#### C.2 Amikacin model equations

This model was recreated from the model proposed in [44]. The parameter values used in the patient simulations were calculated using Equation 4. The model structure was a two compartment model as a central compartment ($C_1$) and a peripheral compartment ($C_2$) as shown in Figure S2.

$$\frac{d(C_1)}{dt} = \texttt{infusion rate} + \frac{1}{\texttt{Vc}}\Big[(\texttt{Q} \cdot C_1) - (\texttt{Q} \cdot C_2) - (\texttt{CL} \cdot C_1)\Big] \tag{5}$$

$$\frac{d(C_2)}{dt} = \frac{1}{\texttt{Vp}}\Big[(\texttt{Q} \cdot C_1) - (\texttt{Q} \cdot C_2)\Big] \tag{6}$$

#### C.3 Amikacin model flowchart

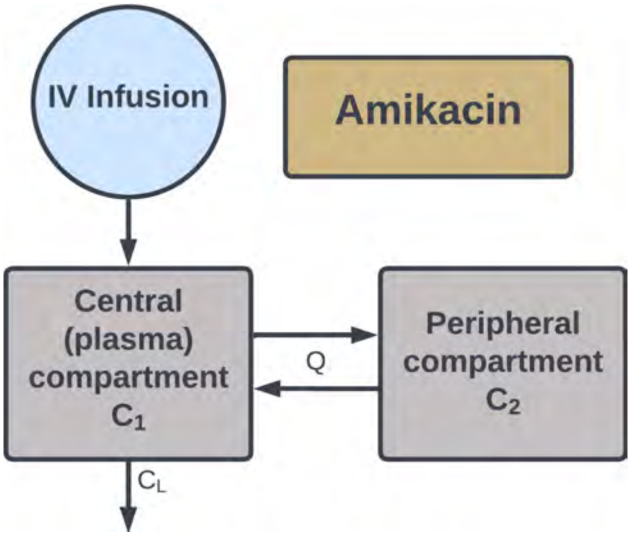

Figure S2: Schematic of the two-compartment population pharmacokinetic model for amikacin, showing drug transfer between the central ($C_1$) and peripheral ($C_2$) compartments, systemic clearance (`CL`), and intercompartmental transfer (`Q`).

#### C.4 Amikacin model AUC distributions

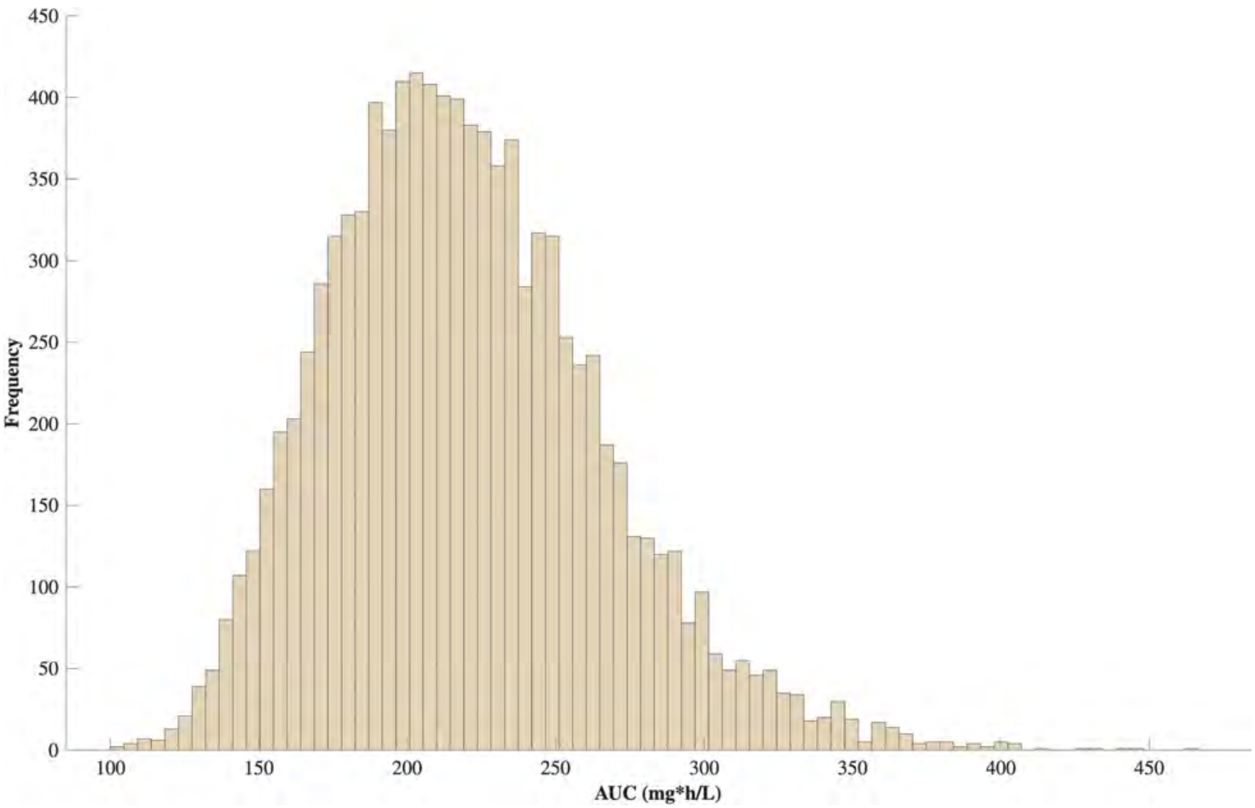

Figure S3: Steady-state $AUC_{0-24}$ distributions across 10,000 simulated patient profiles for amikacin at 15 mg/kg q24h. Distributions reflect interindividual variability drawn from the parameter distributions in Table S2.

#### C.5 Amikacin model concentration time-curves

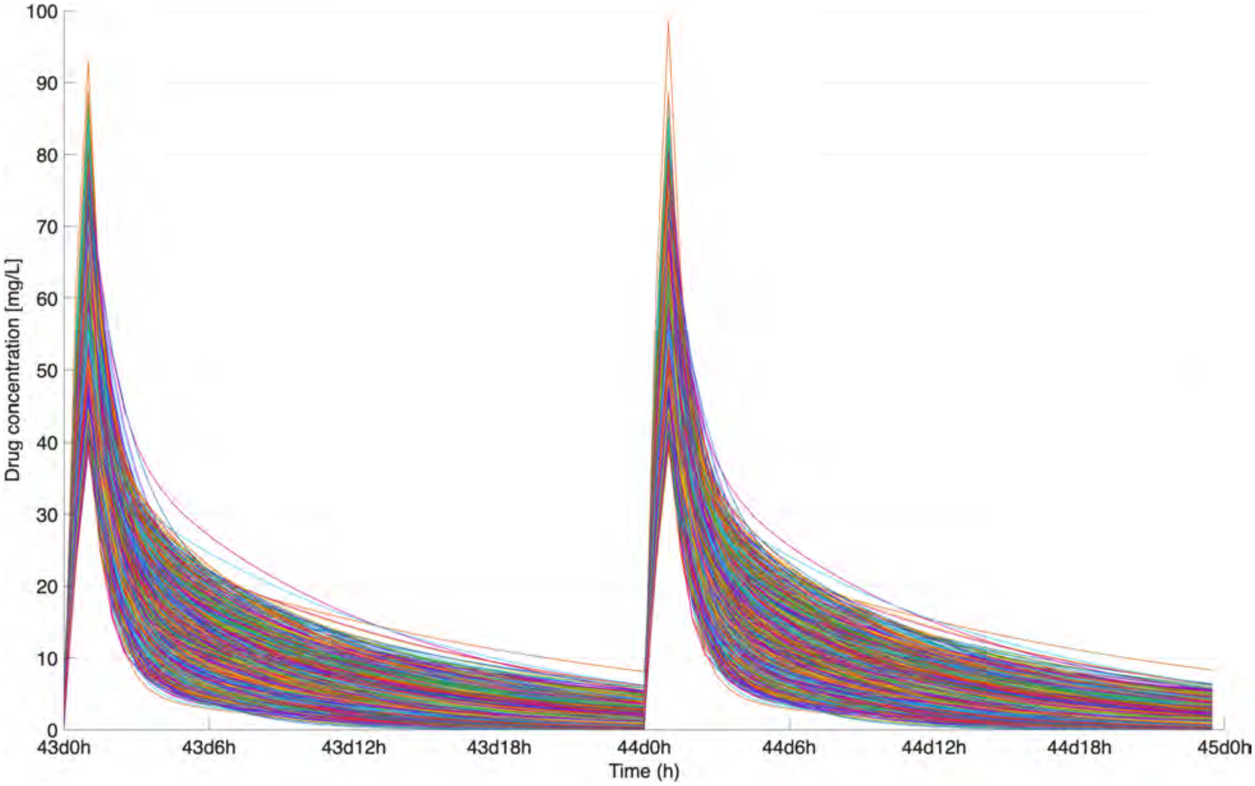

Figure S4: Simulated steady-state central compartment concentration-time profiles for amikacin across 10,000 virtual patients receiving 15 mg/kg q24h.

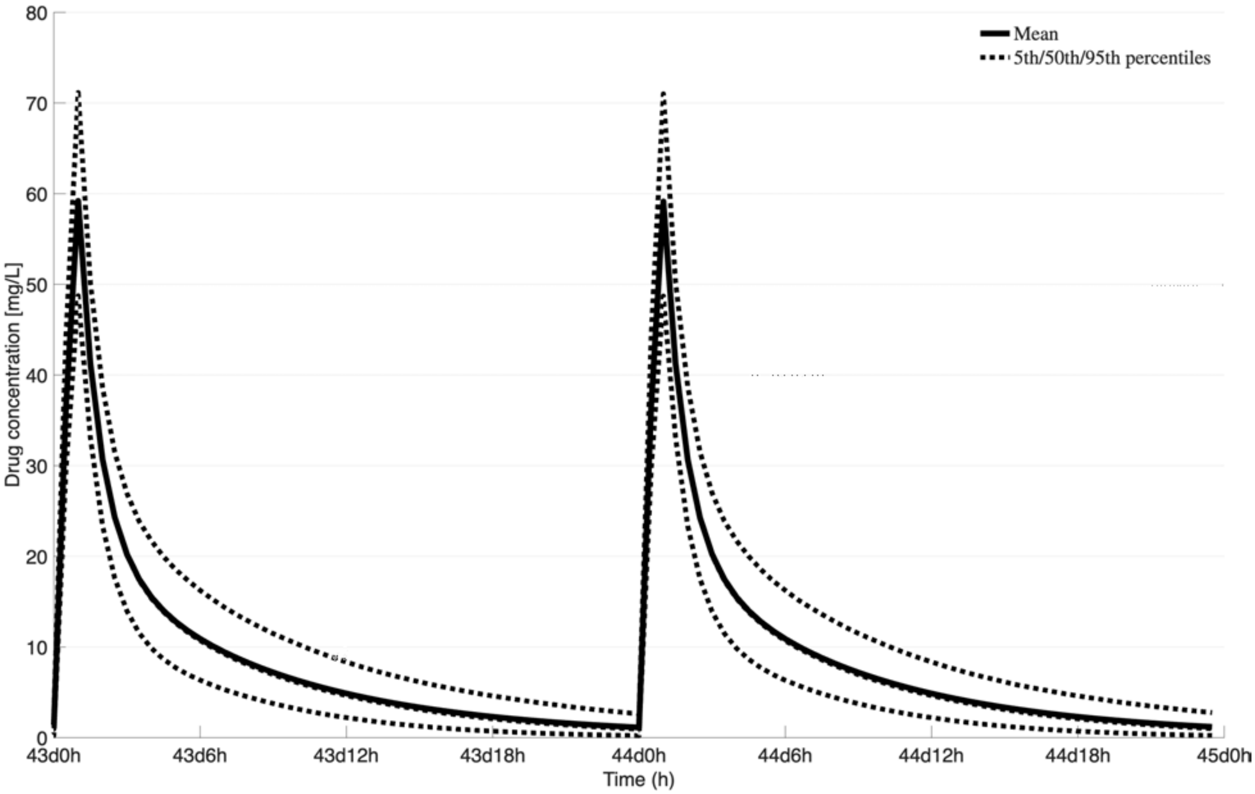

Figure S5: Simulated steady-state central compartment concentration-time profiles for amikacin showing the mean and 90% prediction interval across 10,000 virtual patients receiving 15 mg/kg q24h.

### D Bedaquiline

#### D.1 Bedaquiline model parameters

Table S3: Population pharmacokinetic parameters for the ten-compartment bedaquiline model. Values represent typical population means; IIV is expressed as percent coefficient of variation ($\%CV$).

| Parameter | Mean | IIV % CV | Units | Parameter | Mean | IIV % CV | Units |
|---|---|---|---|---|---|---|---|
| V_C | 120 | 51 | L | Q_2 | 6.8 | 26 | L/h |
| V_2 | 79 | 27 | L | Q_3 | 7.3 | 59 | L/h |
| V_3 | 750 | 45 | L | k_a | 0.9 | 29 | 1/h |
| CL | 6.7 | 30 | L/h | k_tr | 2.6 | 49 | 1/h |

V_C is volume in central compartment; V_2 is volume in peripheral 2 compartment; V_3 is volume in peripheral 3 compartment; CL is clearance rate constant; Q_2 is inter-compartmental clearance constant between central and peripheral 2; Q_3 is inter-compartmental clearance constant between central and peripheral 3 compartment; k_a is first order absorption rate constant; k_tr is transit rate constant between transit compartments.

#### D.2 Bedaquiline model equations

This model was recreated from the model proposed in [27]. The parameter values used in the patient simulations were calculated using Equation 4. The model structure was a 10 compartment model with an absorption compartment ($q0$), 5 transit compartments ($q1 - q5$), a final transit compartment ($q$), a central compartment ($C$), and two peripheral compartments ($C_2$ & $C_3$) as shown in Figure S6.

$$\frac{d(q_0)}{dt} = -\texttt{k\_tr} \cdot (q0) \tag{7}$$

$$\frac{d(q_i)}{dt} = \texttt{k\_tr}\Big((q_{i-1}) - (q_i)\Big), \quad i = 1, \ldots, 5 \tag{8}$$

$$\frac{d(q)}{dt} = -\texttt{k\_a} \cdot (q) + \texttt{k\_tr} \cdot (q_5) \tag{9}$$

$$\frac{d(C)}{dt} = \frac{1}{\texttt{V\_C}}\Big[\texttt{k\_a} \cdot (q) - \texttt{Q\_2}\big((C) - (C_2)\big) - \texttt{Q\_3}\big((C) - (C_3)\big) - \texttt{CL} \cdot (C)\Big] \tag{10}$$

$$\frac{d(C_2)}{dt} = \frac{1}{\texttt{V\_2}}\Big[\texttt{Q\_2}\big((C) - (C_2)\big)\Big] \tag{11}$$

$$\frac{d(C_3)}{dt} = \frac{1}{\texttt{V\_3}}\Big[\texttt{Q\_3}\big((C) - (C_3)\big)\Big] \tag{12}$$

##### D.3 BDQ model flowchart

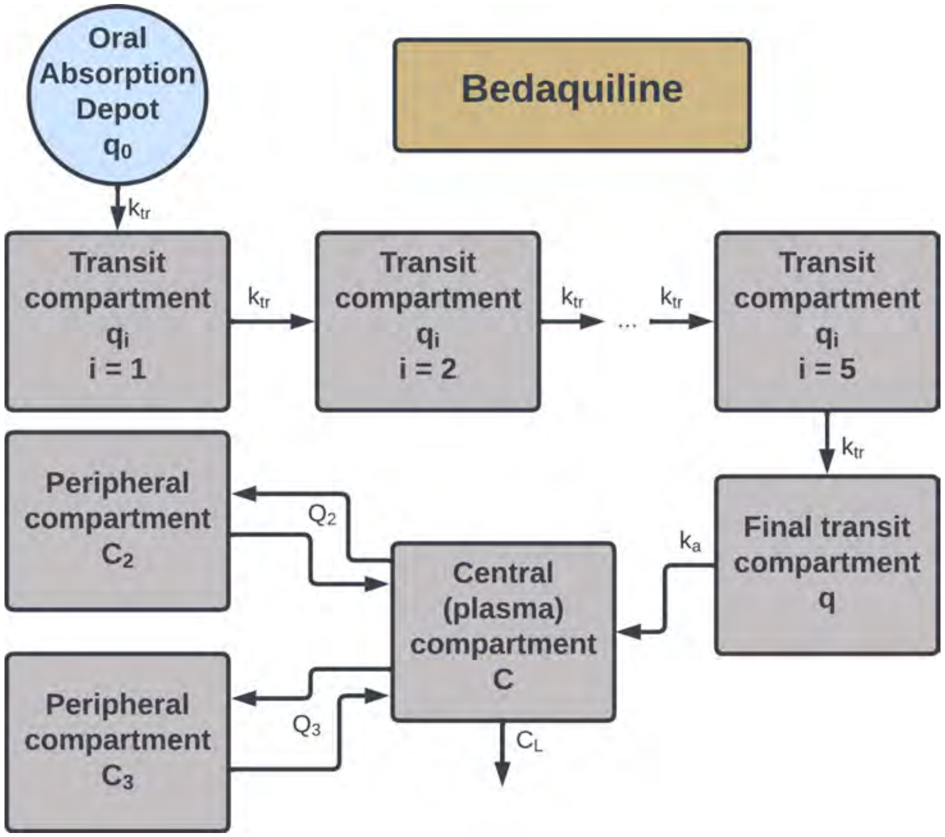

Figure S6: Schematic of the ten-compartment population pharmacokinetic model for bedaquiline, showing the absorption compartment ($q_0$), five transit compartments ($q_1$–$q_5$), final transit compartment ($q$), central compartment ($C$), and two peripheral compartments ($C_2$) and ($C_3$), with systemic clearance (CL) and intercompartmental transfer (Q_2, Q_3).

##### D.4 Bedaquiline model AUC distributions

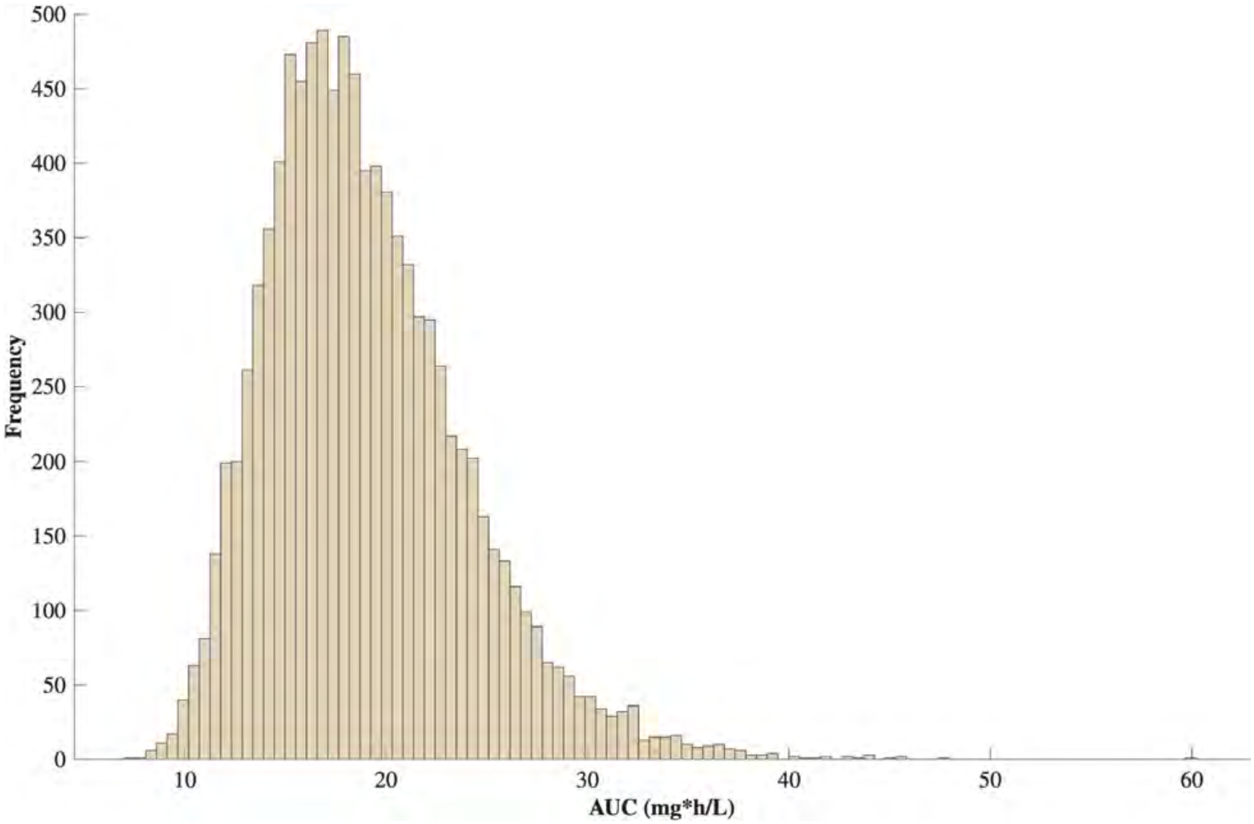

Figure S7: Steady-state $AUC_{0-24}$ distributions across 10,000 simulated patient profiles for bedaquiline at 400 mg q24h for two weeks followed by 200 mg three times weekly. Distributions reflect interindividual variability drawn from the parameter distributions in Table S3.

#### D.5 Bedaquiline model concentration time-courses

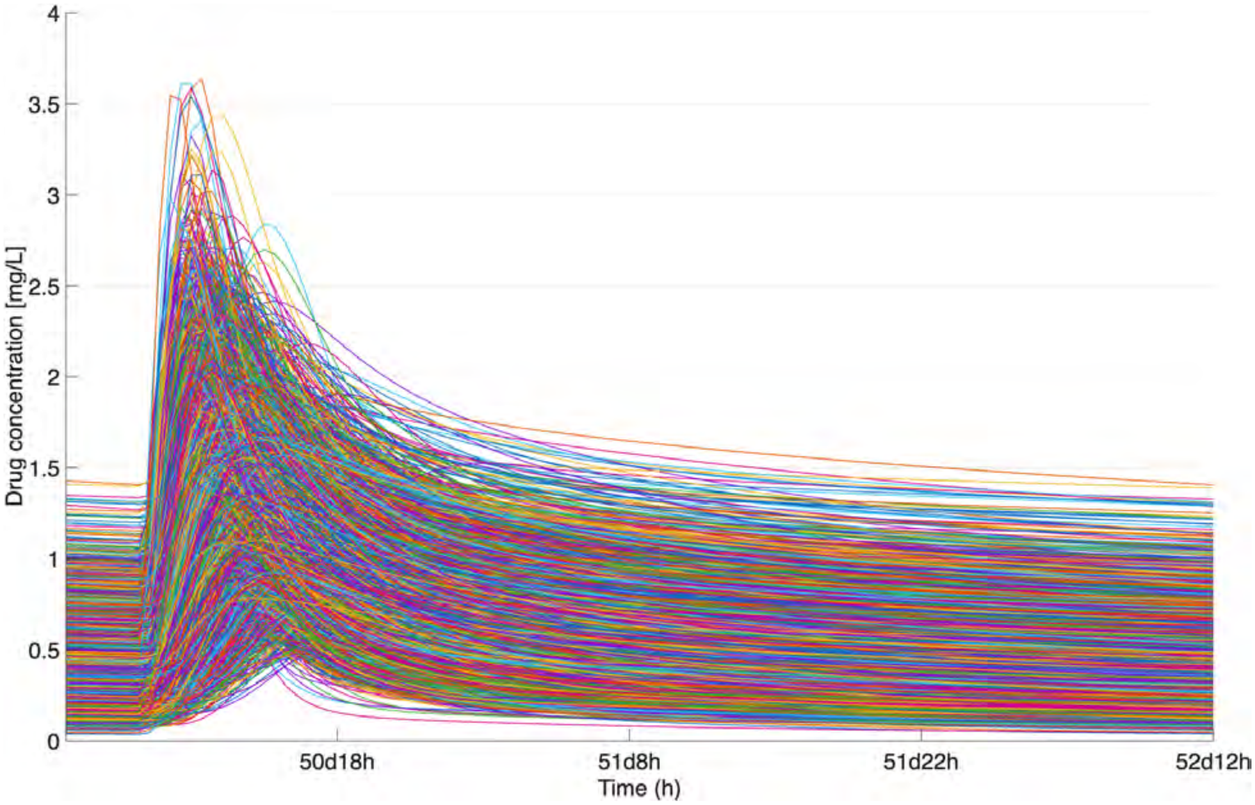

Figure S8: Simulated steady-state central compartment concentration-time profiles for bedaquiline across 10,000 virtual patients during the maintenance phase of 200 mg three times weekly (following 400 mg q24h for two weeks).

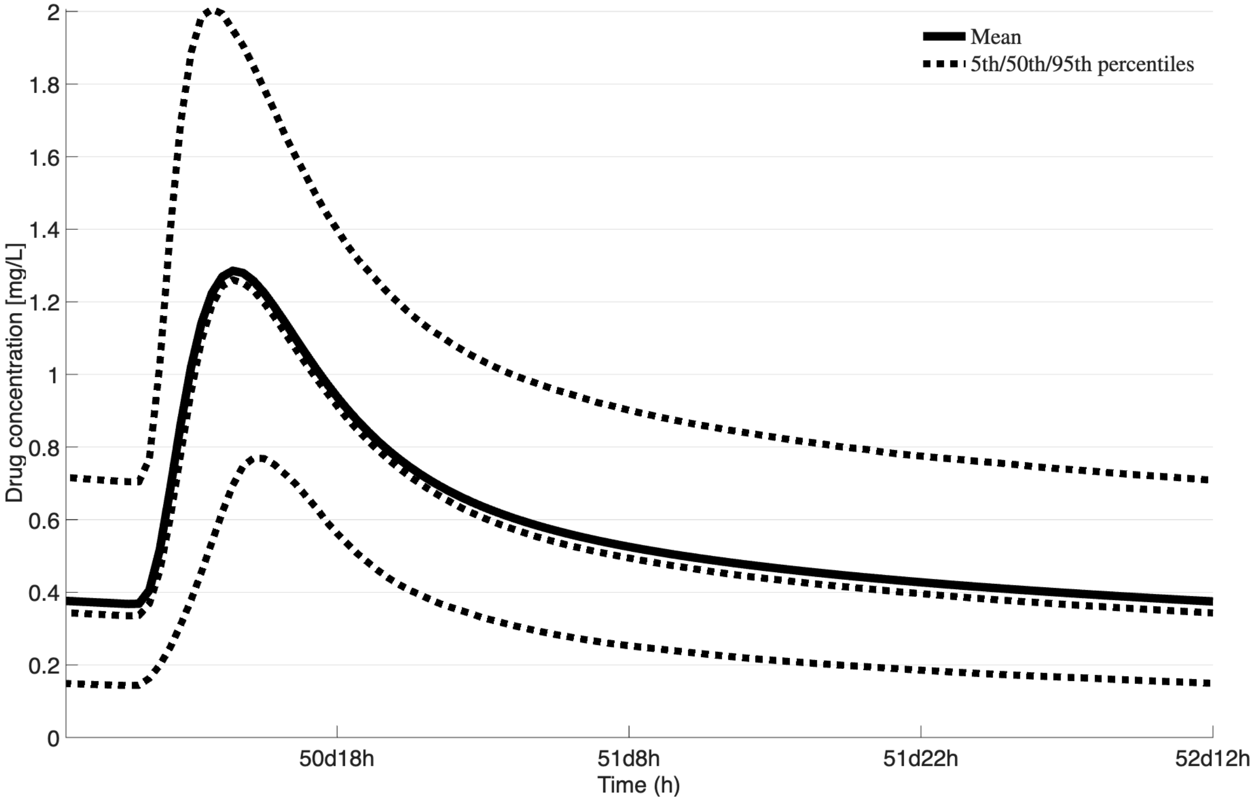

Figure S9: Simulated steady-state central compartment concentration-time profiles for bedaquiline showing the mean and 90% prediction interval across 10,000 virtual patients during the maintenance phase of 200 mg three times weekly (following 400 mg q24h for two weeks).

### E Clofazimine

#### E.1 Clofazimine model parameters

Table S4: Population pharmacokinetic parameters for the three-compartment clofazimine model. Values represent typical population means; IIV and IOV are expressed as percent coefficient of variation ($\%CV$).

| Parameter | Mean | IIV % CV | IOV % CV | Units |
|---|---|---|---|---|
| V_C | 262 | 23.5 | – | L |
| V_p1 | 10500 | 29.6 | – | L |
| V_p2 | 889 | 54.6 | – | L |
| CL/F | 11.5 | 25.6 | – | L/h |
| n | 4.75 | – | – | [–] |
| Q1/F | 56.3 | – | – | L/h |
| Q2/F | 86 | – | – | L/h |
| k_a | 0.209 | – | 32.6 | 1/h |
| MTT | 1.41 | – | 46.6 | 1/h |

V_C is volume in central compartment; V_p1 is volume in peripheral 1 compartment; V_p2 is volume in peripheral 2 compartment; CL/F is clearance rate constant; Q1/F is inter-compartmental clearance constant between central and peripheral 1; Q2/F is inter-compartmental clearance constant between central and peripheral 2 compartment; k_a is first order absorption rate constant; MTT is the mean transit time between transit compartments; n is the number of transit compartments.

#### E.2 Clofazimine model equations

This model was recreated from the model proposed in [1]. The parameter values used in the patient simulations were calculated using Equation 4. The model structure was a 4 compartment model with an absorption compartment ($q0$), a central compartment ($C$), and two peripheral compartments ($C_{p1}$ & $C_{p2}$).

$$\texttt{k\_tr} = \frac{\texttt{n}+1}{\texttt{MTT}} \tag{13}$$

$$\texttt{q0\_frac} = \Big(\texttt{k\_tr} \cdot t\Big)^{\texttt{n}} \tag{14}$$

$$\frac{d(q0)}{dt} = \texttt{dose} \cdot \texttt{F} \cdot \texttt{k\_tr} \cdot \texttt{q0\_frac} - \texttt{k\_a} \cdot (q0) \tag{15}$$

$$\frac{d(C)}{dt} = \frac{1}{\texttt{V\_C}}\Big[\texttt{k\_a} \cdot (q0) - \texttt{Q1/F}\big(C - C_{p1}\big) - \texttt{Q2/F}\big(C - C_{p2}\big) - \texttt{CL} \cdot (C)\Big] \tag{16}$$

$$\frac{d(C_{p1})}{dt} = \frac{1}{\texttt{V\_pq}}\Big[\texttt{Q1/F}\big(C - C_{p1}\big)\Big] \tag{17}$$

$$\frac{d(C_{p2})}{dt} = \frac{1}{\texttt{V\_p2}}\Big[\texttt{Q2/F}\big(C - C_{p2}\big)\Big] \tag{18}$$

#### E.3 Clofazimine model flowchart

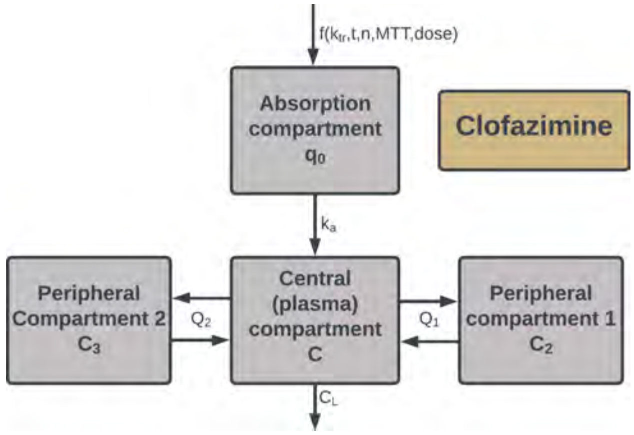

Figure S10: Schematic of the three-compartment population pharmacokinetic model for clofazimine, showing the absorption compartment ($q_0$), central compartment ($C$), and two peripheral compartments ($C_{p1}$) and ($C_{p2}$), with systemic clearance (`CL/F`) and intercompartmental transfer (`Q1/F`, `Q2/F`).

#### E.4 Clofazimine model AUC distributions

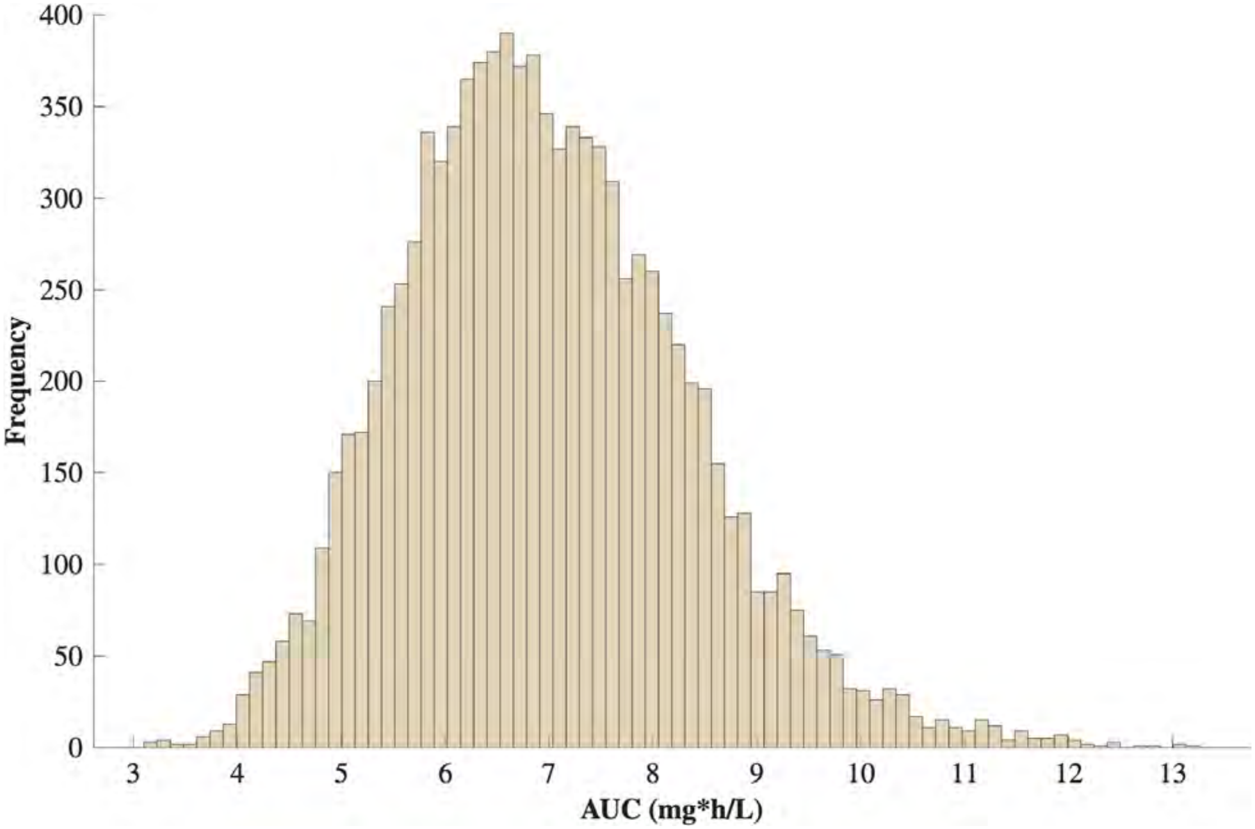

Figure S11: Steady-state $AUC_{0-24}$ distributions across 10,000 simulated patient profiles for clofazimine at 100 mg q24h. Distributions reflect interindividual variability drawn from the parameter distributions in Table S4.

#### E.5 Clofazimine model concentration time-courses

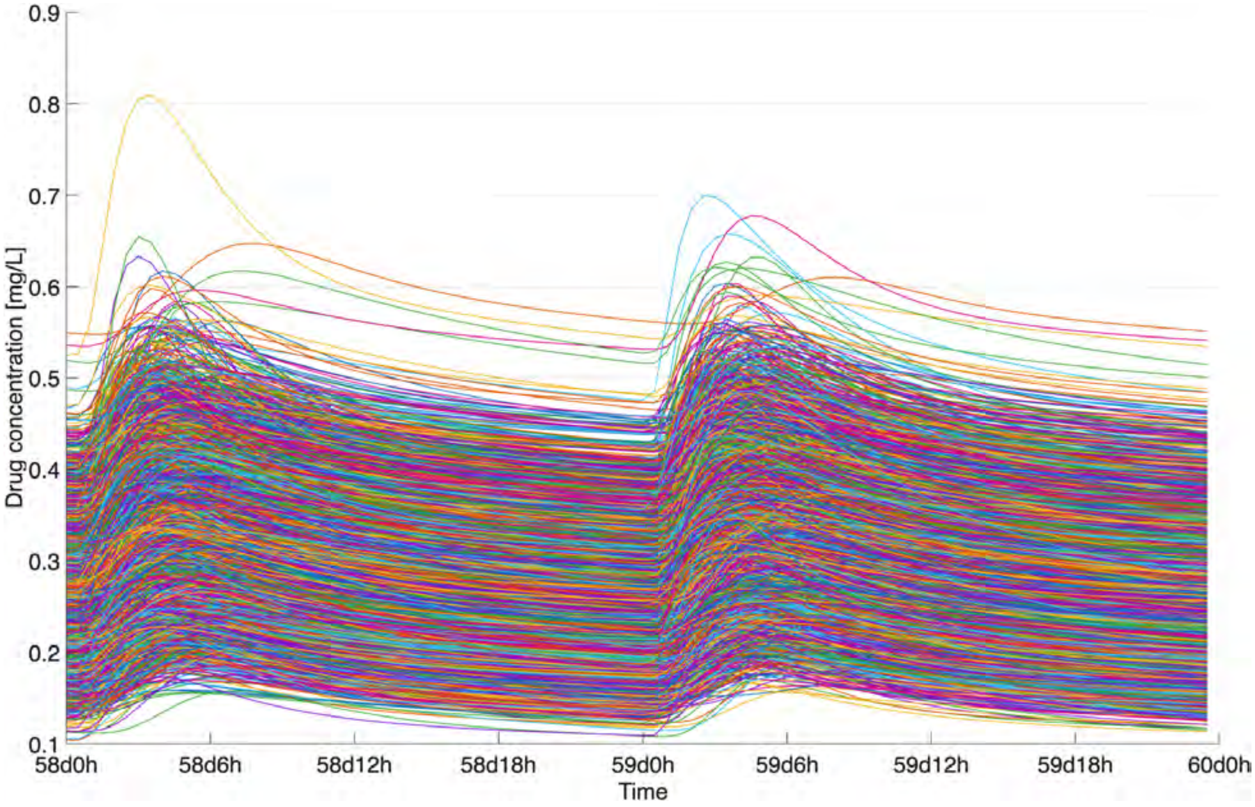

Figure S12: Simulated steady-state central compartment concentration-time profiles for clofazimine across 10,000 virtual patients receiving 100 mg q24h.

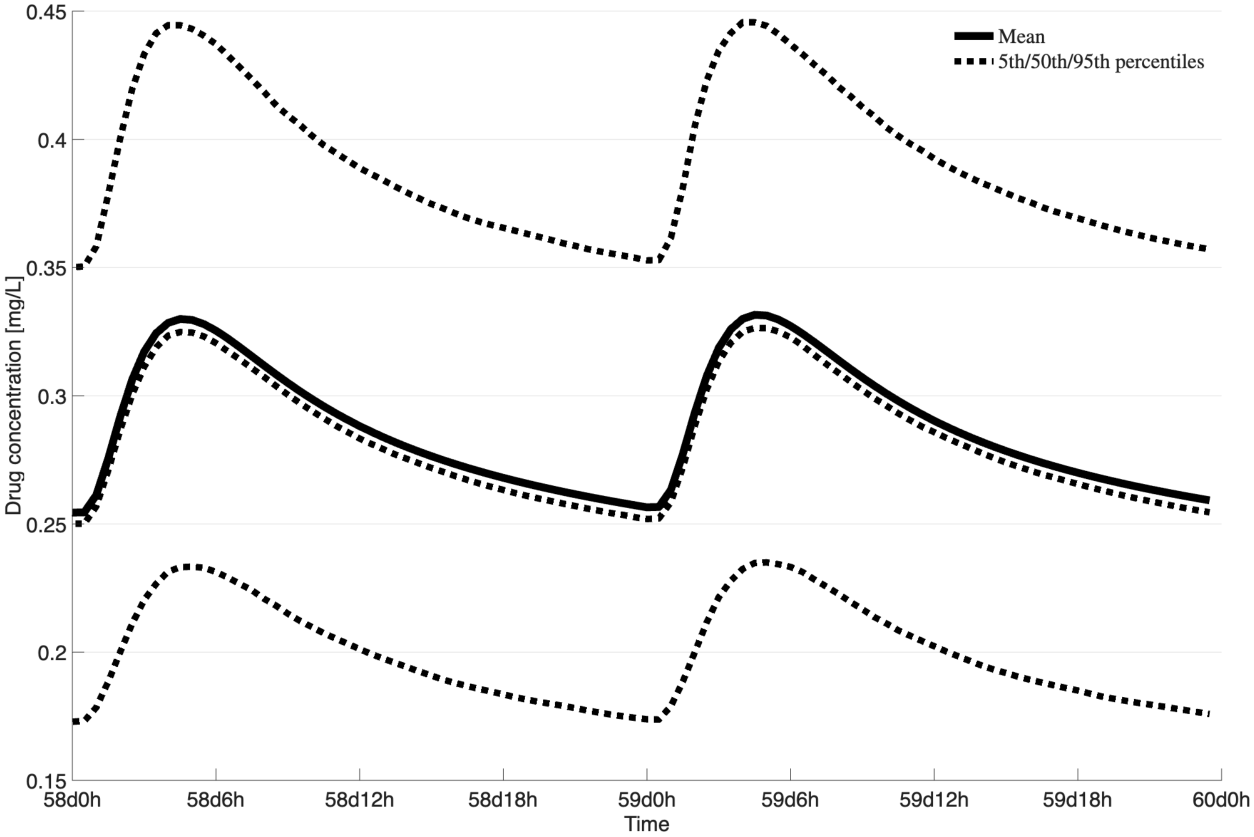

Figure S13: Simulated steady-state central compartment concentration-time profiles for clofazimine showing the mean and 90% prediction interval across 10,000 virtual patients receiving 100 mg q24h.

### F Clarithromycin

#### F.1 Clarithromycin model parameters

Table S5: Population pharmacokinetic parameters for the three-compartment clarithromycin model. Values represent typical population means; IIV is expressed as percent coefficient of variation ($\%CV$).

| Parameter | Mean | IIV % CV | Units |
|---|---|---|---|
| V1_F | 204.7 | 12.0 | L |
| V2_F | 168.9 | – | L |
| V3_F | 67.1 | 34.0 | L |
| CL/F | 34.4 | 13.2 | L/h |
| k_a | 0.680 | 33.9 | 1/h |
| K_12 | 0.019 | – | 1/h |
| K_21 | 0.434 | – | 1/h |
| K_13 | 0.667 | 0.133 | 1/h |
| K_31 | 0.260 | 0.128 | 1/h |

V1_F is volume in central compartment; V2_F is volume in peripheral compartment; V3_F is volume in epithelial compartment; CL/F is clearance rate constant; k_a is first order absorption rate constant; K_12 is inter-compartmental clearance constant between central and peripheral; K_21 is inter-compartmental clearance constant between peripheral and central; K_13 is inter-compartmental clearance constant between central and epithelial compartment; K_31 is inter-compartmental clearance constant between epithelial and central compartment.

#### F.2 Clarithromycin model equations

This model was recreated from the model proposed in [20]. The parameter values used in the patient simulations were calculated using Equation 4. The model structure was a 3 compartment model consisting of an absorption compartment ($q_0$), a central compartment ($C$), a peripheral compartment ($C_2$), and an epithelial compartment ($C_3$).

$$\frac{d(C)}{dt} = \frac{1}{\texttt{V1\_F}}\Big[\texttt{k\_a} \cdot q_0 - \texttt{K\_12} \cdot \texttt{V1\_F} \cdot C + \texttt{K\_21} \cdot \texttt{V2\_F} \cdot C_2 - \texttt{K\_13} \cdot \texttt{V1\_F} \cdot C + \texttt{K\_31} \cdot \texttt{V3\_F} \cdot C_3 - \texttt{CL/F} \cdot C\Big] \tag{19}$$

$$\frac{d(C_2)}{dt} = \frac{1}{\texttt{V2\_F}}\Big[\texttt{K\_12} \cdot \texttt{V1\_F} \cdot C - \texttt{K\_21} \cdot \texttt{V2\_F} \cdot C_2\Big] \tag{20}$$

$$\frac{d(C_3)}{dt} = \frac{1}{\texttt{V3\_F}}\Big[\texttt{K\_13} \cdot \texttt{V1\_F} \cdot C - \texttt{K\_31} \cdot \texttt{V3\_F} \cdot C_3\Big] \tag{21}$$

#### F.3 Clarithromycin model flowchart

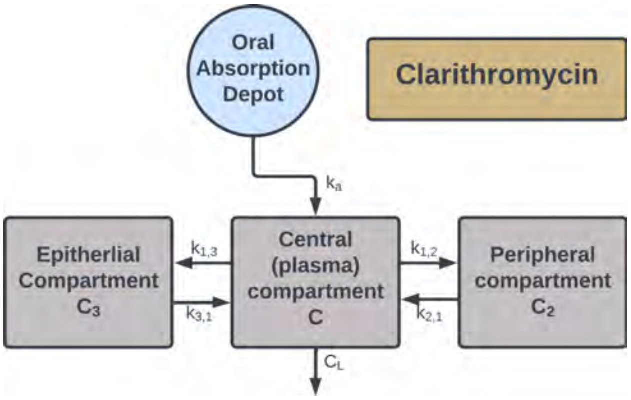

Figure S14: Schematic of the three-compartment population pharmacokinetic model for clarithromycin, showing the absorption compartment ($q_0$), central compartment ($C$), peripheral compartment ($C_2$), and epithelial compartment ($C_3$), with systemic clearance (CL/F) and intercompartmental transfer rate constants (K_12, K_21, K_13, K_31).

#### F.4 Clarithromycin model AUC distributions

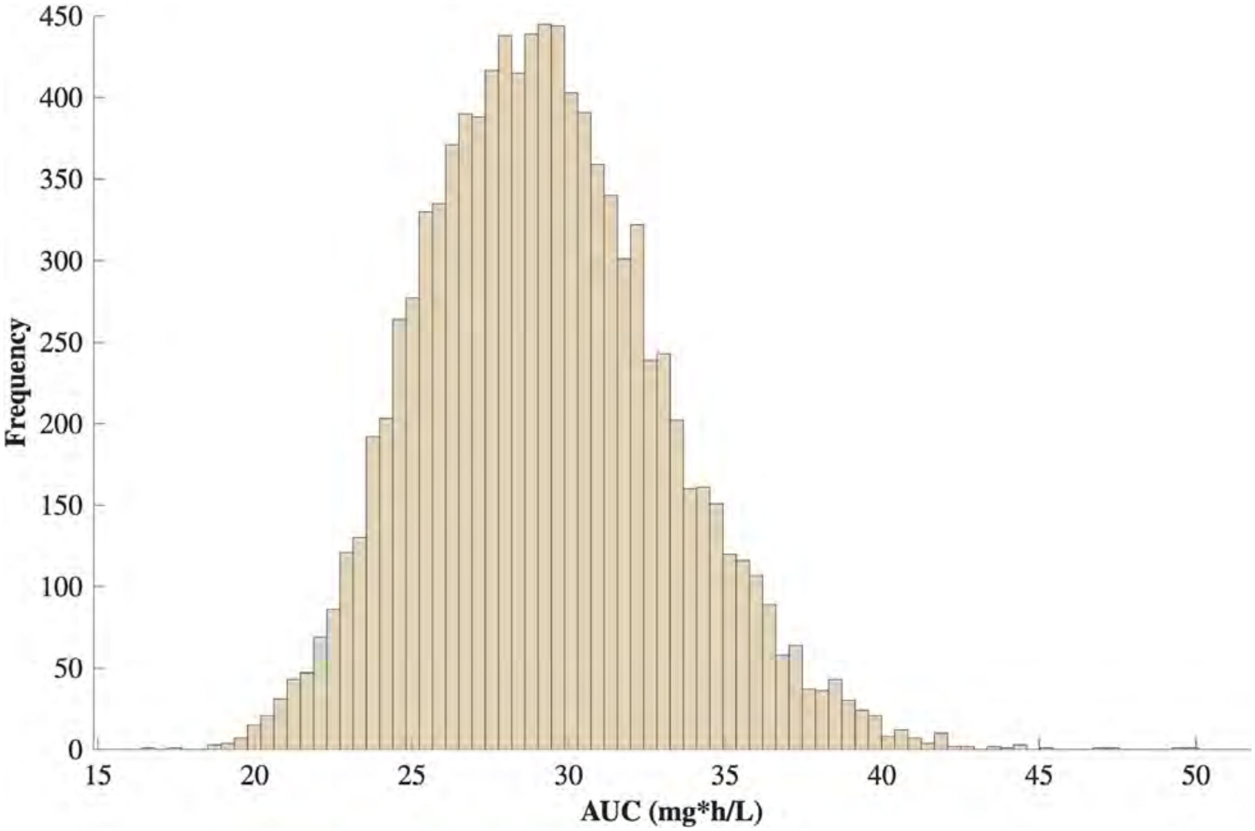

Figure S15: Steady-state $AUC_{0-24}$ distributions across 10,000 simulated patient profiles for clarithromycin at 500 mg q12h. Distributions reflect interindividual variability drawn from the parameter distributions in Table S5.

#### F.5 Clarithromycin model concentration time-courses

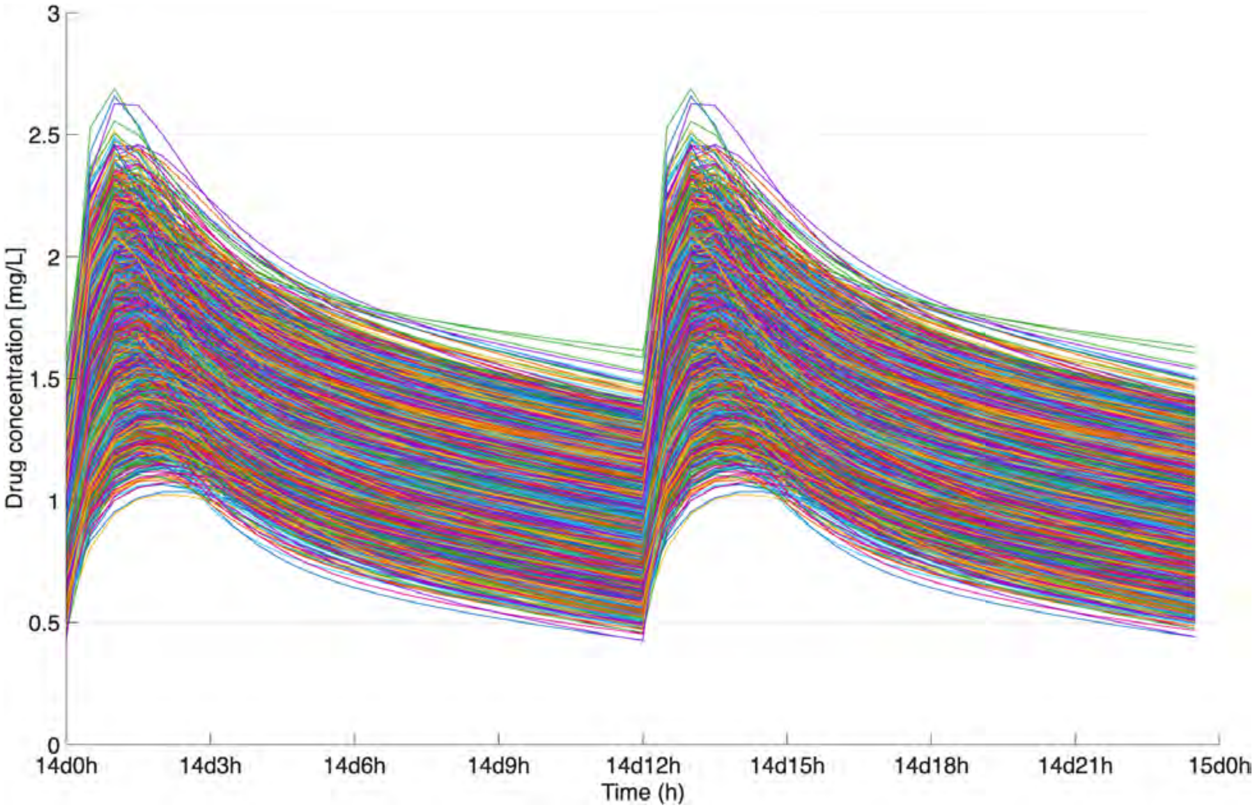

Figure S16: Simulated steady-state central compartment concentration-time profiles for clarithromycin across 10,000 virtual patients receiving 500 mg q12h.

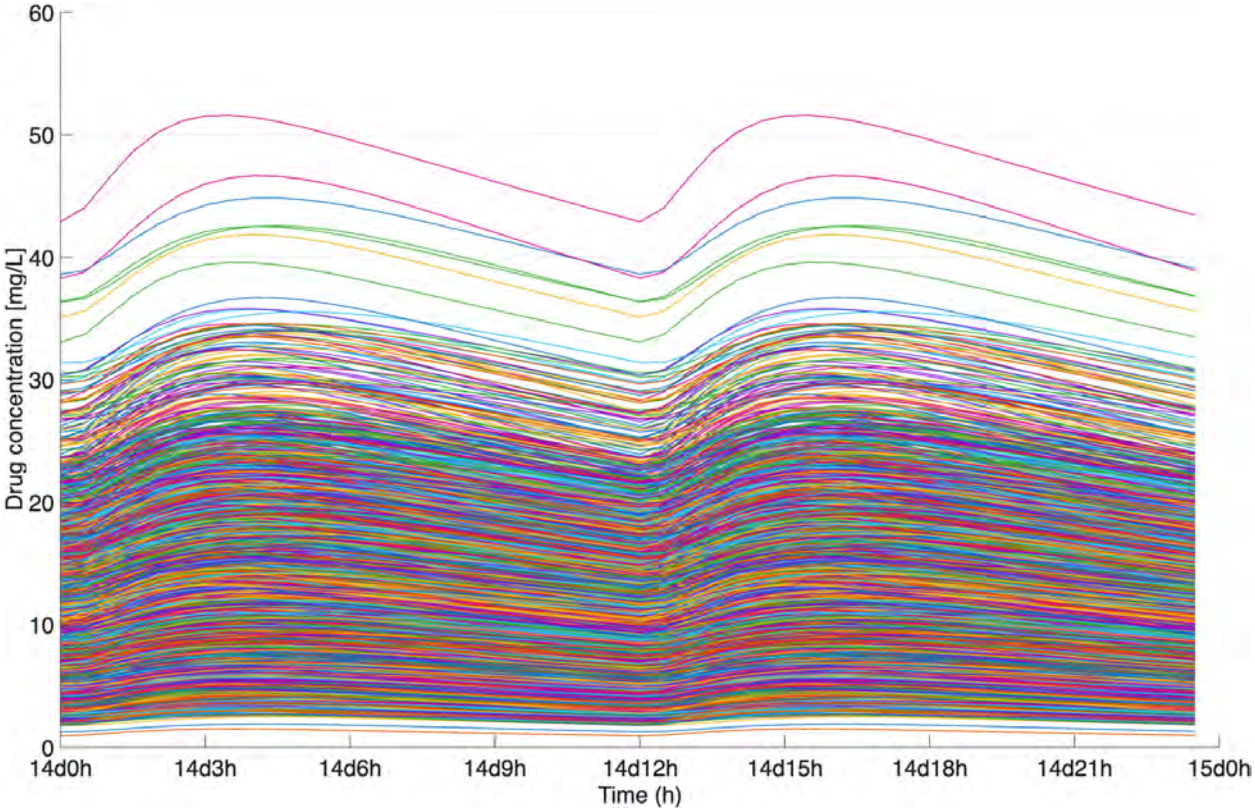

Figure S17: Simulated steady-state epithelial lining fluid concentration-time profiles for clarithromycin across 10,000 virtual patients receiving 500 mg q12h.

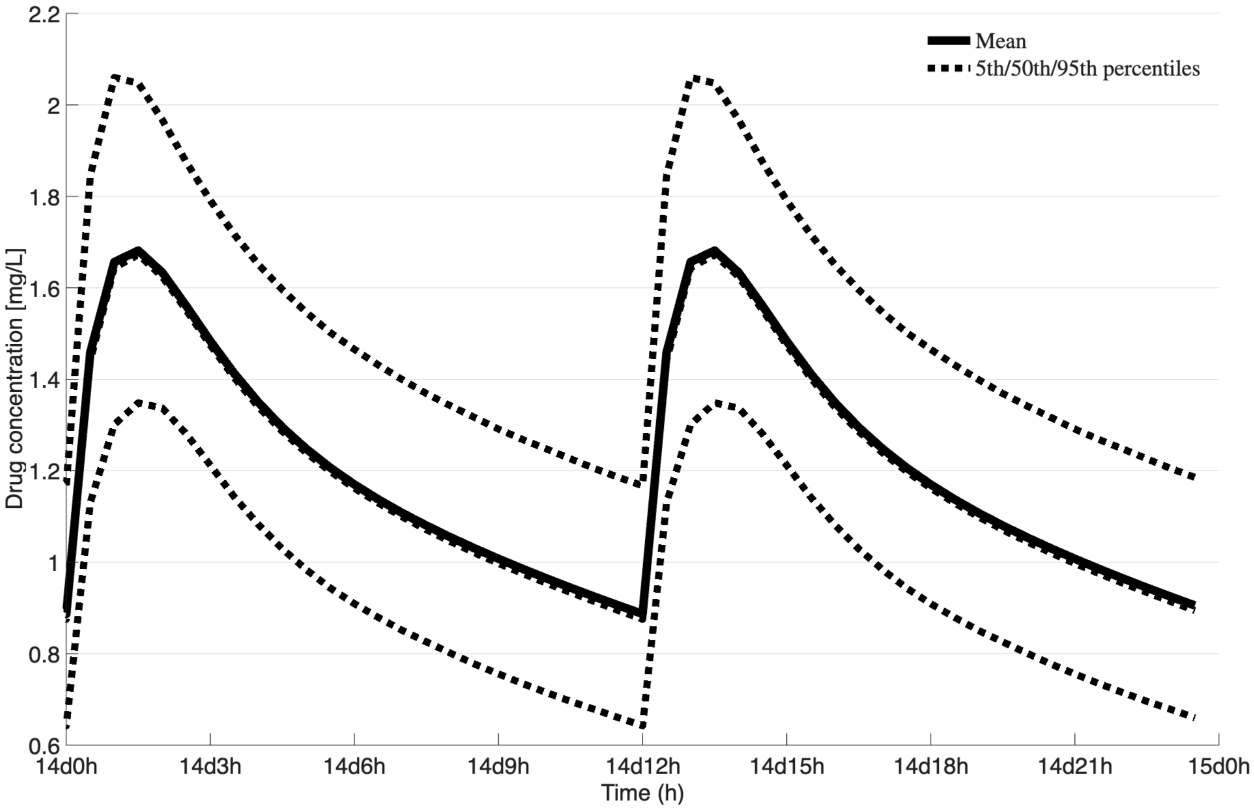

Figure S18: Simulated steady-state central compartment concentration-time profiles for clarithromycin showing the mean and 90% prediction interval across 10,000 virtual patients receiving 500 mg q12h.

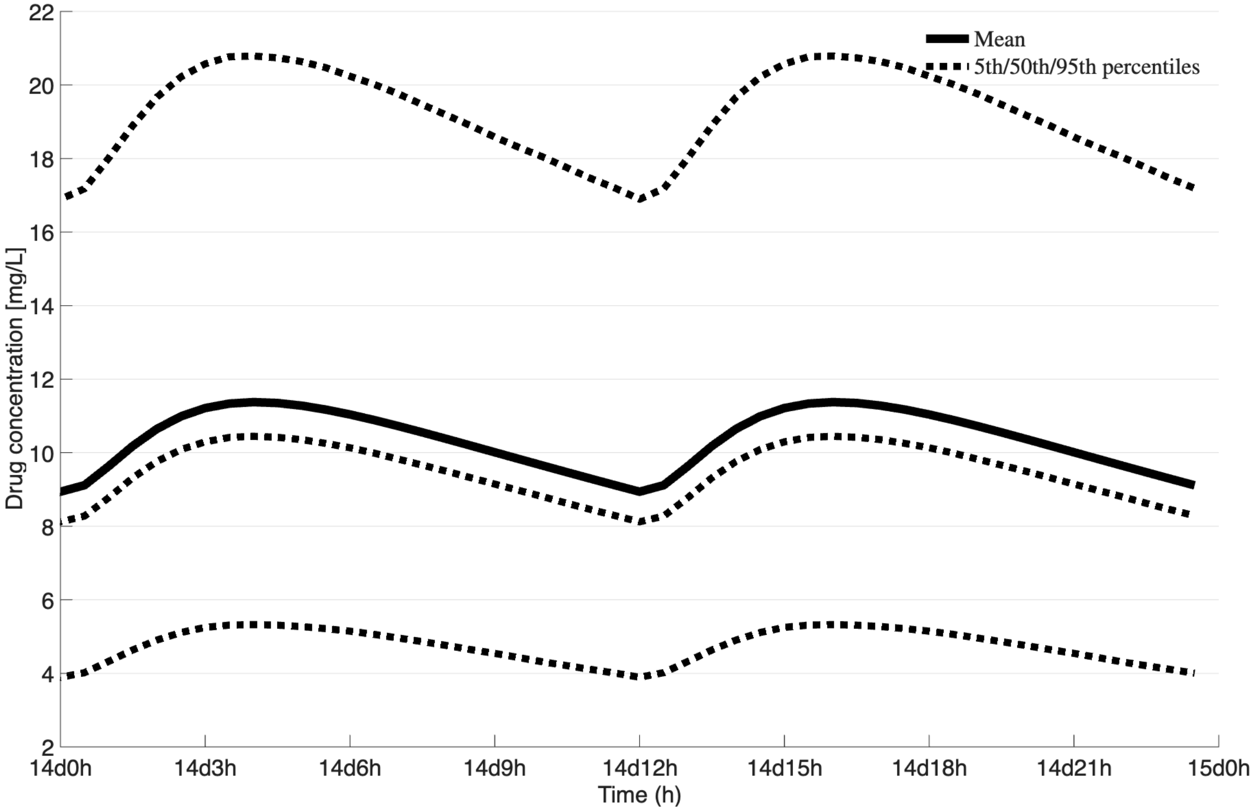

Figure S19: Simulated steady-state epithelial lining fluid concentration-time profiles for clarithromycin showing the mean and 90% prediction interval across 10,000 virtual patients receiving 500 mg q12h.

### G Ethambutol

#### G.1 Ethambutol model parameters

Table S6: Population pharmacokinetic parameters for the transit-compartment ethambutol model. Values represent typical population means; IIV and IOV are expressed as percent coefficient of variation ($\%CV$).

| **Parameter** | **Mean** | **IIV % CV** | **IOV % CV** | **Units** |
|---|---|---|---|---|
| Vc_F | 82.4 | – | – | L |
| Vp_F | 623 | – | – | L |
| CL_F | 39.9 | 20 | 36 | L/h |
| Q_F | 34.3 | – | – | L/h |
| k_a | 0.474 | 39 | – | 1/h |
| MTT | 0.789 | 93 | – | h |
| HIV | -0.154 | – | – | – |

Vc_F is the volume of distribution for the central compartment. Vp_F is the volume of distribution for the peripheral compartment. CL_F is the clearance rate for the antibiotic from the central compartment. Q_F is the inter-compartmental clearance rate between the central and peripheral compartment in both directions. k_a is the absorption rate from the absorption compartment to the central compartment. MTT is the mean transit time between transit compartments. HIV is the parameter that affects the bio-absorption of the antibiotic in the system. For this parameter, if the patient is randomly sampled in the model to have HIV, then the value that HIV is multiplied by in Equation 22 is 1, otherwise, it is 0.

#### G.2 Ethambutol model equations

This model was recreated from the model proposed in [24]. The parameter values used in the patient simulations were calculated using Equation 4. The model structure consists of a transit-compartment absorption input, a central compartment ($C$), and a peripheral compartment ($C_p$), with an HIV covariate effect on bioavailability.

$$\texttt{F} = 1 \cdot (1 - \texttt{HIV} \cdot (1|0)) \tag{22}$$

$$\texttt{k\_tr} = \frac{\texttt{n} + 1}{\texttt{MTT}} \tag{23}$$

$$\texttt{q0\_frac} = \Big(\texttt{k\_tr} \cdot t\Big)^{\texttt{n}} \cdot \Big(\exp\left(-\texttt{k\_tr} \cdot t\right)\Big)/\Gamma_{\texttt{n}} \tag{24}$$

$$\frac{d(q)}{dt} = \texttt{F} \cdot \texttt{dose} \cdot \texttt{k\_tr} \cdot \texttt{q0\_frac} \tag{25}$$

Where k_tr was calculated using Equation 23, and q0_frac was calculated using Equation 24. n was set to 1 to represent 1 transit compartment.

$$\frac{d(C)}{dt} = \texttt{k\_a} \cdot q - \left(\texttt{Q\_F} \cdot \frac{C}{\texttt{Vc\_F}}\right) + \left(\texttt{Q\_F} \cdot \frac{C_p}{\texttt{Vp\_F}}\right) - \left(\texttt{CL\_F} \cdot \frac{C}{\texttt{Vc\_F}}\right) \tag{26}$$

$$\frac{d(C_p)}{dt} = \texttt{Q\_F} \cdot \frac{C}{\texttt{Vc\_F}} - \left(\texttt{Q\_F} \cdot \frac{C_p}{\texttt{Vp\_F}}\right) \tag{27}$$

#### G.3 Ethambutol model flowchart

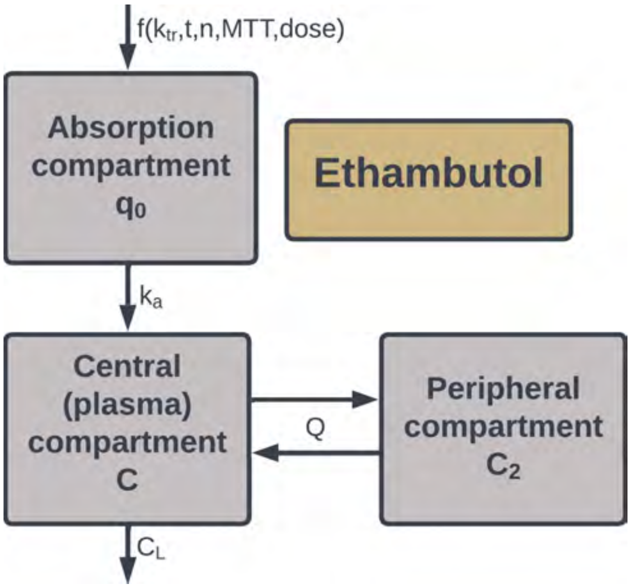

Figure S20: Schematic of the two-compartment population pharmacokinetic model for ethambutol with transit-compartment absorption, showing the absorption input ($q$), central compartment ($C$), and peripheral compartment ($C_p$), with systemic clearance (`CL_F`), intercompartmental clearance (`Q_F`), and HIV covariate effect on bioavailability (`F`).

##### G.4 Ethambutol model AUC distributions

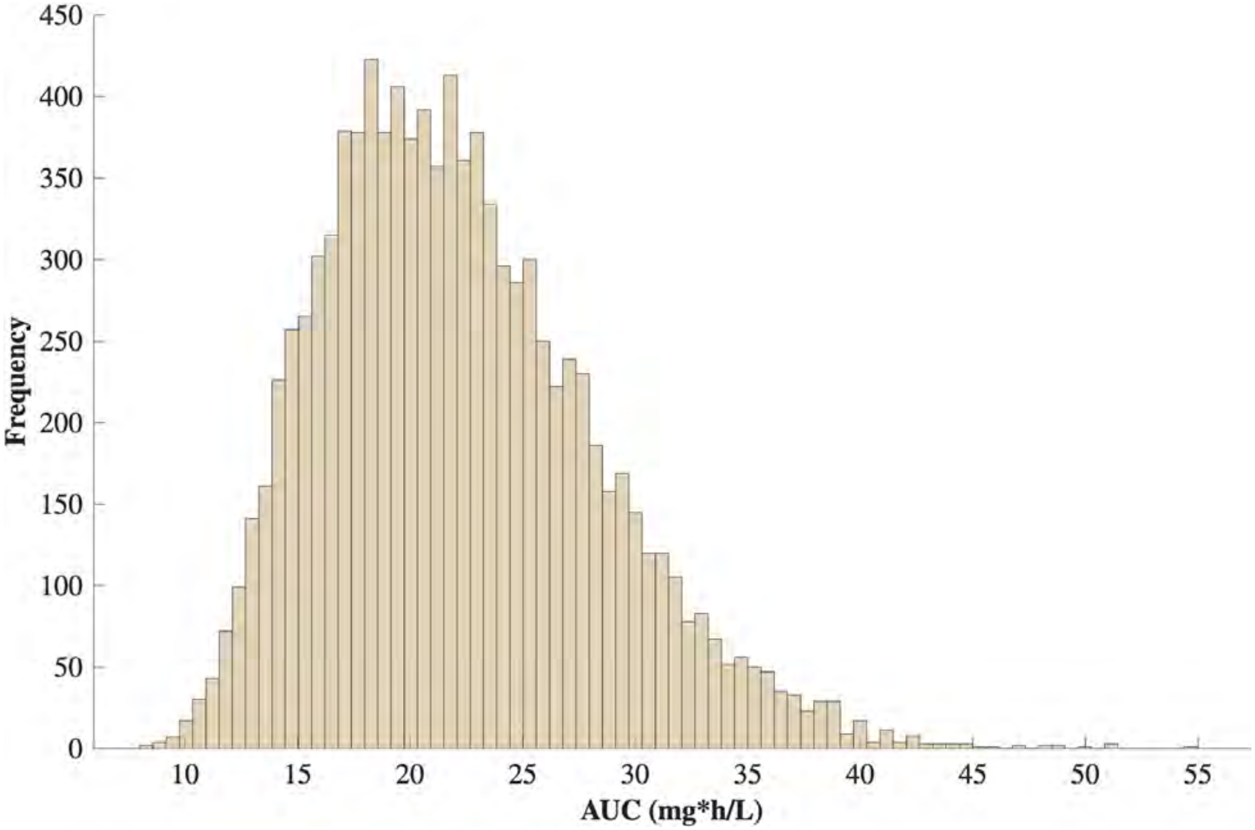

Figure S21: Steady-state $AUC_{0-24}$ distributions across 10,000 simulated patient profiles for ethambutol at 15 mg/kg q24h. Distributions reflect interindividual variability drawn from the parameter distributions in Table S6.

#### G.5 Ethambutol model concentration time-courses

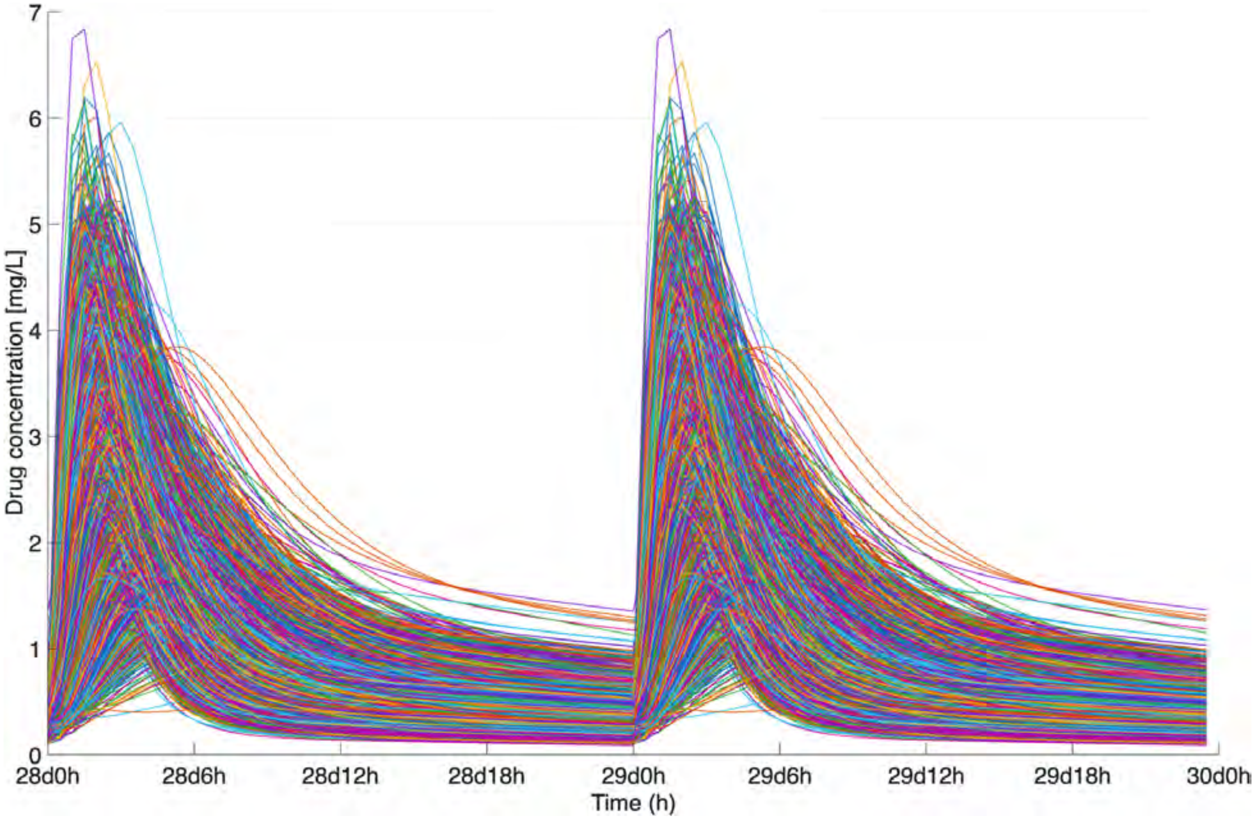

Figure S22: Simulated steady-state central compartment concentration-time profiles for ethambutol across 10,000 virtual patients at 15 mg/kg q24h.

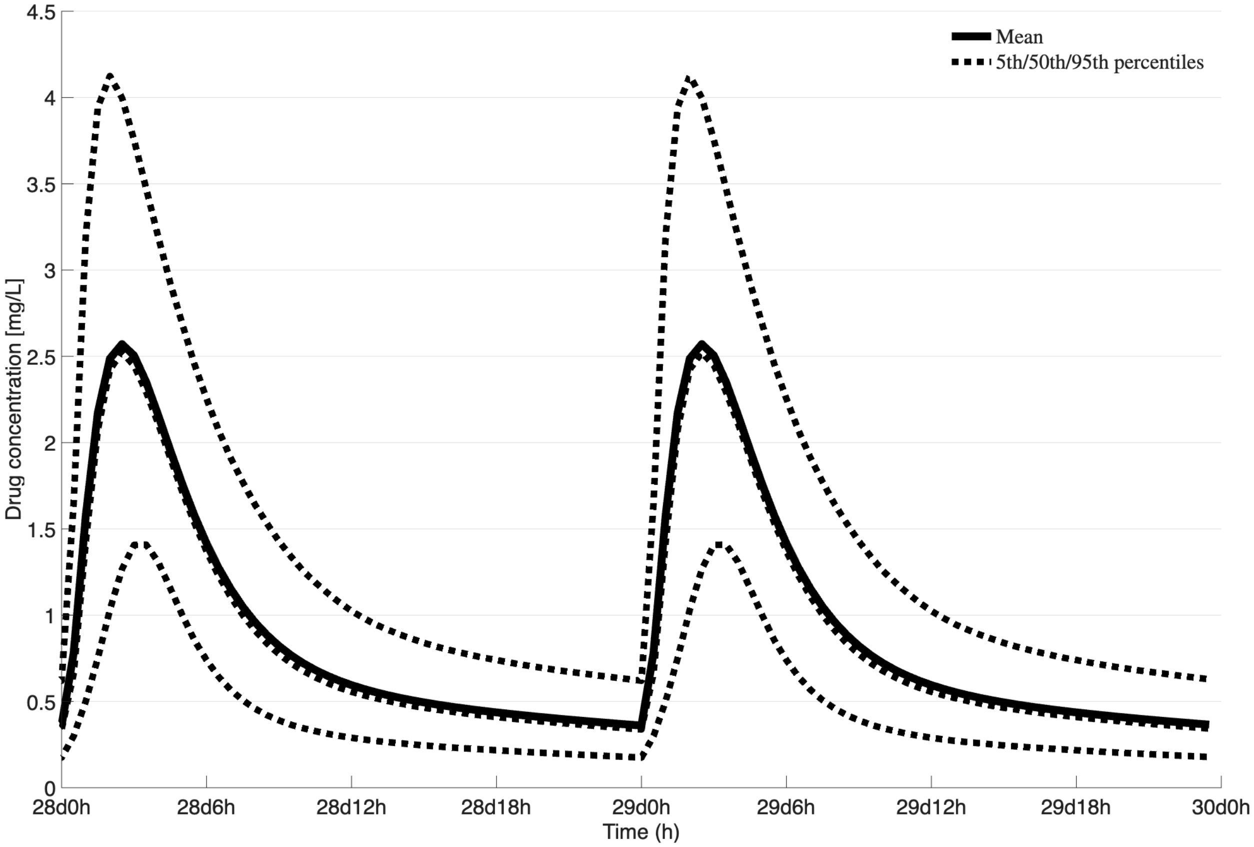

Figure S23: Simulated steady-state central compartment concentration-time profiles for ethambutol showing the mean and 90% prediction interval across 10,000 virtual patients at 15 mg/kg q24h.

### H Cefoxitin

#### H.1 Cefoxitin model parameters

Table S7: Population pharmacokinetic parameters for the one-compartment cefoxitin model. Values are the typical population means reported in the original source [32]; interindividual variability (IIV) is expressed as percent coefficient of variation ($\%CV$).

| Parameter | Mean | IIV % CV | Units |
|---|---|---|---|
| V_c | 23.4 | 44.8 | L |
| CL | 10.9 | 56.3 | L/h |

The cefoxitin model parameters are presented in Table S7 as they appear in the reimplemented model. The structural parameters are the systemic clearance (CL) and the central volume of distribution (V_c). In the original analysis, none of the tested covariates—including body weight, body composition, serum creatinine, and the various estimates of glomerular filtration rate—significantly improved the model [32]; consequently, no covariate scaling is applied to the parameters here. For each simulated patient, V_c and CL are drawn independently from log-normal distributions about their population means according to the IIV model described in Equation 4.

#### H.2 Cefoxitin model equations

This model was recreated from the one-compartment model proposed in [32]. The model structure consists of a single central compartment ($C$) with first-order elimination, with drug administered as a short IV infusion described by Equation 1. The parameter values used in the patient simulations were generated using the interindividual variability model in Equation 4.

$$\frac{d(A)}{dt} = \texttt{infusion rate} - \texttt{CL} \cdot \frac{A}{\texttt{V\_c}} \tag{28}$$

$A$ is the amount of cefoxitin in the central compartment.

$$C = \frac{A}{\texttt{V\_c}} \tag{29}$$

The reported plasma concentration $C$ is obtained by dividing the central-compartment amount by the patient-specific volume of distribution.

##### H.3 Cefoxitin model flowchart

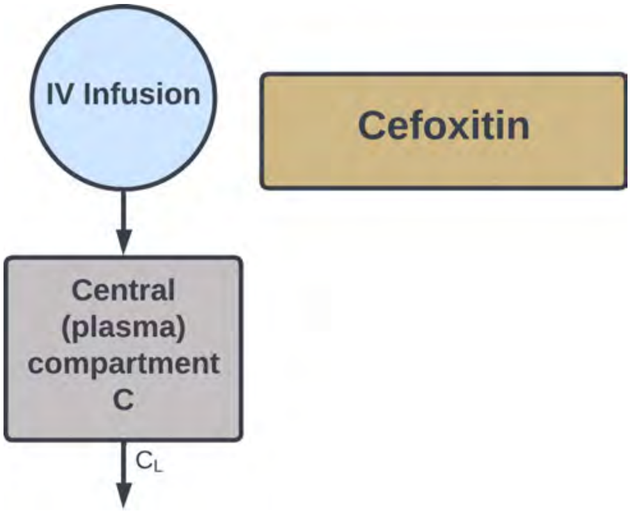

Figure S24: Schematic of the one-compartment population pharmacokinetic model for cefoxitin, showing the central compartment ($C$) with systemic clearance (CL) and a short IV infusion input. Drug is eliminated from the central compartment by a first-order process.

#### H.4 Cefoxitin model AUC distributions

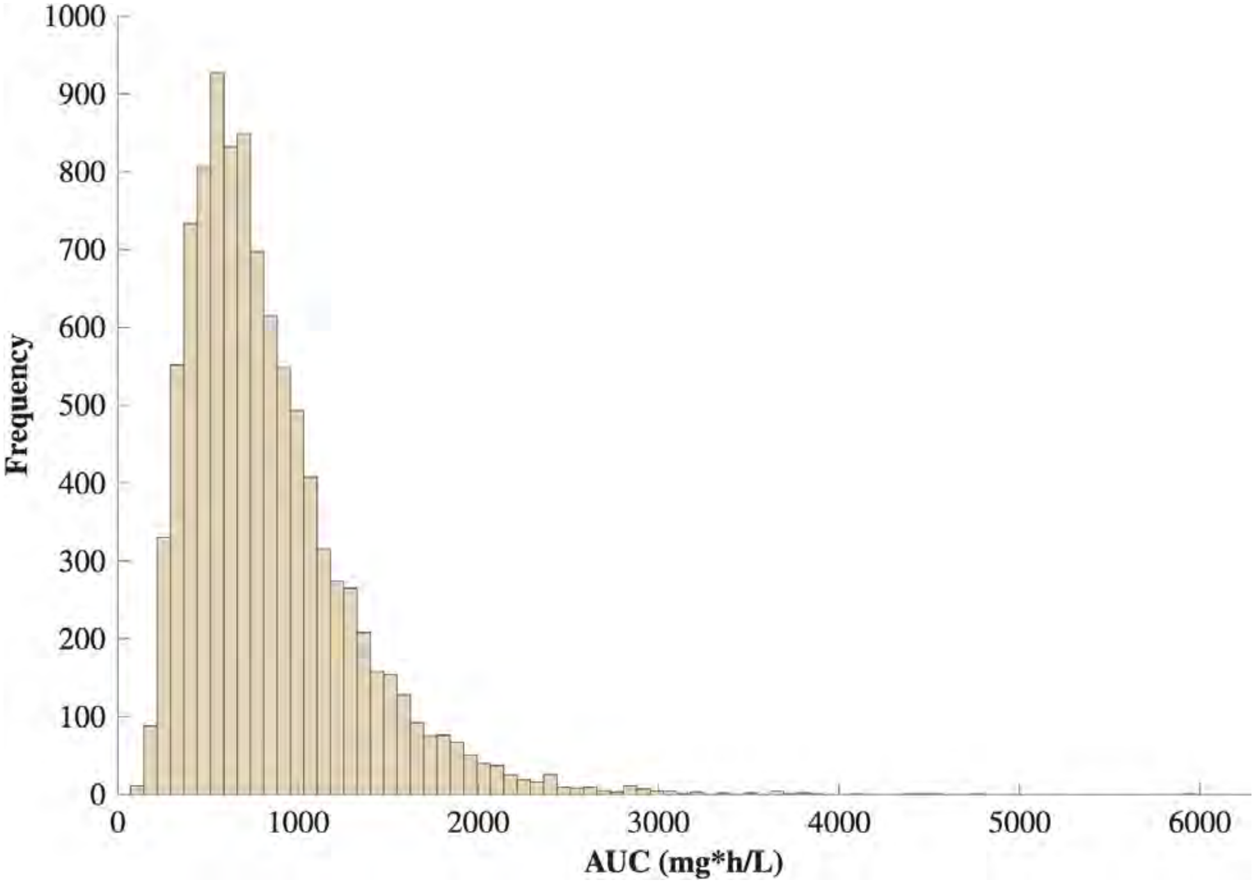

Figure S25: Steady-state $AUC_{0-24}$ distribution across the simulated virtual-patient population for cefoxitin at 4 g q12h administered as a 3-hour IV infusion. The distribution reflects interindividual variability in `V_c` and `CL` drawn from the parameters in Table S7.

#### H.5 Cefoxitin model concentration time-courses

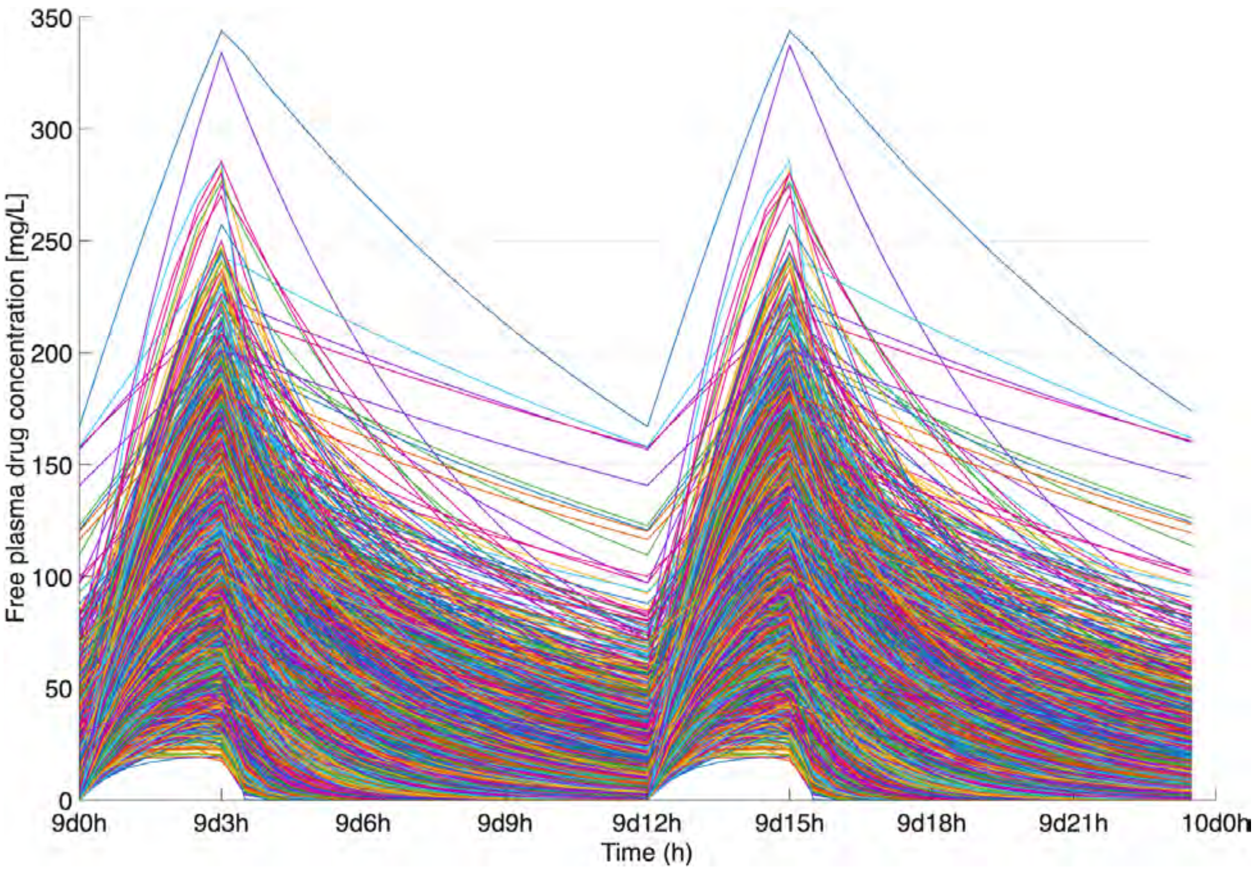

Figure S26: Simulated steady-state central-compartment concentration-time profiles for cefoxitin across the virtual-patient population receiving 4 g q12h as a 3-hour IV infusion.

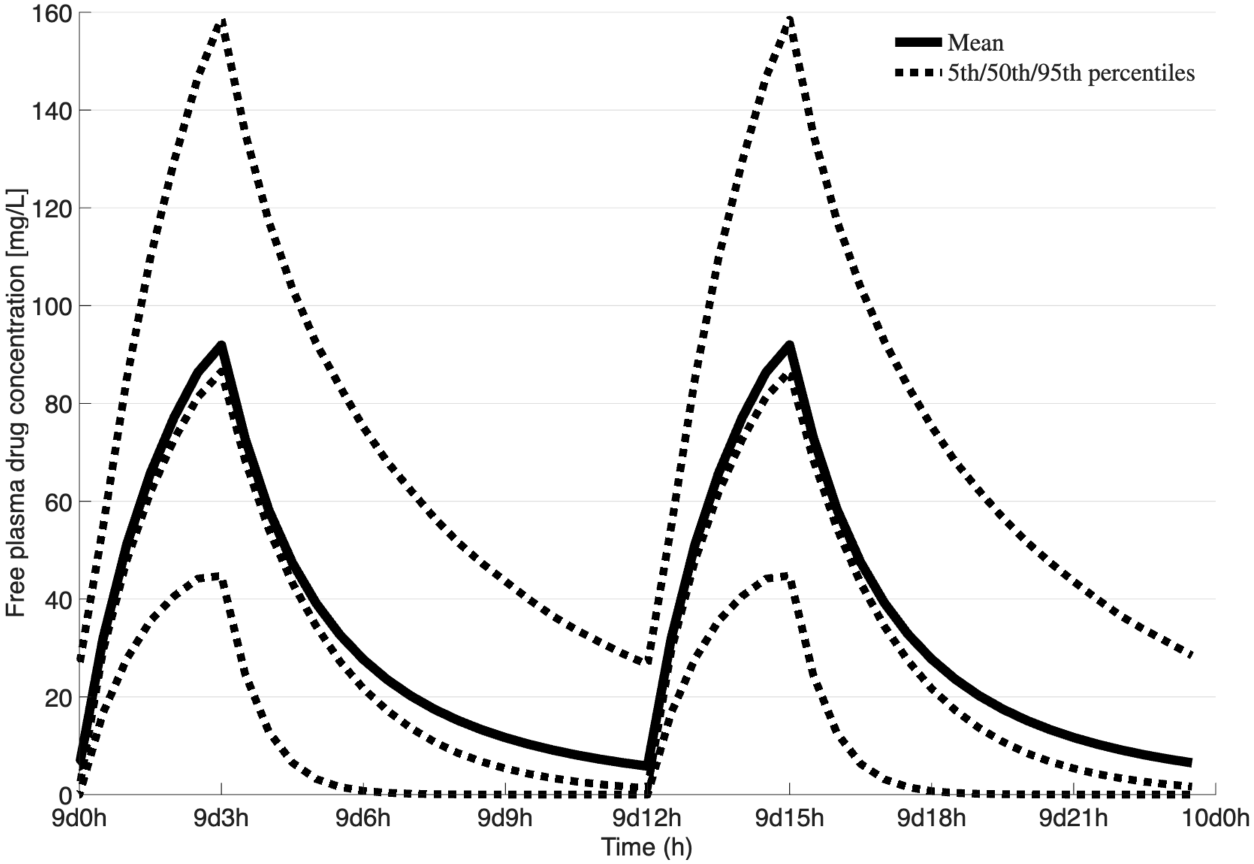

Figure S27: Simulated steady-state central-compartment concentration-time profiles for cefoxitin showing the median (50th percentile) and the 5th–95th percentile (90%) prediction interval across the virtual-patient population receiving 4 g q12h as a 3-hour IV infusion.

### I Imipenem

#### I.1 Imipenem model parameters

Table S8: Population pharmacokinetic parameters for the two-compartment imipenem model. Values represent typical population means; IIV is expressed as percent coefficient of variation ($\%CV$).

| Parameter | Mean | IIV % CV | Units |
|---|---|---|---|
| Vc_F | 20.5 | 14.8 | L |
| Vp_F | 8.86 | – | L |
| CL_F | 8.88 | 17.7 | L/h |
| Q_F | 1.74 | – | L/h |

All parameters for the reimplementation of the imipenem model are presented in Table S8 as they appear in the model from [9]. These parameters are used in the following equations to generate the pharmacokinetic profiles seen in Figure S30 and used to calculate the AUC distribution in Figure S29.

#### I.2 Imipenem model equations

This model was recreated from the model proposed in [9]. The parameter values used in the patient simulations were calculated using Equation 4. The model structure consists of a central compartment ($C$) and a peripheral compartment ($C_p$), with drug administered as a 3-hour IV infusion as described by Equation 1.

$$\frac{d(C)}{dt} = \texttt{infusion rate} - \texttt{Q\_F} \cdot \frac{C}{\texttt{Vc\_F}} + \left(\texttt{Q\_F} \cdot \frac{C_p}{\texttt{Vc\_F}}\right) - \left(\texttt{CL\_F} \cdot \frac{C}{\texttt{Vc\_F}}\right) \tag{30}$$

$$\frac{d(C_p)}{dt} = \texttt{Q\_F} \cdot \frac{C}{\texttt{Vp\_F}} - \left(\texttt{Q\_F} \cdot \frac{C_p}{\texttt{Vp\_F}}\right) \tag{31}$$

#### I.3 Imipenem model flowchart

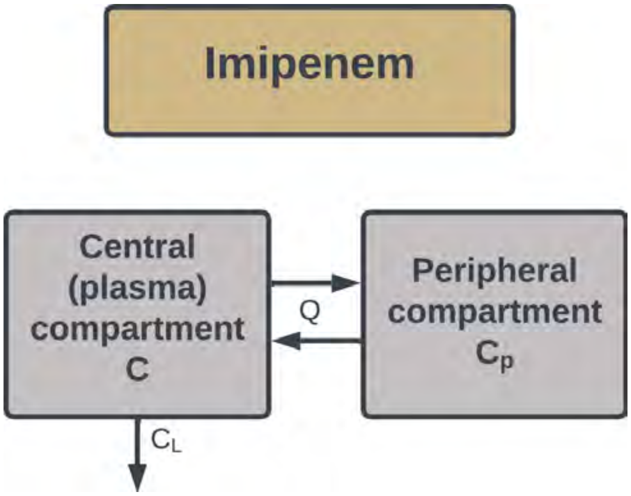

Figure S28: Schematic of the two-compartment population pharmacokinetic model for imipenem, showing the central compartment ($C$) and peripheral compartment ($C_p$), with systemic clearance (CL_F), intercompartmental clearance (Q_F), and 3-hour IV infusion input.

#### I.4 Imipenem model AUC distributions

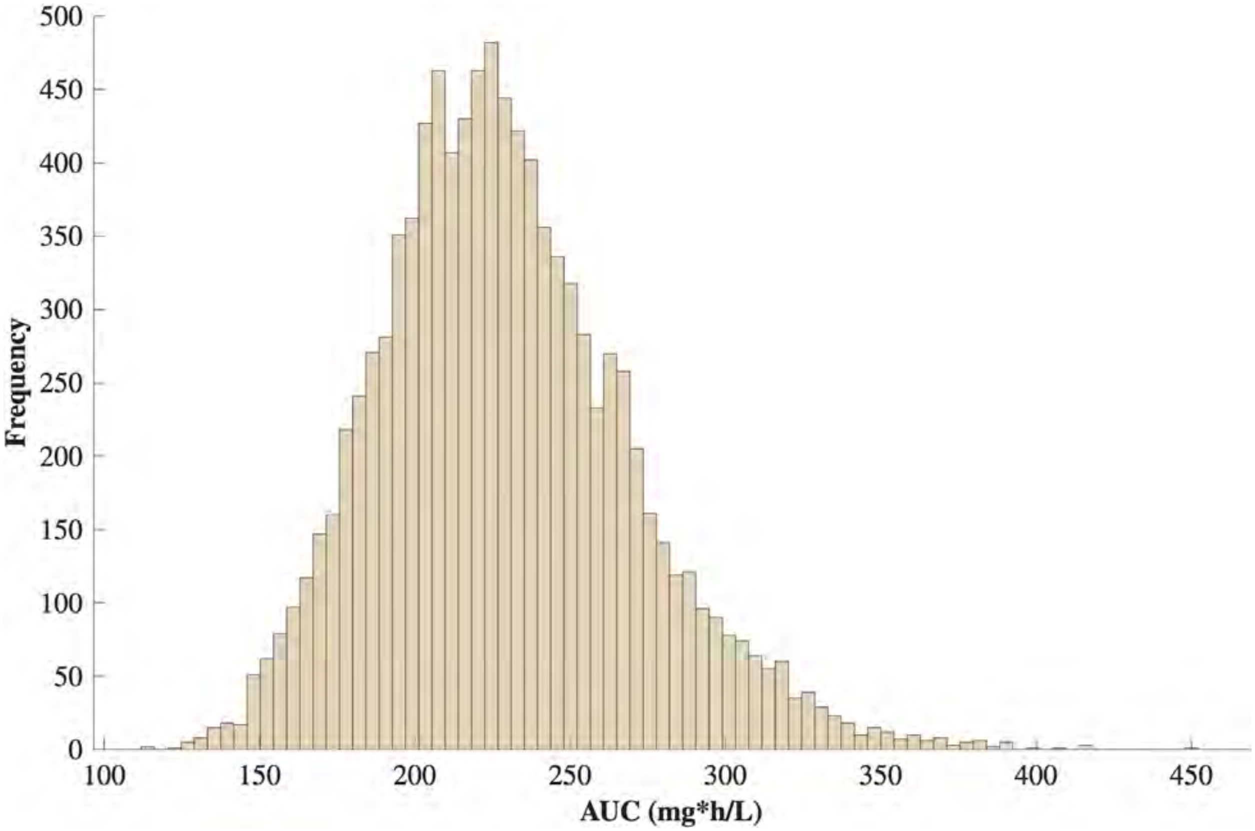

Figure S29: Steady-state $AUC_{0-24}$ distributions across 10,000 simulated patient profiles for imipenem at 1000 mg q12h administered as a 3-hour IV infusion. Distributions reflect interindividual variability drawn from the parameter distributions in Table S8.

#### I.5 Imipenem model concentration time-courses

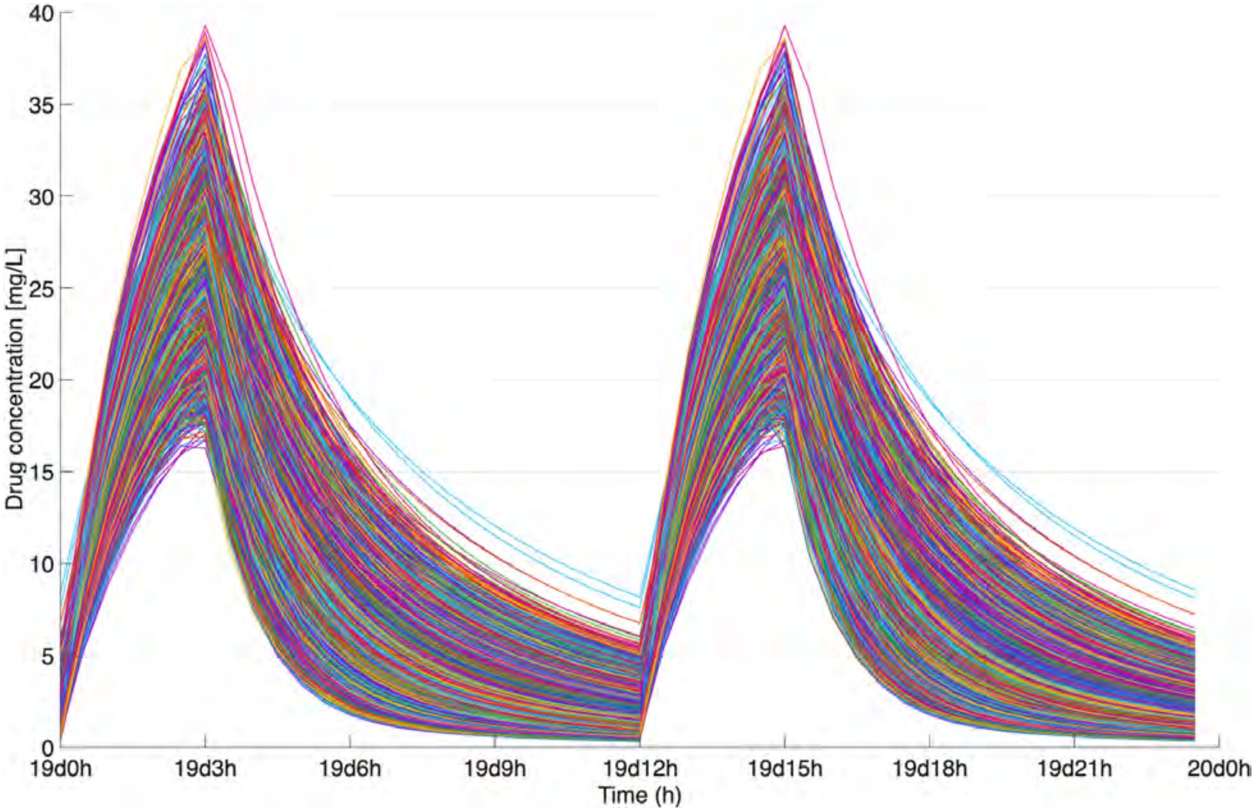

Figure S30: Simulated steady-state central compartment concentration-time profiles for imipenem across 10,000 virtual patients receiving 1000 mg q12h as a 3-hour IV infusion.

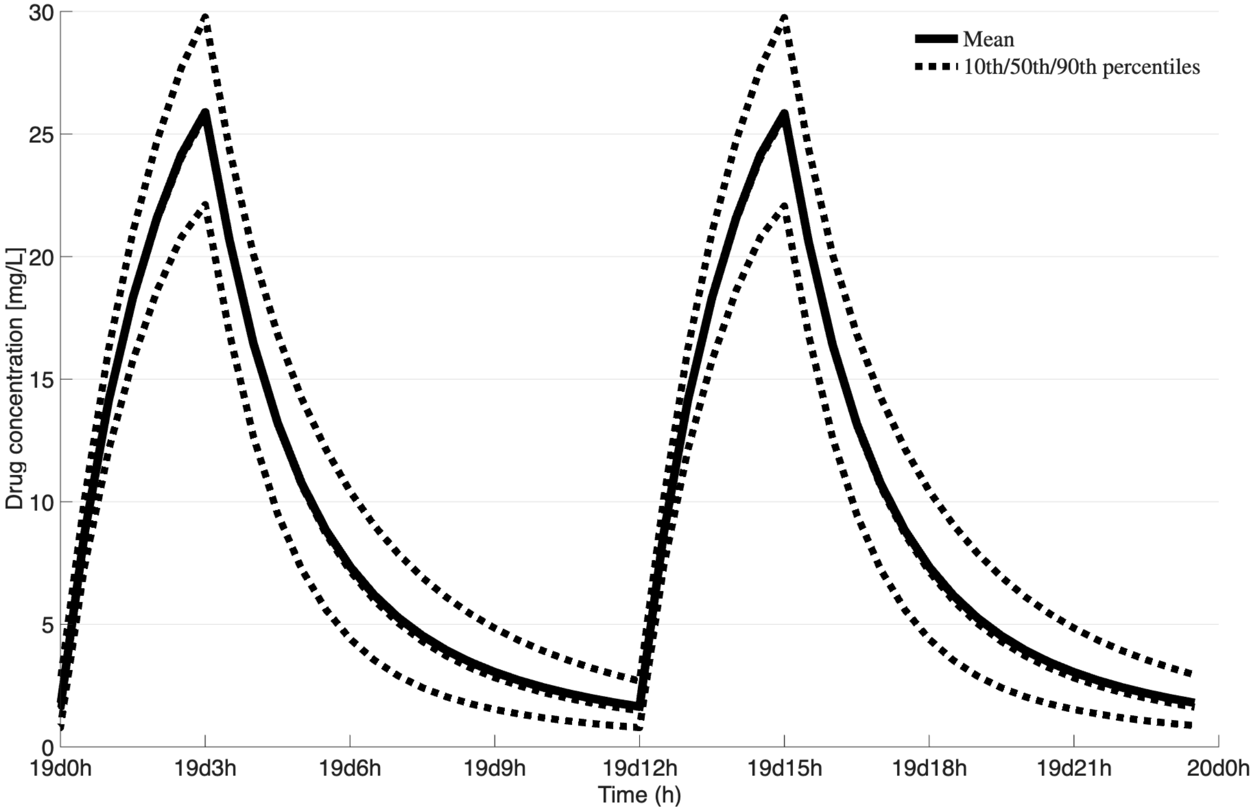

Figure S31: Simulated steady-state central compartment concentration-time profiles for imipenem showing the mean and 90% prediction interval across 10,000 virtual patients receiving 1000 mg q12h as a 3-hour IV infusion.

### J Linezolid

#### J.1 Linezolid model parameters

Table S9: Population pharmacokinetic parameters for the transit-compartment linezolid model. Values represent typical population means; IIV and IOV are expressed as percent coefficient of variation ($\%CV$).

| Parameter | Mean | IIV % CV | IOV % CV | Units |
|---|---|---|---|---|
| V_C | 40.2 | – | – | L |
| MTT | 0.528 | – | 56.8 | h |
| n | 5 | – | – | – |
| K_a | 1.22 | – | 78.5 | 1/h |
| F | 1 | – | 22.0 | – |
| CL | 3.57 | 37.1 | – | L/h |

V_C is volume in central compartment; MTT is the absorption mean transit time; n is the number of transit compartments; K_a is the absorption rate constant; F is the bioavailability; CL is the clearance.

#### J.2 Linezolid model equations

This model was recreated from the model proposed in [3]. The parameter values used in the patient simulations were calculated using Equation 4. The model structure was an eight (8) compartment model with an absorption compartment ($q_0$), five (5) transit compartments ($q_1$–$q_5$), a final pre-absorption compartment ($q$), and a central compartment ($C$).

$$\frac{d(q_0)}{dt} = -\texttt{k\_tr} \cdot q_0 \tag{32}$$

$$\frac{d(q_i)}{dt} = \texttt{k\_tr}\Big(q_{i-1} - q_i\Big), \quad i = 1, \ldots, 5 \tag{33}$$

$$\frac{d(q)}{dt} = \texttt{k\_tr} \cdot q_5 - \texttt{K\_a} \cdot q \tag{34}$$

$$\frac{d(C)}{dt} = \texttt{F} \cdot \texttt{K\_a} \cdot q - C \cdot \left(\frac{\texttt{CL}}{\texttt{V\_C}}\right) \tag{35}$$

##### J.3 Linezolid model flowchart

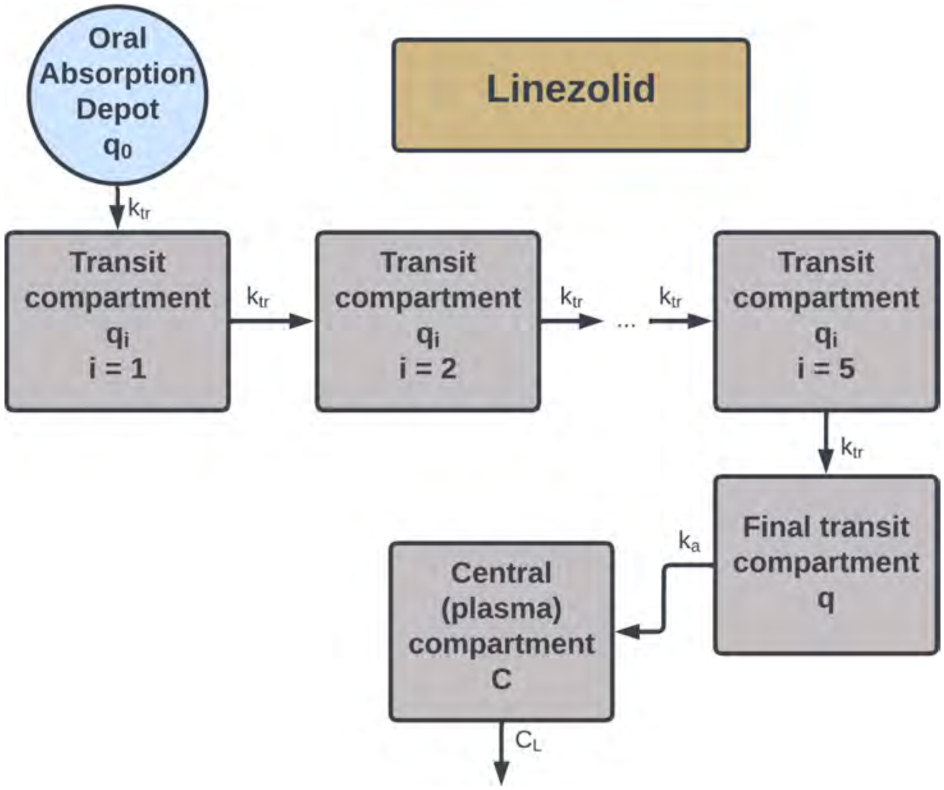

Figure S32: Schematic of the eight-compartment population pharmacokinetic model for linezolid, showing the absorption compartment ($q_0$), five transit compartments ($q_1$–$q_5$), final pre-absorption compartment ($q$), and central compartment ($C$), with absorption rate constant (`K_a`), transit rate constant (`k_tr`), bioavailability (`F`), and systemic clearance (`CL`).

#### J.4 Linezolid model AUC distributions

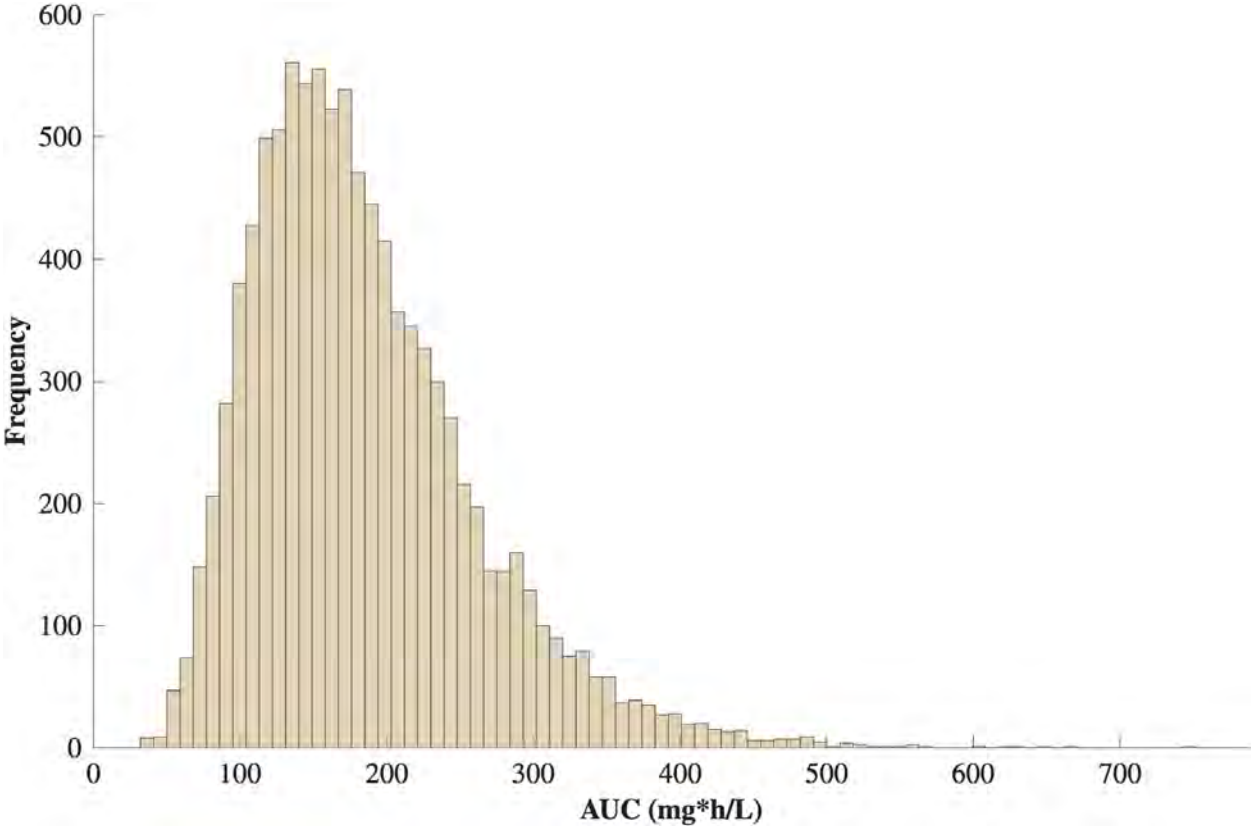

Figure S33: Steady-state $AUC_{0-24}$ distributions across 10,000 simulated patient profiles for linezolid at 600 mg q24h. Distributions reflect interindividual variability drawn from the parameter distributions in Table S9.

##### J.5 Linezolid model concentration time-courses

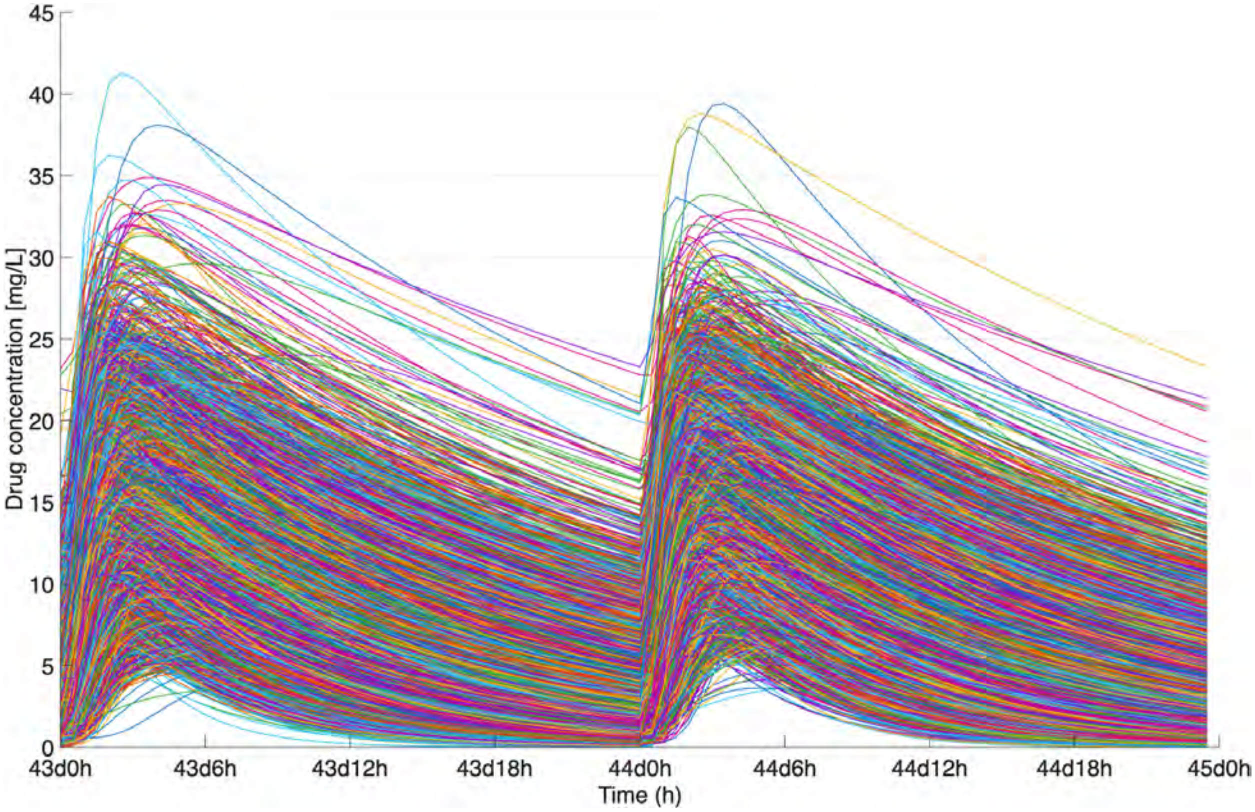

Figure S34: Simulated steady-state central compartment concentration-time profiles for linezolid across 10,000 virtual patients receiving 600 mg q24h.

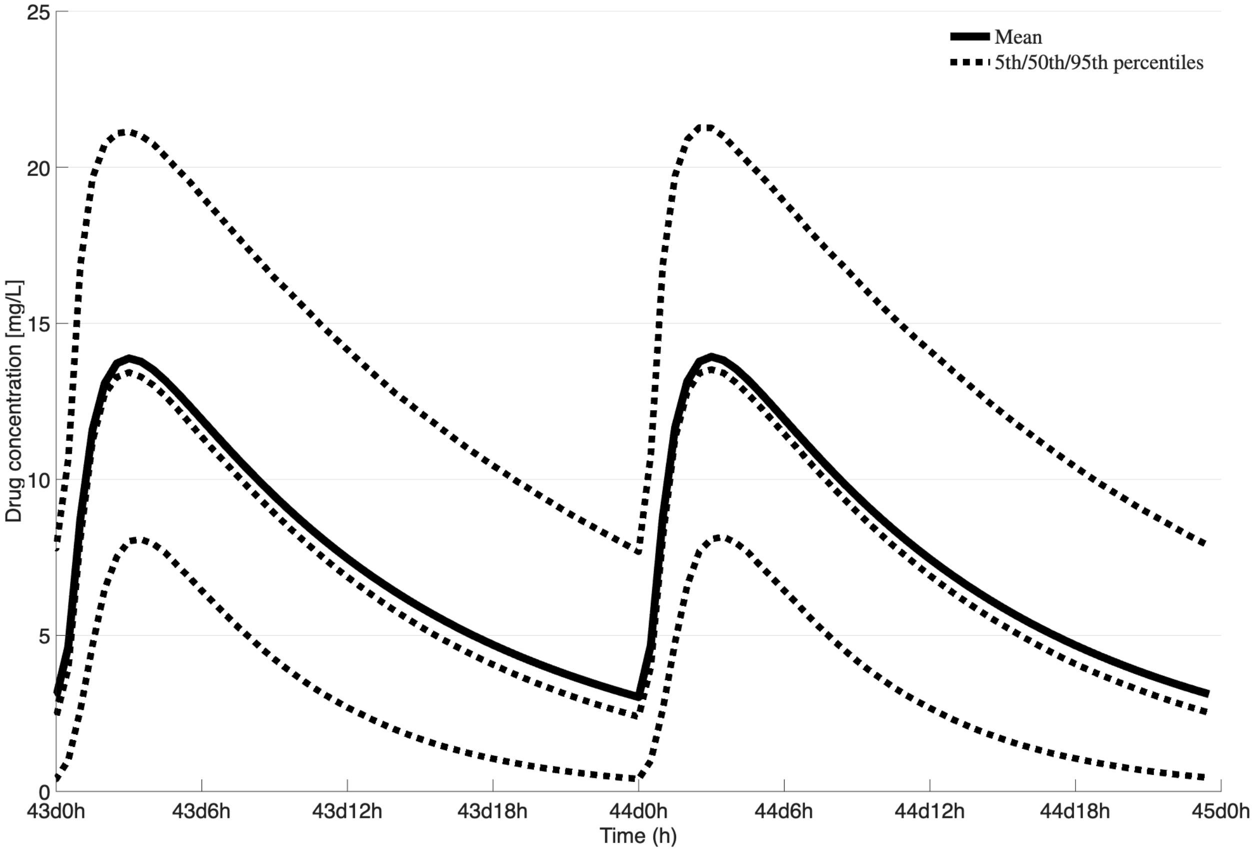

Figure S35: Simulated steady-state central compartment concentration-time profiles for linezolid showing the mean and 90% prediction interval across 10,000 virtual patients receiving 600 mg q24h.

### K Moxifloxacin

#### K.1 Moxifloxacin model parameters

Table S10: Population pharmacokinetic parameters for the one-compartment moxifloxacin model. Values represent typical population means; IIV is expressed as percent coefficient of variation ($\%CV$).

| Parameter | Mean | IIV % CV | Units |
|---|---|---|---|
| V_F | 110 | 38 | L |
| CL_F | 9.59 | 30 | L/h |
| ka | 2.69 | 196 | 1/h |

All parameters used for the reimplementation of the moxifloxacin model are presented in Table S10 as they appear in the model from [38]. These parameters are used in the following equations to generate the pharmacokinetic profiles seen in Figure S38 and used to calculate the AUC distributions in Figure S37.

#### K.2 Moxifloxacin model equations

This model was recreated from the model proposed in [38]. The parameter values used in the patient simulations were calculated using Equation 4. The model structure consists of an absorption compartment ($C_q$) and a central compartment ($C$).

$$\frac{d(C_q)}{dt} = -\texttt{ka} \cdot C_q \tag{36}$$

$$\frac{d(C)}{dt} = \texttt{ka} \cdot C_q - \left(\texttt{CL\_F} \cdot \frac{C}{\texttt{V\_F}}\right) \tag{37}$$

#### K.3 Moxifloxacin model flowchart

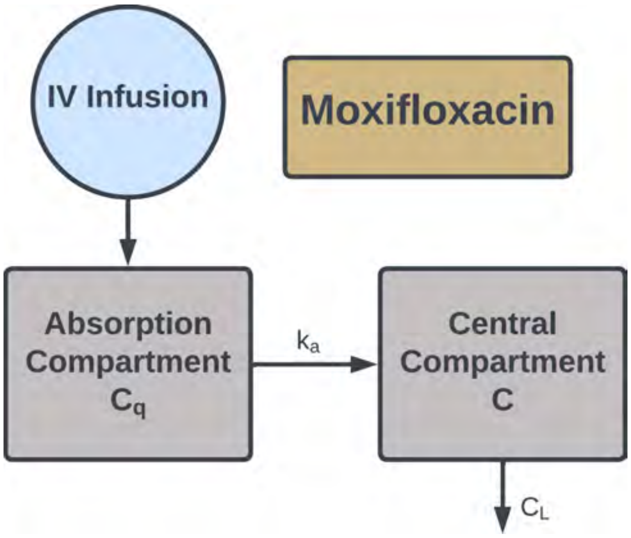

Figure S36: Schematic of the one-compartment population pharmacokinetic model for moxifloxacin, showing the absorption compartment ($C_q$) and central compartment ($C$), with first-order absorption rate constant (ka) and systemic clearance (CL_F).

#### K.4 Moxifloxacin model AUC distributions

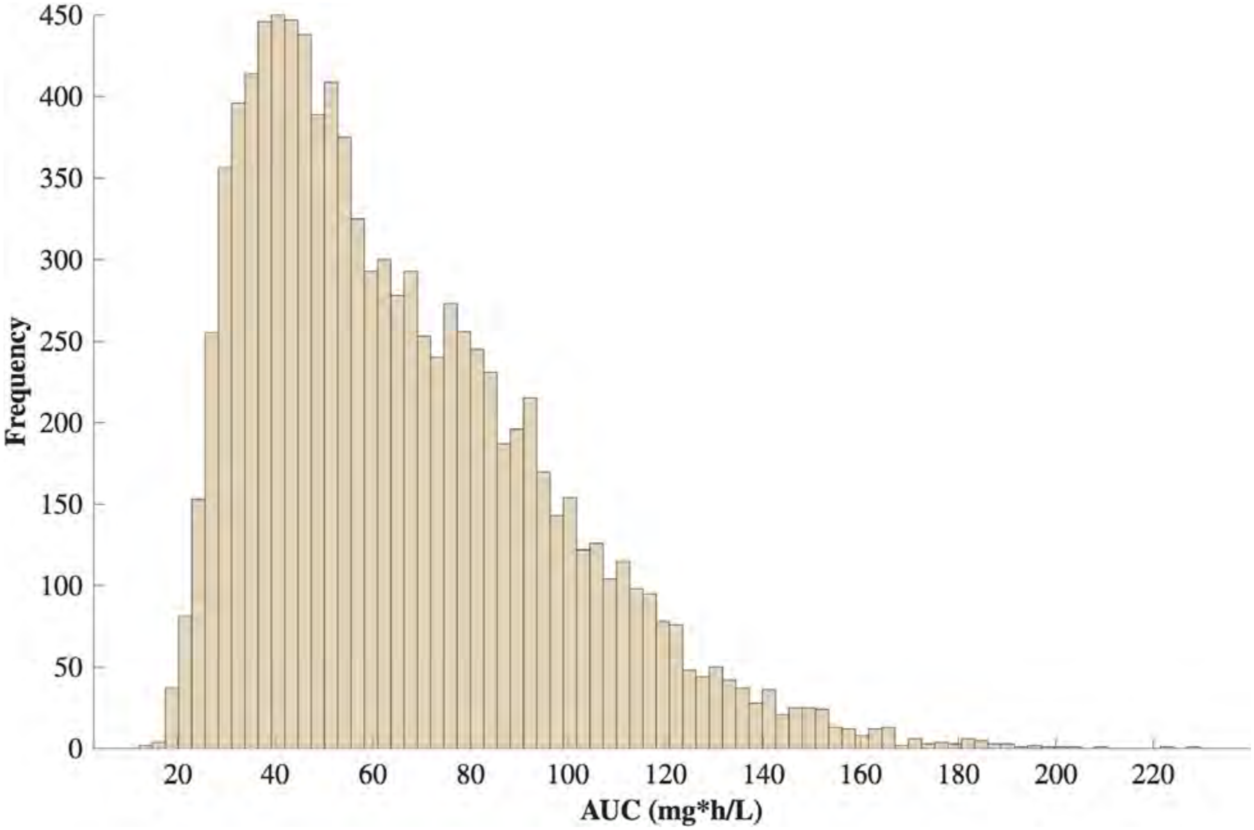

Figure S37: Steady-state $AUC_{0-24}$ distributions across 10,000 simulated patient profiles for moxifloxacin at 400 mg q24h and 800 mg q24h. Distributions reflect interindividual variability drawn from the parameter distributions in Table S10.

#### K.5 Moxifloxacin model concentration time-courses

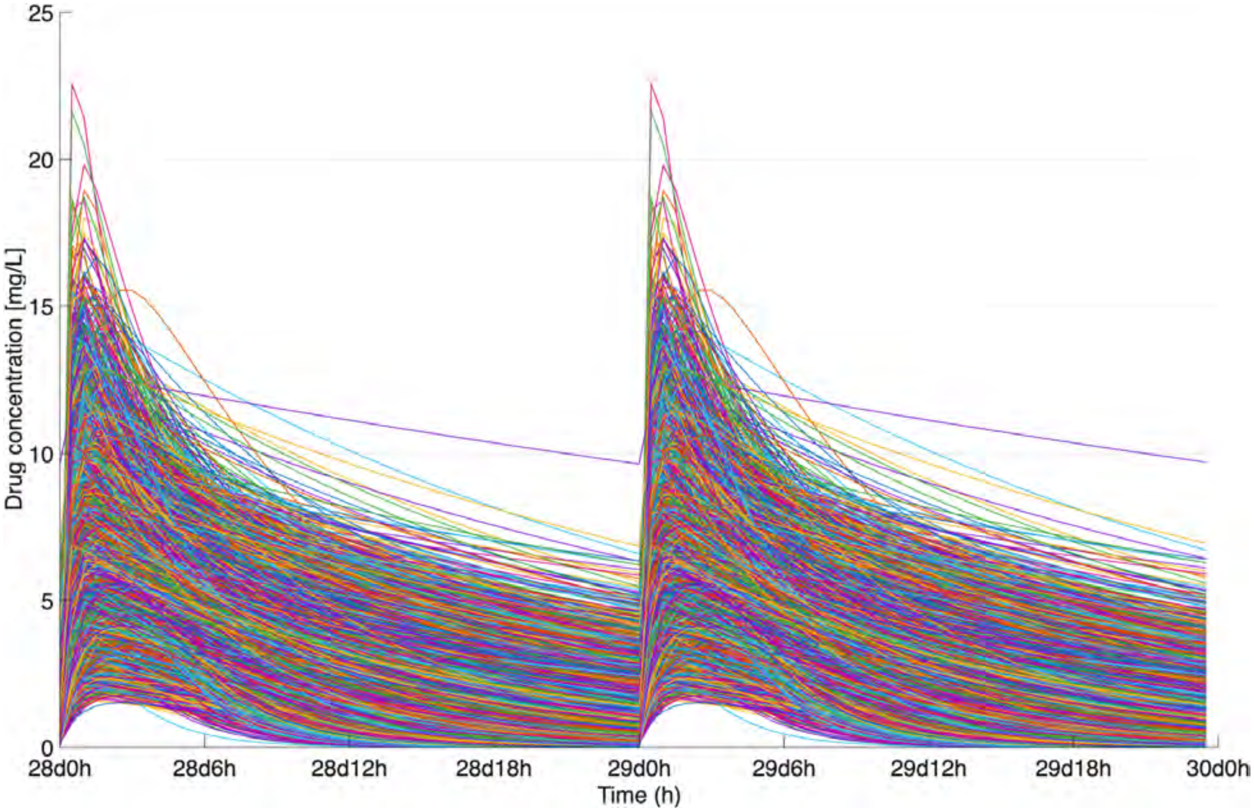

Figure S38: Simulated steady-state central compartment concentration-time profiles for moxifloxacin across 10,000 virtual patients receiving 400 mg q24h and 800 mg q24h.

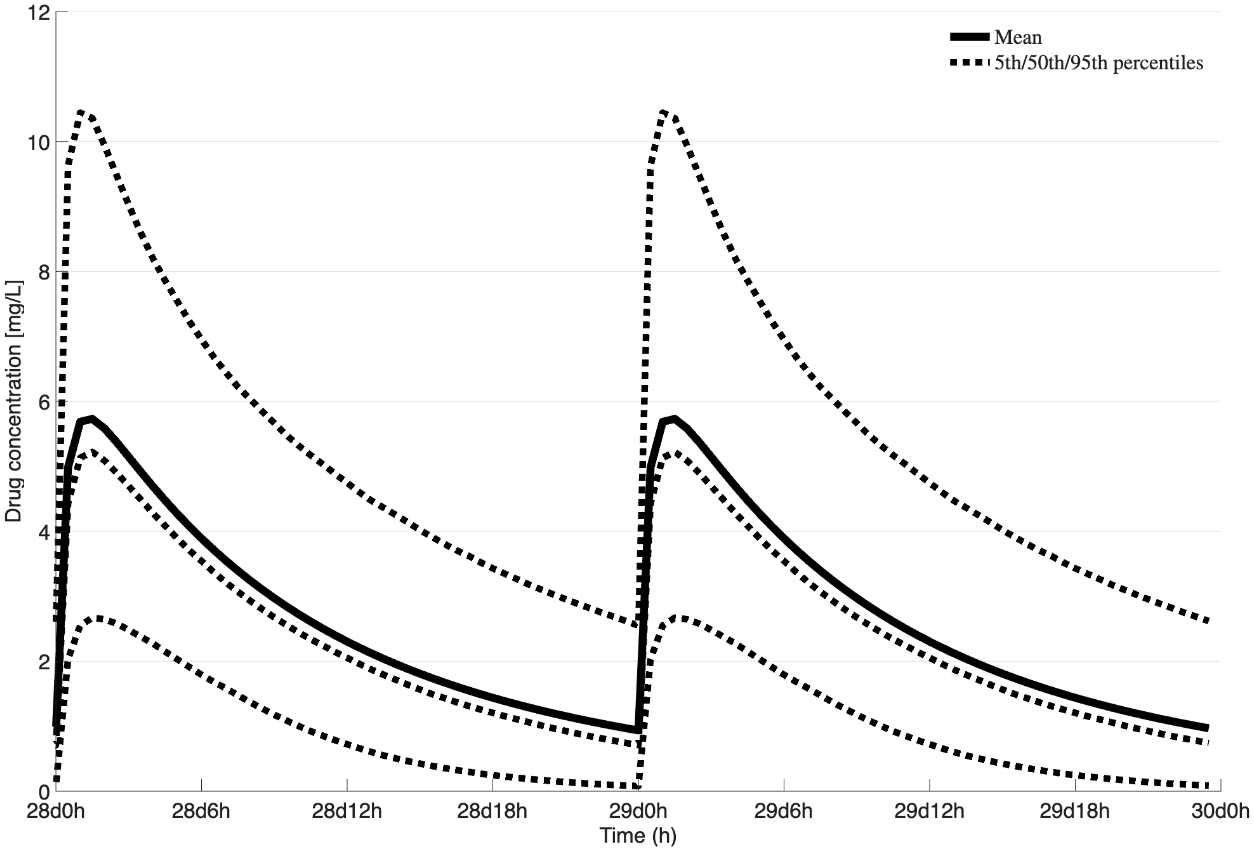

Figure S39: Simulated steady-state central compartment concentration-time profiles for moxifloxacin showing the mean and 90% prediction interval across 10,000 virtual patients receiving 400 mg q24h and 800 mg q24h.

### L Rifabutin

#### L.1 Rifabutin model parameters

Table S11: Population pharmacokinetic parameters for the transit-compartment rifabutin model with parallel des-rifabutin metabolite tracking. Values represent typical population means; IIV is expressed as percent coefficient of variation ($\%CV$).

| Parameter | Mean | IIV % CV | Units |
|---|---|---|---|
| MTT | 2.16 | 0.518 | h |
| n | 7.15 | – | – |
| k_tr | 3.77 | – | /h |
| k_a | 0.25 | 0.310 | /h |
| F_m | 0.09 | 0.893 | – |
| CL_1 | 58.80 | 0.540 | L/h |
| CL_2 | 122.00 | 0.783 | L/h |
| Q_1 | 62.21 | 0.602 | L/h |
| Q_2 | 71.80 | 0.608 | L/h |
| Q_3 | 5.76 | 0.515 | L/h |
| Vd_C,RFB | 6.55 | 1.691 | L |
| Vd_C,dRFB | 37.30 | 1.114 | L |
| Vd_P,RFB | 1580 | 0.656 | L |
| Vd_P,dRFB | 1220 | 0.268 | L |

MTT, mean transit time through transit compartments; n, number of transit compartments; k_tr, transit rate constant; k_a, absorption rate constant from the absorption compartment to the central compartment; F_m, first-pass metabolism fraction of rifabutin converted to des-rifabutin; CL_1, clearance rate of rifabutin from the central compartment; CL_2, clearance rate of des-rifabutin from the central compartment; Q_1, intercompartmental clearance between central and peripheral compartments for rifabutin; Q_2, intercompartmental clearance between central and peripheral compartments for des-rifabutin; Q_3, conversion clearance from rifabutin to des-rifabutin in the central compartment; Vd_C,RFB, volume of distribution for rifabutin in the central compartment; Vd_C,dRFB, volume of distribution for des-rifabutin in the central compartment; Vd_P,RFB, volume of distribution for rifabutin in the peripheral compartment; Vd_P,dRFB, volume of distribution for des-rifabutin in the peripheral compartment.

#### L.2 Rifabutin model equations

This model was recreated from the model proposed in [16]. The parameter values used in the patient simulations were calculated using Equation 4. The Stirling approximation for the gamma function is given by Equation 2 from the introduction. The model structure consists of a transit-compartment absorption input ($m_{abs}$), a rifabutin central compartment ($C_{C,RFB}$), a rifabutin peripheral compartment ($C_{P,RFB}$), a des-rifabutin central compartment ($C_{C,dRFB}$), and a des-rifabutin peripheral compartment ($C_{P,dRFB}$), with first-pass metabolism generating the des-rifabutin metabolite from both absorption and central compartment conversion.

$$\frac{d(m_{abs})}{dt} = \texttt{dose} \cdot \texttt{k\_tr} \cdot \frac{(\texttt{k\_tr} \cdot t)^n \cdot \exp(-\texttt{k\_tr} \cdot t)}{\Gamma_n} - \texttt{k\_a} \cdot m_{abs} \tag{38}$$

$$\frac{d(C_{C,RFB})}{dt} = \frac{\texttt{k\_a} \cdot (1 - \texttt{F\_m})}{\texttt{Vd\_C,RFB}} \cdot m_{abs} + \frac{-\texttt{Q\_1} - \texttt{CL\_1} - \texttt{Q\_3}}{\texttt{Vd\_C,RFB}} \cdot C_{C,RFB} + \frac{\texttt{Q\_1}}{\texttt{Vd\_C,RFB}} \cdot C_{P,RFB} \tag{39}$$

$$\frac{d(C_{C,dRFB})}{dt} = \frac{\texttt{k\_a} \cdot \texttt{F\_m}}{\texttt{Vd\_C,dRFB}} \cdot m_{abs} + \frac{\texttt{Q\_3}}{\texttt{Vd\_C,dRFB}} \cdot C_{C,RFB} + \frac{-\texttt{Q\_2} - \texttt{CL\_2}}{\texttt{Vd\_C,dRFB}} \cdot C_{C,dRFB} + \frac{\texttt{Q\_2}}{\texttt{Vd\_C,dRFB}} \cdot C_{P,dRFB} \tag{40}$$

$$\frac{d(C_{P,RFB})}{dt} = \frac{\texttt{Q\_1}}{\texttt{Vd\_P,RFB}} \cdot C_{C,RFB} - \frac{\texttt{Q\_1}}{\texttt{Vd\_P,RFB}} \cdot C_{P,RFB} \tag{41}$$

$$\frac{d(C_{P,dRFB})}{dt} = \frac{\texttt{Q\_2}}{\texttt{Vd\_P,dRFB}} \cdot C_{C,dRFB} - \frac{\texttt{Q\_2}}{\texttt{Vd\_P,dRFB}} \cdot C_{P,dRFB} \tag{42}$$

#### L.3 Rifabutin model flowchart

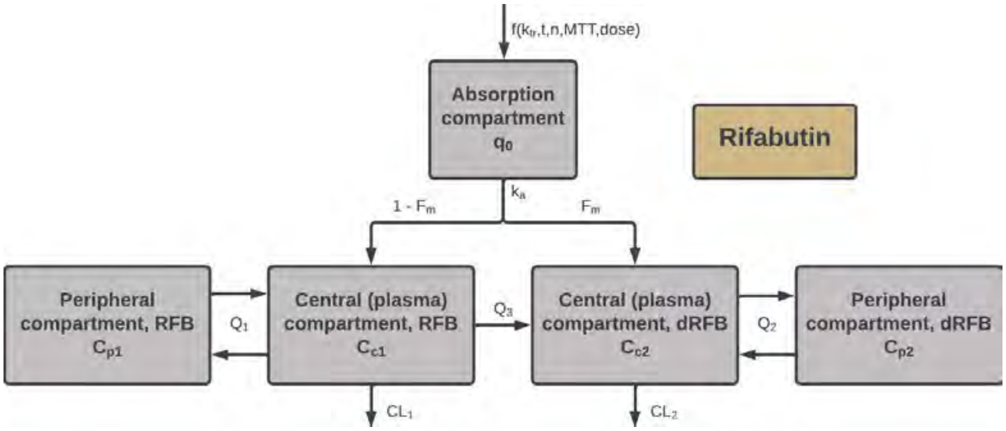

Figure S40: Schematic of the population pharmacokinetic model for rifabutin, showing the transit-compartment absorption input ($m_{abs}$), rifabutin central compartment ($C_{C,RFB}$), rifabutin peripheral compartment ($C_{P,RFB}$), des-rifabutin central compartment ($C_{C,dRFB}$), and des-rifabutin peripheral compartment ($C_{P,dRFB}$), with first-pass metabolism fraction (F_m), absorption rate constant (k_a), systemic clearances (CL_1, CL_2), intercompartmental clearances (Q_1, Q_2), and central compartment conversion clearance (Q_3).

#### L.4 Rifabutin model AUC distributions

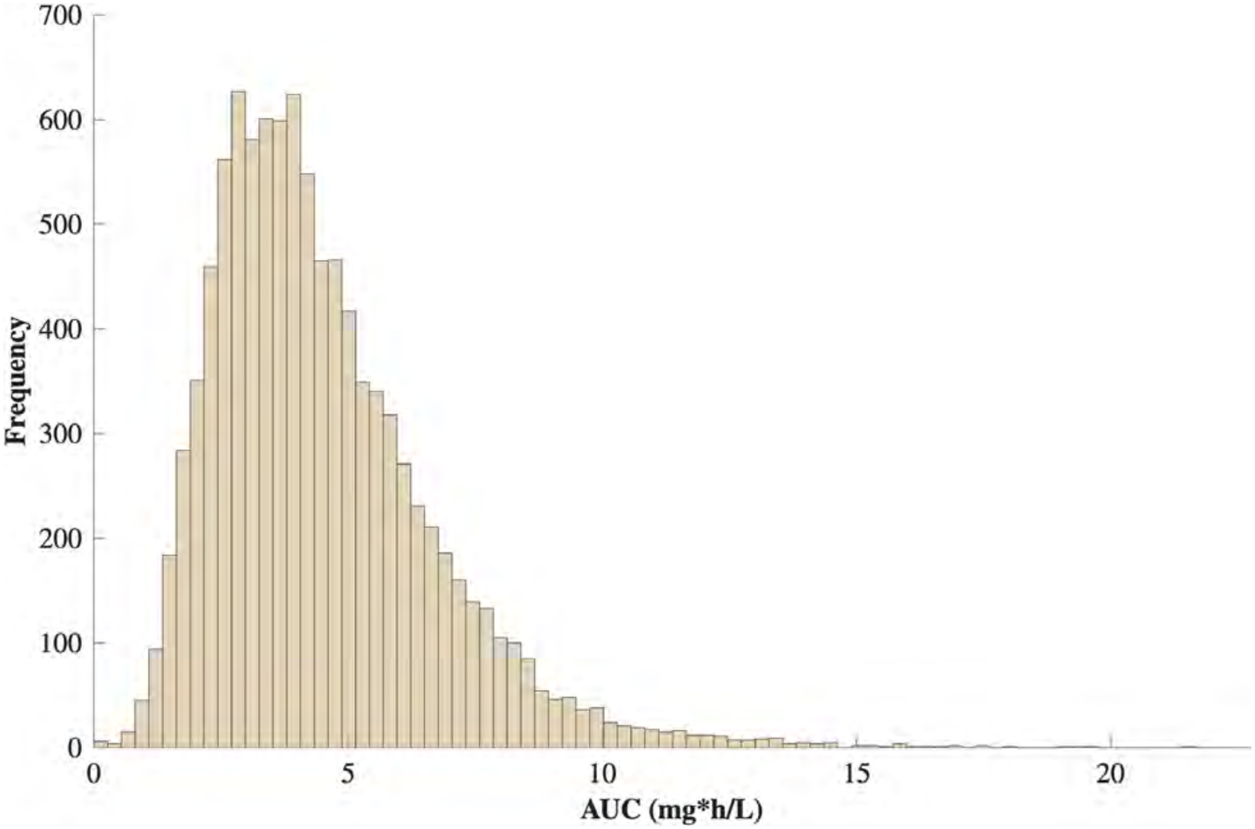

Figure S41: Steady-state $AUC_{0-24}$ distributions across 10,000 simulated patient profiles for rifabutin at 300 mg q24h. Distributions reflect interindividual variability drawn from the parameter distributions in Table S11.

#### L.5 Rifabutin model concentration time-courses

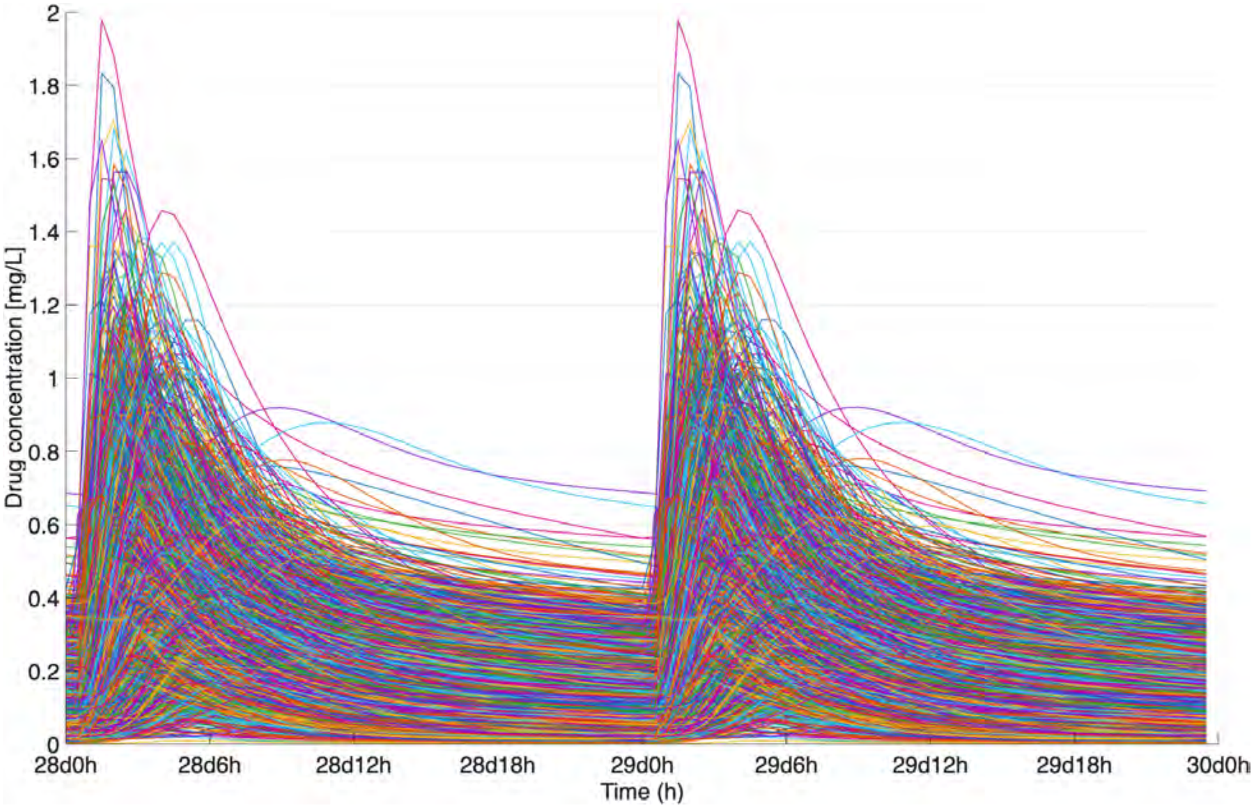

Figure S42: Simulated steady-state central compartment concentration-time profiles for rifabutin across 10,000 virtual patients receiving 300 mg q24h.

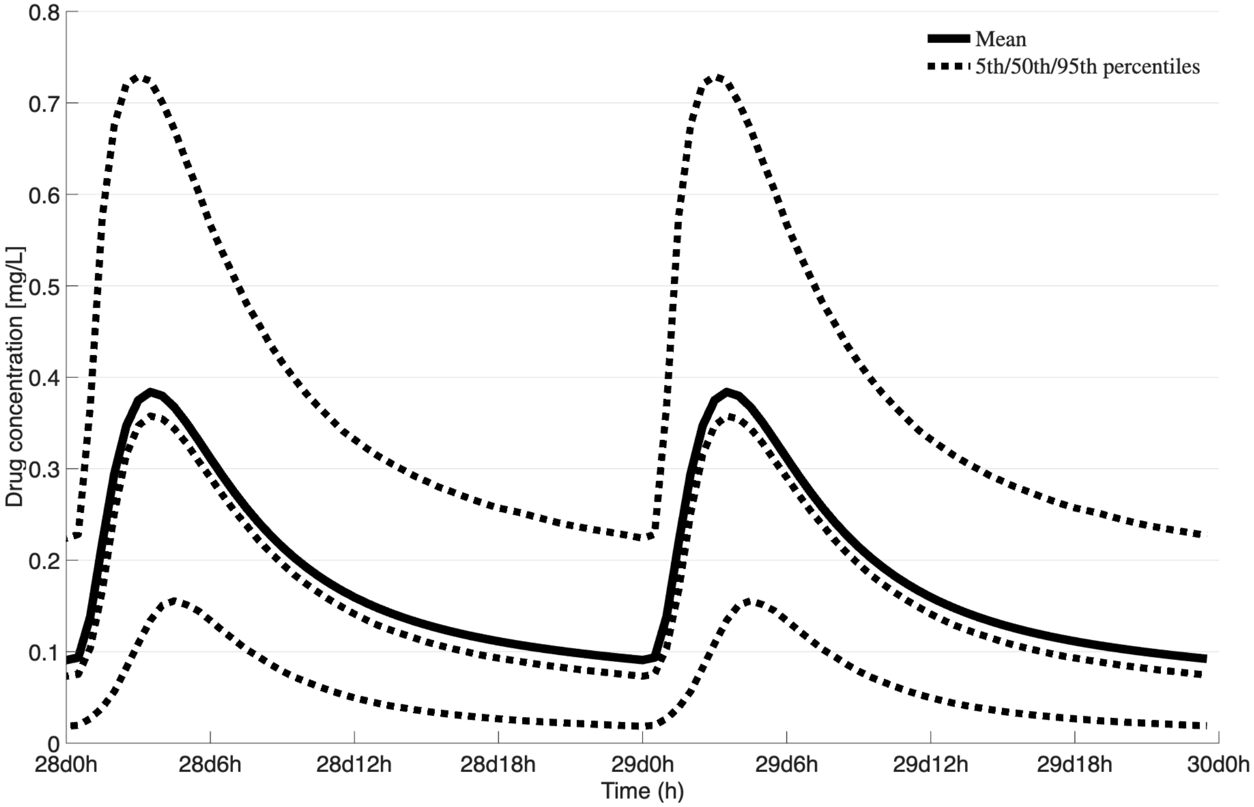

Figure S43: Simulated steady-state central compartment concentration-time profiles for rifabutin showing the mean and 90% prediction interval across 10,000 virtual patients receiving 300 mg q24h.

### M Rifampin

#### M.1 Rifampin model parameters

Table S12: Population pharmacokinetic parameters for the transit-compartment rifampin model. Values represent typical population means; $\theta$ denotes the dose-type effect coefficient, $\eta$ denotes the interindividual variability variance, and $\kappa$ denotes the interoccasional variability variance.

| Parameter | Mean | $\theta$ | $\eta$ | $\kappa$ | Units |
|---|---|---|---|---|---|
| CL/F | 19.2 | 0.236 | 0.279 | 0.0508 | L/hr |
| V/F | 53.2 | - | 0.188 | - | L/hr |
| k_a | 1.15 | - | 0.439 | - | 1/hr |
| MTT | 0.424 | 1.04 | 0.361 | 0.461 | hr |
| n | 7.13 | - | 2.44 | - | - |

CL/F represents clearance rate constant; V/F is the volume of distribution; K_a is the absorption rate constant; MTT is the mean transit time between n compartments; n is the number of theoretical transit compartments; F is bioavailability.

The parameters for this RIF model were calculated using the general form below.

$$\texttt{CL\_pt} = \texttt{CL\_mean} \cdot \left(1 + \theta_{CL/F} \cdot \texttt{dose\_type}\right) \cdot \left(\exp(\texttt{random}(\texttt{`Normal'}, 0, \sqrt{\eta})\right) \tag{43}$$

Where the parameters listed in Table S12 that have $\theta$ values include that aspect of Equation 43 and parameters that have $\eta$ values in Table S12 include that aspect of Equation 43. The $\eta$ part of Equation 43 employs an exponential value from a randomly distributed normal distribution centered around 0 with standard deviation of $\eta$. $\kappa$ values are used in the same manner as $\eta$ values, but for interoccasional variances. An example of this is shown in Equation 44

$$\texttt{CL\_pt} = \texttt{CL\_pt} \cdot \exp(\texttt{random}(\texttt{`Normal'}, 0, \sqrt{\kappa})) \tag{44}$$

#### M.2 Rifampin model equations

The Rifampin population PK model was re-implemented from [46]. Briefly, the two compartment model contains an absorption compartment ($q_0$) (Equation 47) and a central compartment ($C$) (Equation 48). The model also accounts for $n$ transit compartments.

$$\texttt{k\_tr} = \frac{\texttt{n} + 1}{\texttt{MTT}} \tag{45}$$

$$\texttt{q0\_frac} = \left(\texttt{k\_tr} \cdot t\right)^{\texttt{n}} \cdot \left(\exp\left(-\texttt{ktr} \cdot t\right)\right) / \Gamma_{\texttt{n}} \tag{46}$$

$$\frac{d(q_0)}{dt} = \texttt{dose} \cdot \texttt{F} \cdot \texttt{k\_tr} \cdot \texttt{q0\_frac} - \texttt{k\_a} \cdot (q_0) \tag{47}$$

$$\frac{d(C)}{dt} = \texttt{k\_a} \cdot (q_0) - \left(\frac{\texttt{CL\_F}}{\texttt{V\_F}}\right) \cdot (C) \tag{48}$$

##### M.3 Rifampin model flowchart

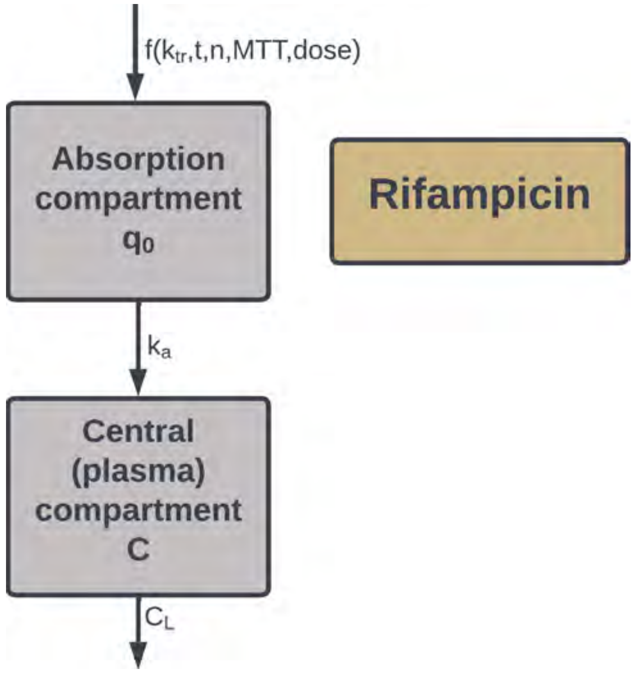

Figure S44: Schematic of the one-compartment population pharmacokinetic model for rifampin with transit-compartment absorption, showing the absorption compartment ($q_0$) and central compartment ($C$), with absorption rate constant (`k_a`), transit rate constant (`k_tr`), bioavailability (`F`), and systemic clearance (`CL/F`).

#### M.4 Rifampin model AUC distributions

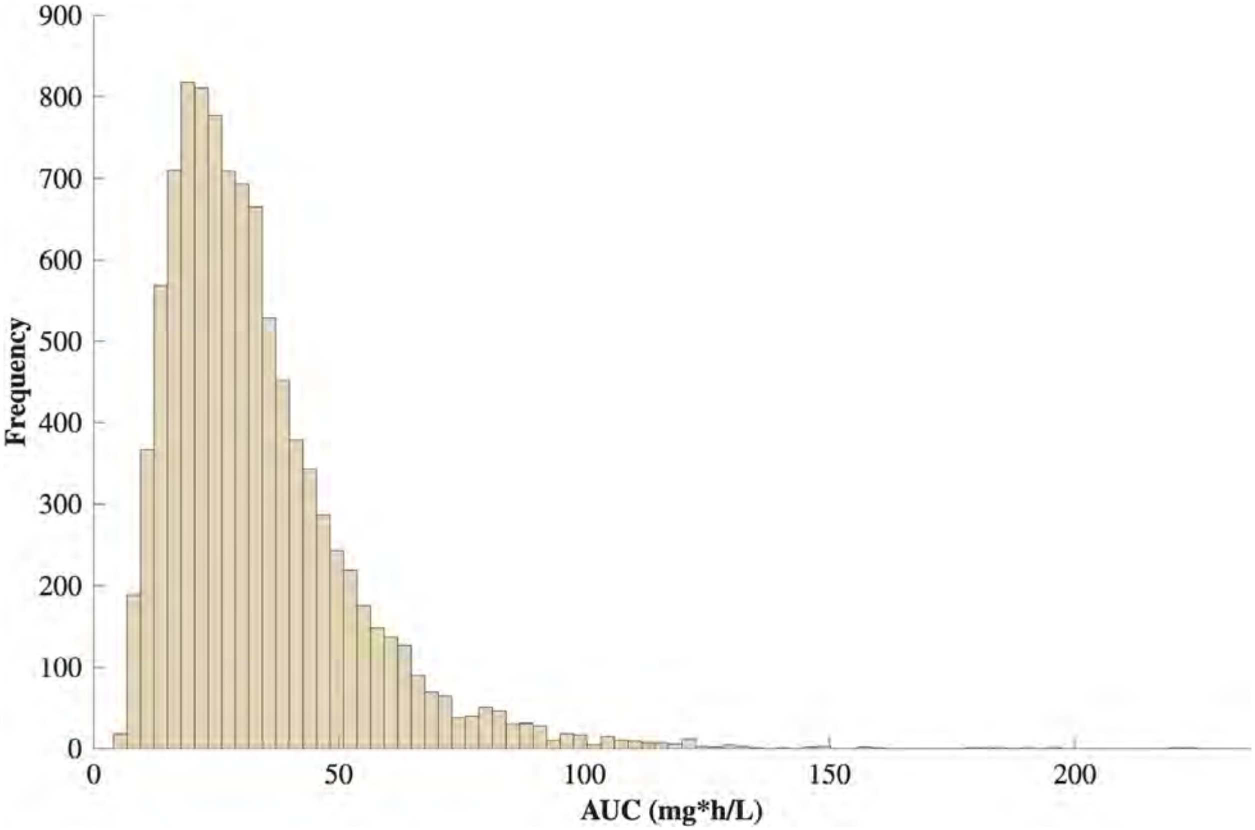

Figure S45: Steady-state $AUC_{0–24}$ distributions across 10,000 simulated patient profiles for rifampin at 600 mg q24h. Distributions reflect interindividual variability drawn from the parameter distributions in Table S12.

#### M.5 Rifampin model concentration time-courses

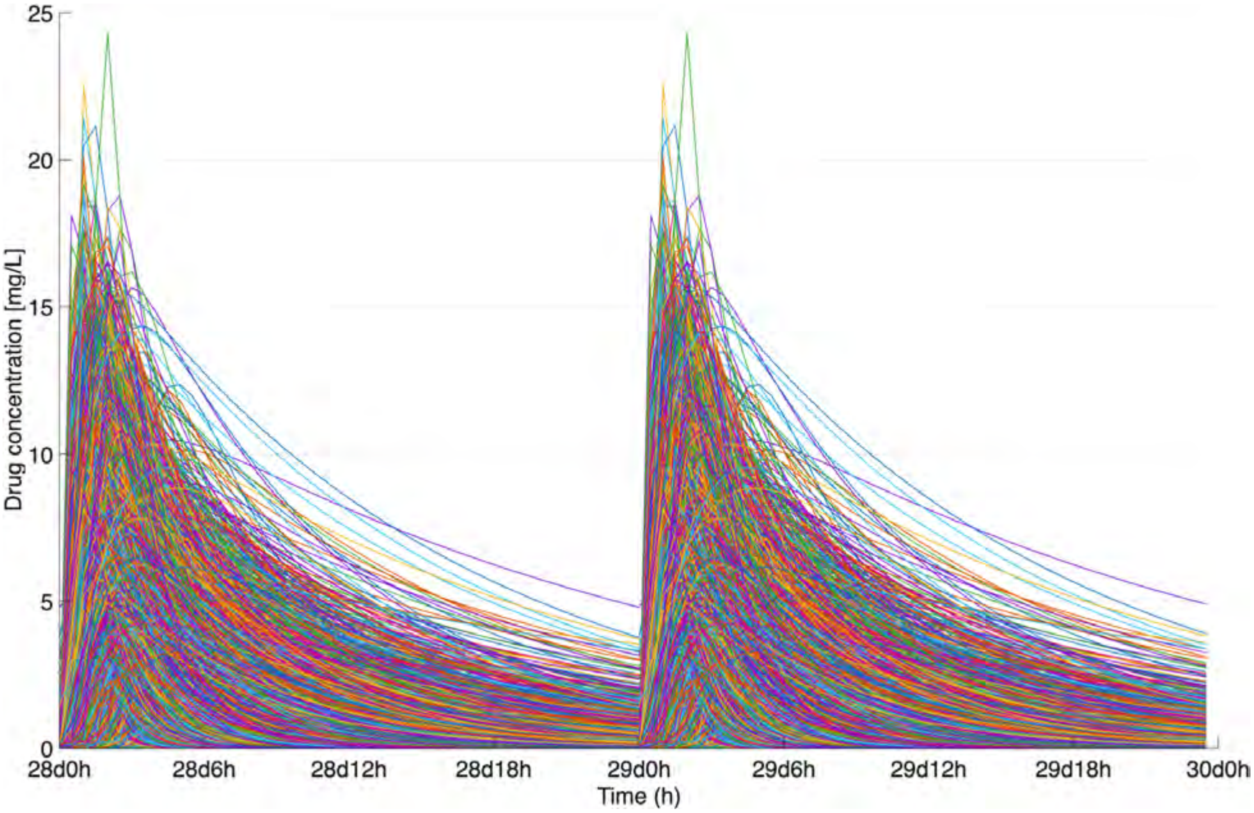

Figure S46: Simulated steady-state central compartment concentration-time profiles for rifampin across 10,000 virtual patients receiving 600 mg q24h.

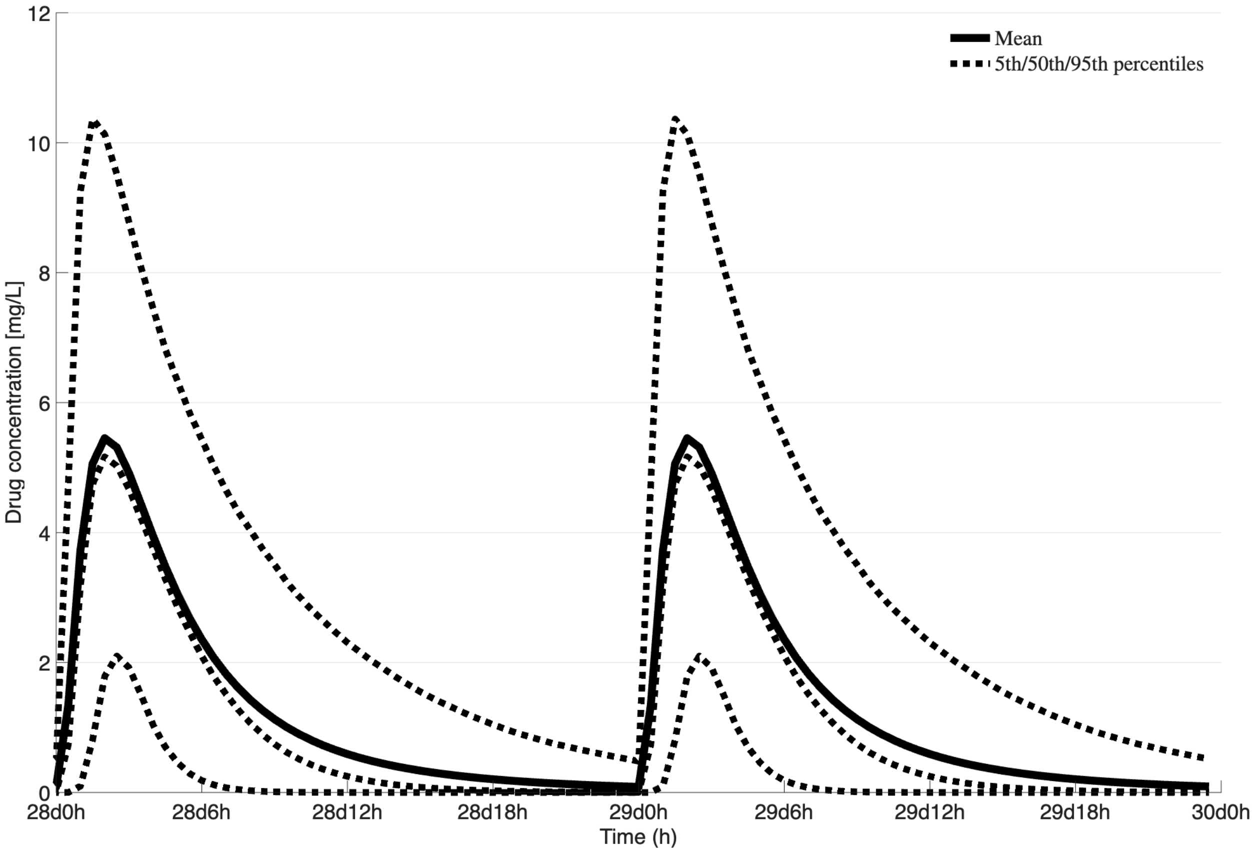

Figure S47: Simulated steady-state central compartment concentration-time profiles for rifampin showing the mean and 90% prediction interval across 10,000 virtual patients receiving 600 mg q24h.

### N Tigecycline

#### N.1 Tigecycline model parameters

Table S13: Population pharmacokinetic parameters for the two-compartment tigecycline model. Values represent typical population means; IIV is expressed as percent coefficient of variation ($\%CV$).

| Parameter | Mean | IIV % CV | Units |
|---|---|---|---|
| V_F,1 | 63.7 | 50.2 | L |
| V_F,2 | 233 | – | L |
| CL_F | 14.8 | 46.6 | L/h |
| Q_F | 38.4 | – | L/h |

V_F,1 is the central compartment volume of distribution; V_F,2 is the peripheral compartment volume of distribution; CL_F is the clearance from the central compartment; Q_F is the intercompartmental clearance between central and peripheral compartments.

#### N.2 Tigecycline model equations

This model was recreated from the model proposed in [6]. The parameter values used in the patient simulations were calculated using Equation 4. The model structure consists of a central compartment ($C$) and a peripheral compartment ($C_p$), with drug administered as a loading dose followed by maintenance doses of 25 mg q12h as a 3-hour IV infusion as described by Equation 1.

$$\frac{dC}{dt} = \texttt{infusion\_rate} - \texttt{Q\_F} \cdot \frac{C}{\texttt{V\_F,1}} + \texttt{Q\_F} \cdot \frac{C_p}{\texttt{V\_F,2}} - \texttt{CL\_F} \cdot \frac{C}{\texttt{V\_F,1}} \tag{49}$$

$$\frac{dC_p}{dt} = \texttt{Q\_F} \cdot \frac{C}{\texttt{V\_F,1}} - \texttt{Q\_F} \cdot \frac{C_p}{\texttt{V\_F,2}} \tag{50}$$

#### N.3 Tigecycline model flowchart

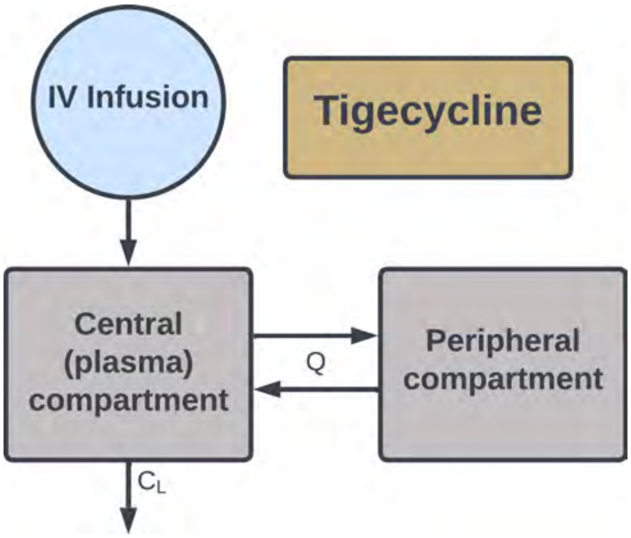

Figure S48: Schematic of the two-compartment population pharmacokinetic model for tigecycline, showing the central compartment ($C$) and peripheral compartment ($C_p$), with systemic clearance (CL_F), intercompartmental clearance (Q_F), and 3-hour IV infusion input with loading dose.

#### N.4 Tigecycline model AUC distributions

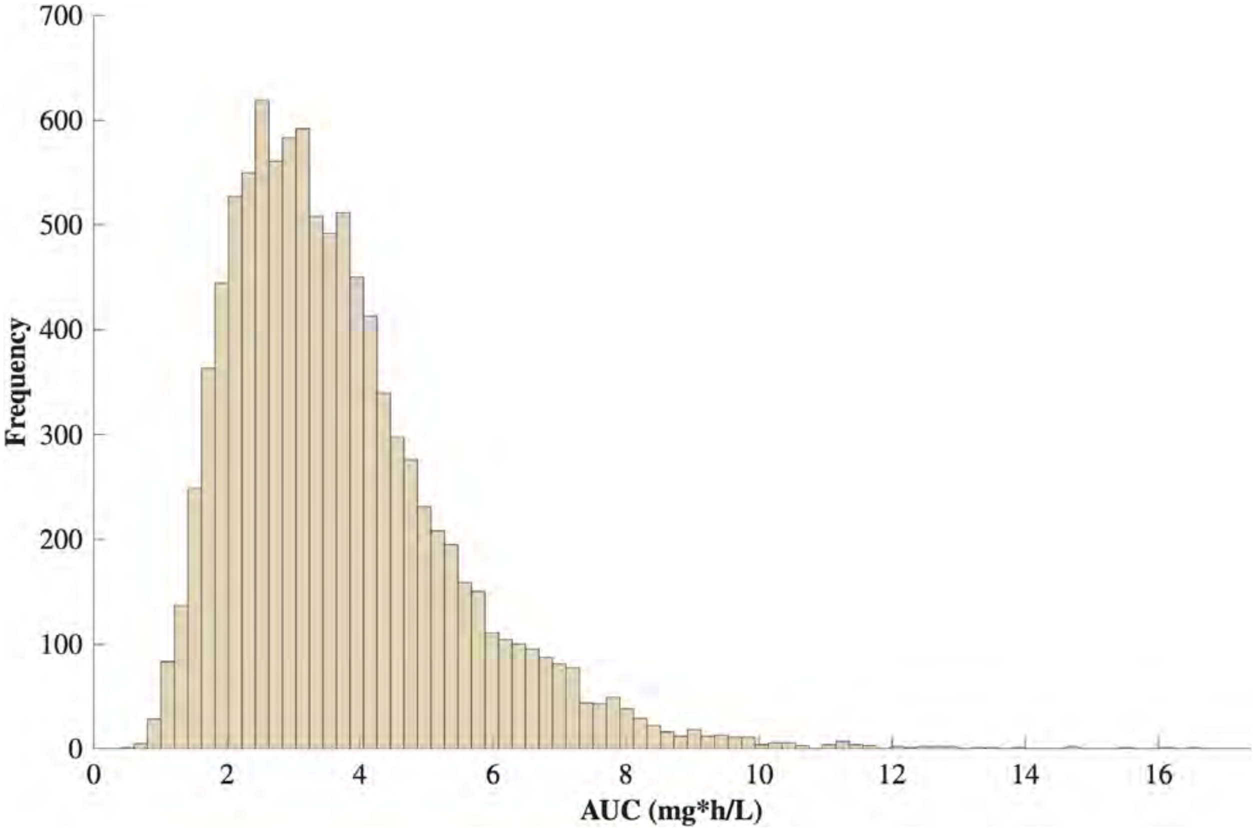

Figure S49: Steady-state $AUC_{0-24}$ distributions across 10,000 simulated patient profiles for tigecycline at 25 mg q12h administered as a 3-hour IV infusion. Distributions reflect interindividual variability drawn from the parameter distributions in Table S13.

#### N.5 Tigecycline model concentration time-courses

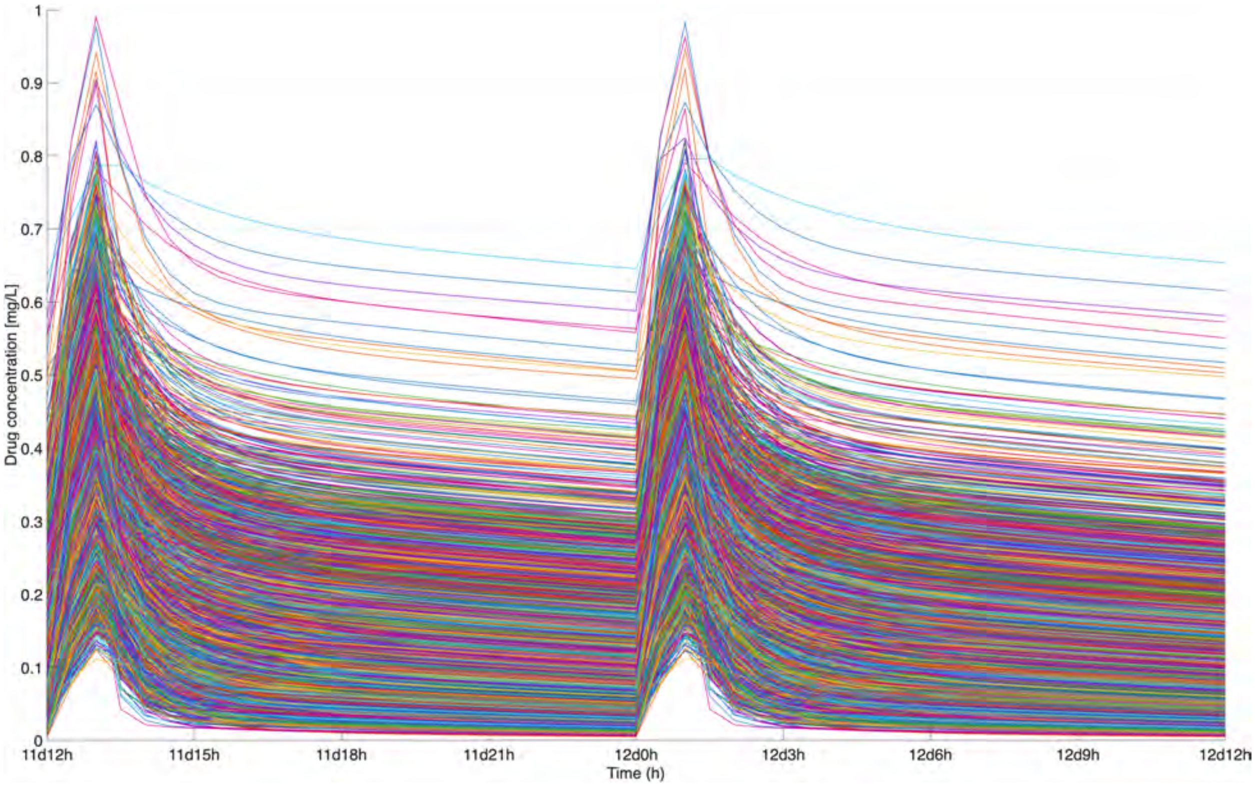

Figure S50: Simulated steady-state central compartment concentration-time profiles for tigecycline across 10,000 virtual patients receiving a loading dose followed by 25 mg q12h as a 3-hour IV infusion.

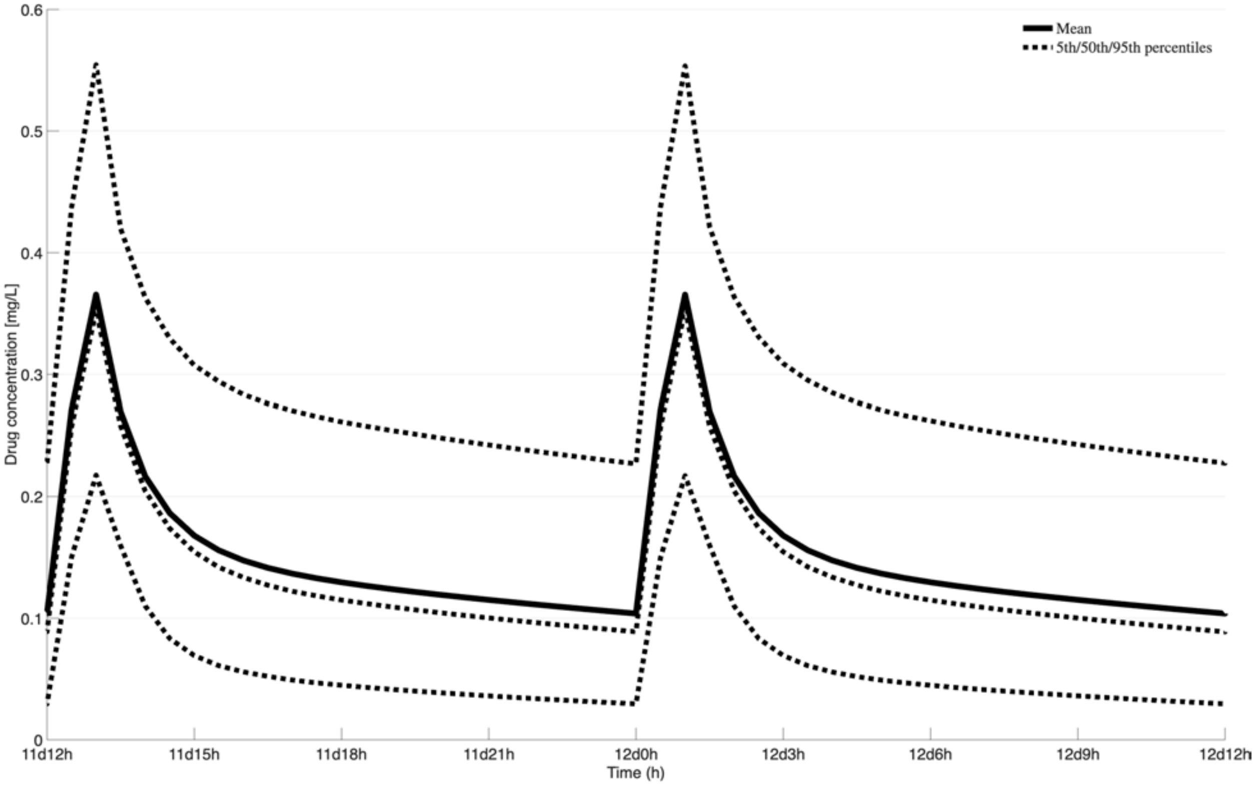

Figure S51: Simulated steady-state central compartment concentration-time profiles for tigecycline showing the mean and 90% prediction interval across 10,000 virtual patients receiving a loading dose followed by 25 mg q12h as a 3-hour IV infusion.

### O Omadacycline

#### O.1 Omadacycline model parameters

The following table represents the omadacycline parameters for 300 mg q24h [48].

Table S14: Population pharmacokinetic parameters for the two-compartment omadacycline model with parallel absorption. Values represent typical population means; IIV is expressed as percent coefficient of variation ($\%CV$).

| Parameter | Mean | IIV % CV | Units |
|---|---|---|---|
| V_F,1 | 103 | 17.0 | L |
| V_F,2 | 244 | 30.1 | L |
| CL_F,1 | 15.2 | 20.7 | L/h |
| CL_F,2 | 34.2 | 23.8 | L/h |
| k_a,1 | 0.342 | 57.2 | 1/h |
| k_a,2 | 4.58 | 72.1 | 1/h |
| P_1 | 0.786 | 10.8 | – |
| T_lag | 1.85 | 15.7 | h |

V_F,1 is the central compartment volume of distribution; V_F,2 is the peripheral compartment volume of distribution; CL_F,1 is the central compartment clearance; CL_F,2 is the intercompartmental clearance between central and peripheral compartments; k_a,1 and k_a,2 are the first-order absorption rate constants for the two parallel absorption compartments; P_1 is the fraction of dose entering the first absorption compartment; T_lag is the lag time applied to the second absorption compartment.

#### O.2 Omadacycline model equations

This model was recreated from the model proposed in [48]. The parameter values used in the patient simulations were calculated using Equation 4. The model structure was a 4-compartment model consisting of two parallel absorption compartments ($q_1, q_2$), a central compartment ($C$), and a peripheral compartment ($P$), with a lag time applied to the second absorption pathway.

$$\frac{d(q_1)}{dt} = -\texttt{k\_a,1} \cdot q_1 \tag{51}$$

$$\frac{d(q_2)}{dt} = -\texttt{k\_a,2} \cdot q_2, \quad t \geq \texttt{T\_lag} \tag{52}$$

$$\text{Rate In} = \texttt{k\_a,1} \cdot q_1 + 1_{(t \geq \texttt{T\_lag})} \cdot \texttt{k\_a,2} \cdot q_2 \tag{53}$$

$$\frac{dC}{dt} = \text{Rate In} - \texttt{CL\_F,1} \cdot \frac{C}{\texttt{V\_F,1}} - \texttt{CL\_F,2} \cdot \frac{C}{\texttt{V\_F,1}} + \texttt{CL\_F,2} \cdot \frac{P}{\texttt{V\_F,2}} \tag{54}$$

$$\frac{dP}{dt} = \texttt{CL\_F,2} \cdot \frac{C}{\texttt{V\_F,1}} - \texttt{CL\_F,2} \cdot \frac{P}{\texttt{V\_F,2}} \tag{55}$$

#### O.3 Omadacycline model flowchart

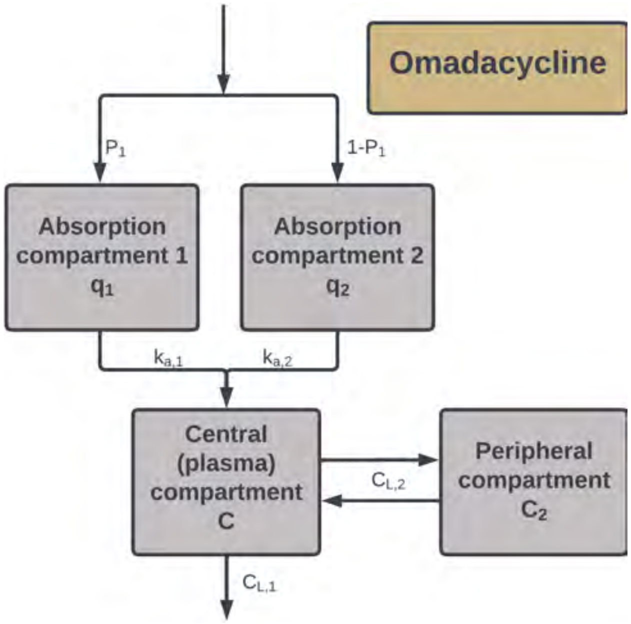

Figure S52: Schematic of the four-compartment population pharmacokinetic model for omadacycline, showing two parallel absorption compartments ($q_1, q_2$), central compartment ($C$), and peripheral compartment ($P$), with absorption rate constants (`k_a,1`, `k_a,2`), lag time (`T_lag`), systemic clearance (`CL_F,1`), and intercompartmental clearance (`CL_F,2`).

#### O.4 Omadacycline model AUC distributions

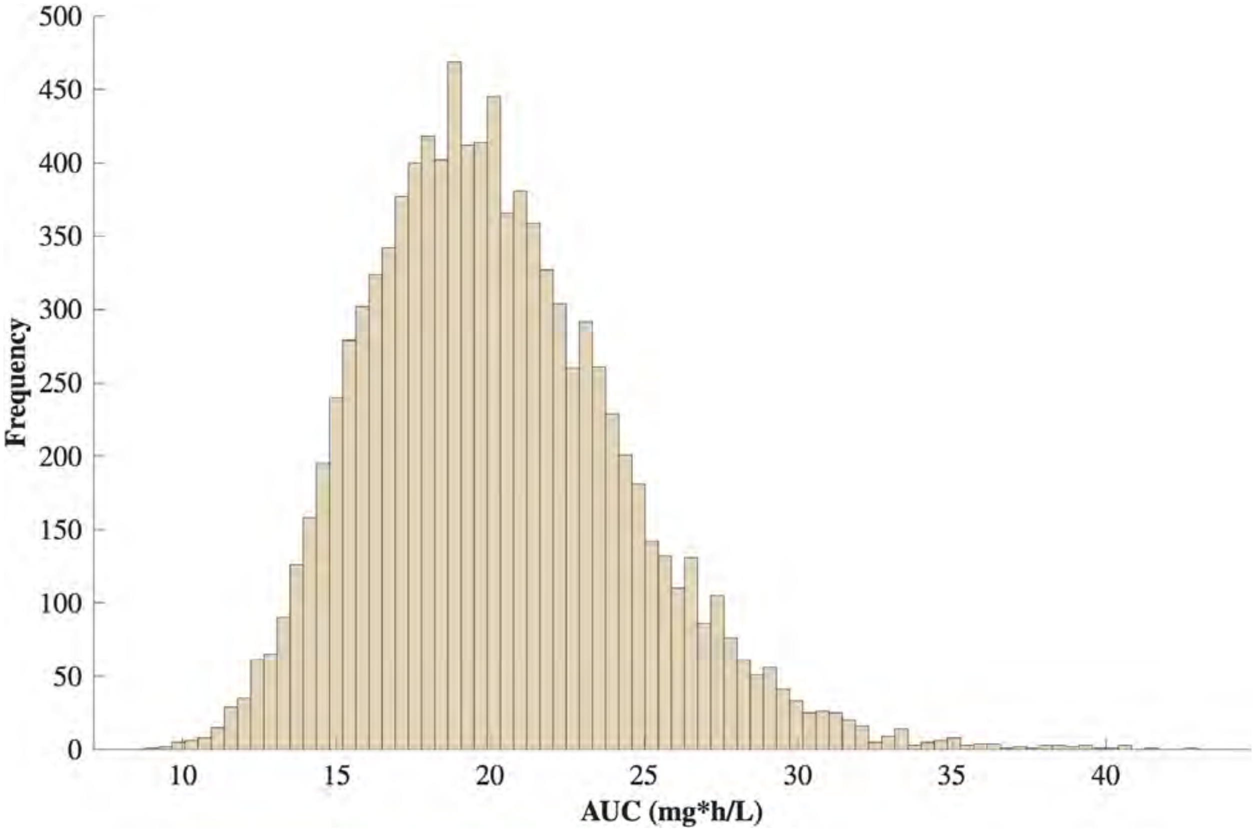

Figure S53: Steady-state $AUC_{0-24}$ distributions across 10,000 simulated patient profiles for omadacycline at 300 mg q24h. Distributions reflect interindividual variability drawn from the parameter distributions in Table S14.

#### O.5 Omadacycline model concentration time-courses

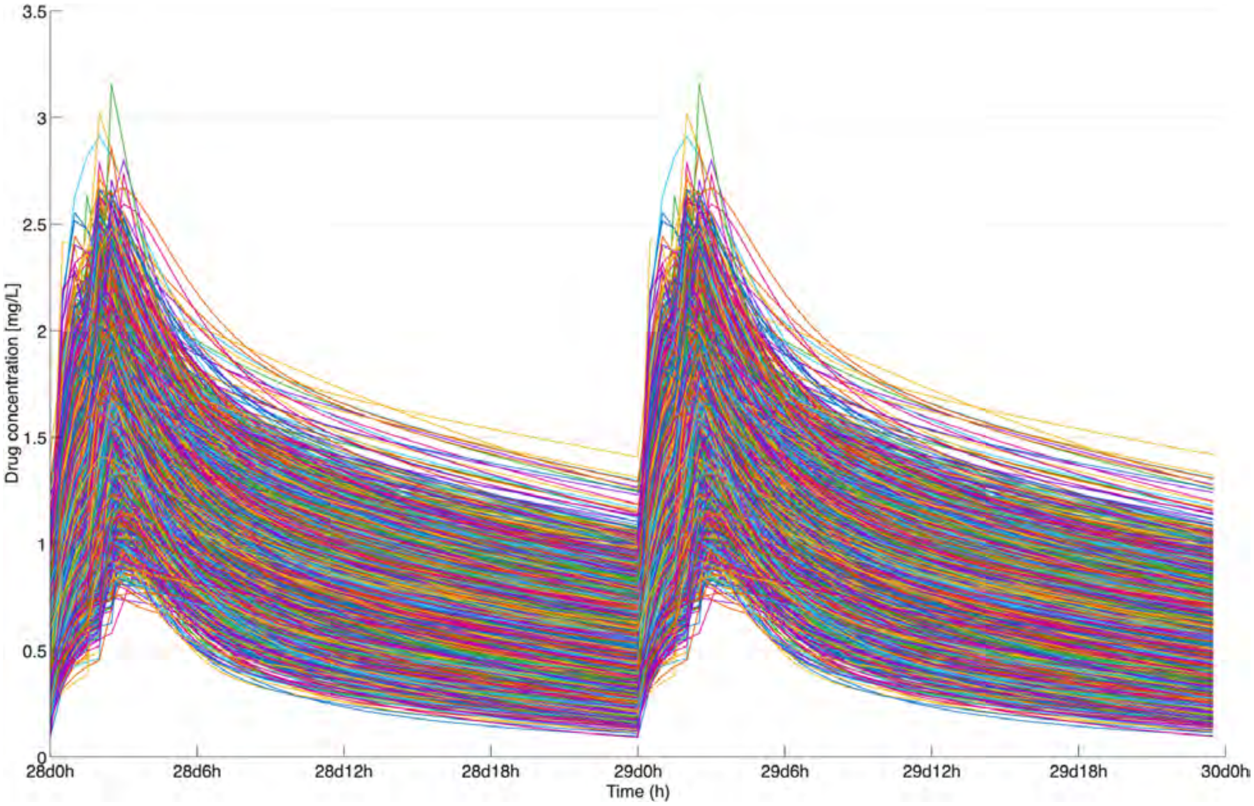

Figure S54: Simulated steady-state central compartment concentration-time profiles for omadacycline across 10,000 virtual patients receiving 300 mg q24h.

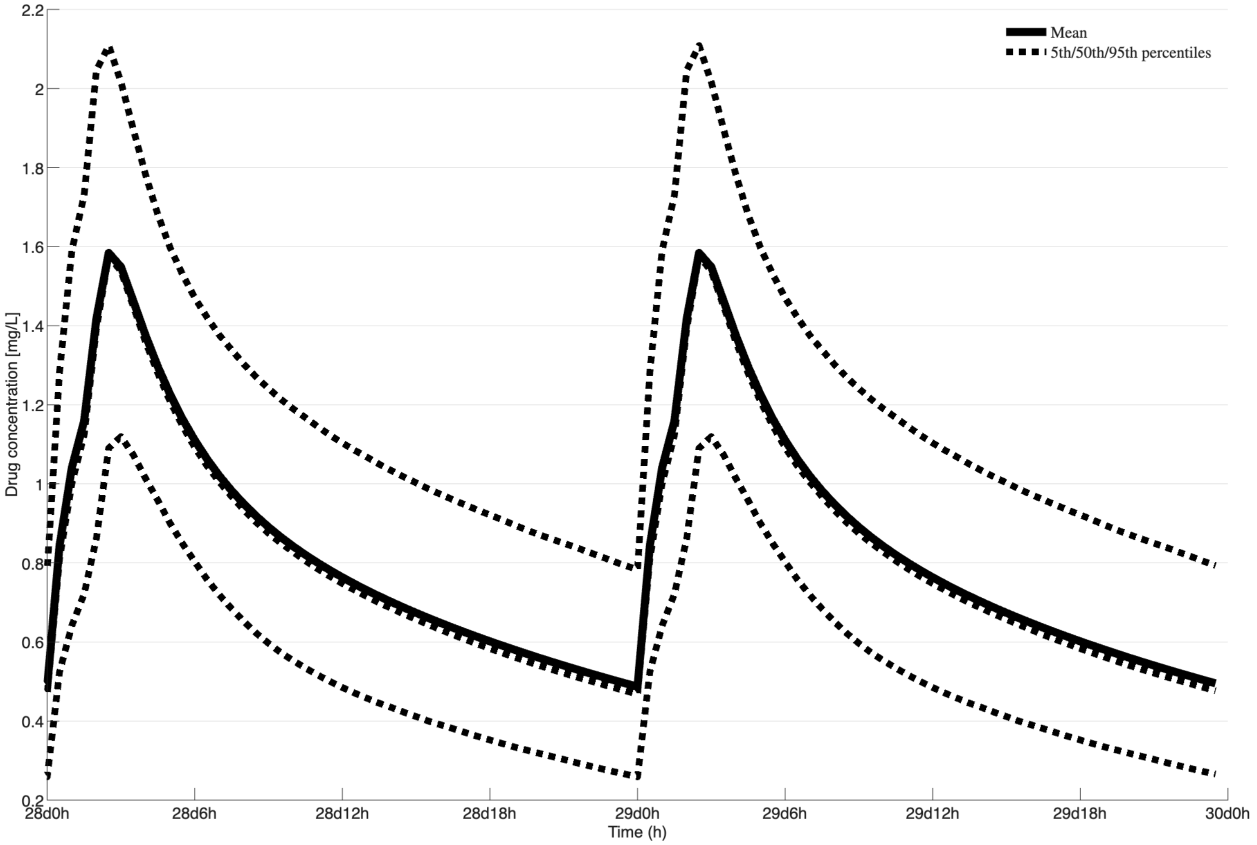

Figure S55: Simulated steady-state central compartment concentration-time profiles for omadacycline showing the mean and 90% prediction interval across 10,000 virtual patients receiving 300 mg q24h.
